# Joint contributions of metabolic dysfunction and biological aging to cardiometabolic multimorbidity and disease progression: a prospective cohort study

**DOI:** 10.64898/2026.08.24.26361219

**Authors:** Baishuang Yang, Qiong Chen, Shasha Yang

**Author notes:** ***Address Correspondence to:*** Shasha Yang, No.87, Xiangya Road, Kaifu District, Changsha, Hunan Province, China, 410008.

## Abstract

**Background:** Cardiometabolic multimorbidity (CMM), which refers to having two or more cardiometabolic conditions like type 2 diabetes, stroke, and coronary heart disease, is becoming an increasing global health challenge. Although metabolic dysfunction and biological aging may jointly contribute to CMM development, most previous studies have examined these dimensions separately. Whether their combined assessment improves risk stratification and prediction across the cardiometabolic disease continuum remains unclear.

**Methods:** This prospective cohort study involved 8,767 participants aged 45 and older who did not have CMM at the start, as part of the China Health and Retirement Longitudinal Study (CHARLS). Baseline evaluations included the triglyceride-glucose (TyG) index and two biological age algorithms, Light-BA and KDM-BA. The residual from regressing biological age on chronological age was used to derive BAA. Continuous TyG-BA composite indices were constructed as the products of TyG and biological age. Cumulative exposure and two-wave trajectory analyses used repeated measurements from 2011 and 2015. Multi-state models examined associations across the cardiometabolic disease continuum. Cox proportional hazards models, along with restricted cubic splines and time-dependent discrimination analyses, were utilized to examine associations, dose-response relationships, and incremental predictive performance.

**Results:** During a median follow-up span of 108 months, 873 participants were newly diagnosed with CMM. TyG and biological age were independently associated with CMM, with mutually adjusted hazard ratios of 1.23–1.27 and 1.39–1.44 per standard-deviation increase, respectively. Individuals with elevated TyG and rapid biological aging faced the greatest CMM risk, showing hazard ratios of 2.37 for Light-BA and 2.26 for KDM-BA, despite the absence of a significant multiplicative interaction. Continuous TyG-BA composites were associated with 49%–62% higher CMM risk per standard-deviation increase, with more than threefold higher risk in the highest versus lowest quartile and nonlinear dose-response relationships. Significantly increased CMM risk was linked to higher cumulative exposure and elevated two-wave trajectory levels, with hazard ratios ranging from 3.93 to 5.17 when comparing the highest and lowest exposure groups. Multi-state analyses demonstrated consistent associations of the composites with transitions across the cardiometabolic disease continuum and with mortality. Adding TyG-BA composites to the prespecified clinical model increased the C-index by 0.015–0.024 and improved net clinical benefit, but did not improve discrimination beyond models containing TyG and biological age as separate covariates. Associations were stronger in younger and non-frail participants in exploratory subgroup analyses.

**Conclusions:** Metabolic dysfunction and biological aging represent complementary dimensions of CMM susceptibility and progression. TyG-BA composites provide a parsimonious summary of combined metabolic-aging burden and improve risk discrimination beyond conventional clinical factors, but should not be interpreted as superior to models retaining TyG and biological age separately. These findings support the potential utility of a metabolic-aging framework for risk stratification and warrant external validation, particularly for its application in earlier stages of cardiometabolic disease development.

**Graphical Abstract:** 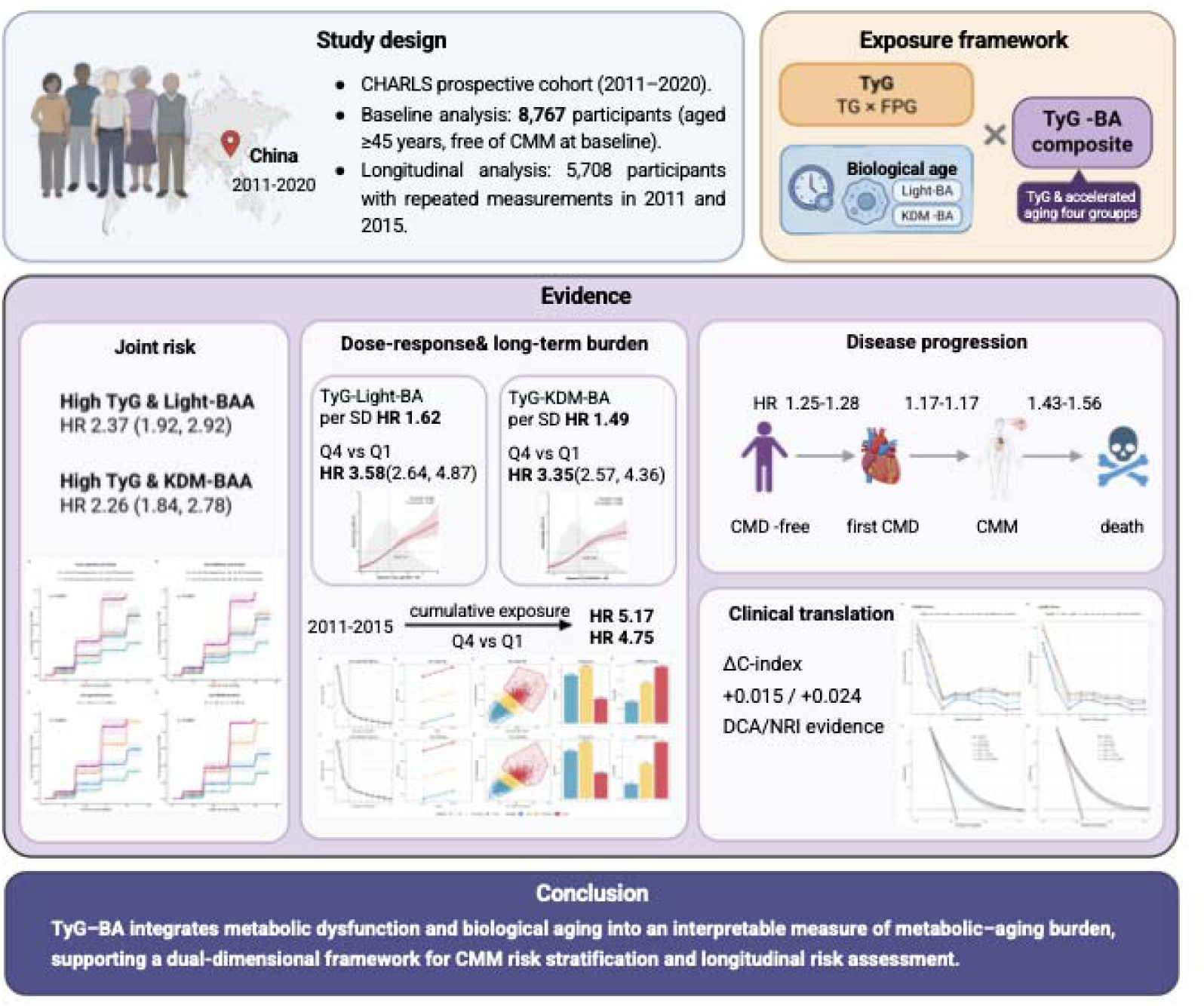

A metabolic-aging composite integrating TyG and biological age improves risk stratification for cardiometabolic multimorbidity.

**Research Insights:** **What is currently known about this topic?**

Insulin resistance and biological aging are each independently linked to CMM, but their combined contribution to risk stratification remains unexplored.

**What is the key research question?**

Does joint assessment of TyG index and biological aging improve CMM risk prediction and progression assessment across the cardiometabolic disease continuum?

**What is new?**

Metabolic dysfunction and biological aging represent complementary dimensions of CMM susceptibility and progression.

**How might this study influence clinical practice?**

Derived from routine biomarkers, the TyG-BA framework offers a practical summary measure for early CMM risk stratification, with stronger utility in younger, non-frail populations.

## 1. Introduction

Cardiometabolic multimorbidity (CMM), which typically refers to having two or more cardiometabolic diseases (CMDs) like type 2 diabetes (T2D), stroke, and coronary heart disease(CHD), is an increasing concern for public health[1, 2]. CMM is associated with greater morbidity, disability, mortality, and healthcare use than individual cardiometabolic conditions alone, and its prevalence rises with age[3–5]. Identifying high-risk individuals before multimorbidity becomes established is therefore important.

Insulin resistance (IR) is a key metabolic issue that plays a role in cardiometabolic diseases, leading to hyperglycemia, dyslipidemia, vascular dysfunction, and inflammation[6–8]. The TyG index, a straightforward indicator of insulin resistance based on fasting triglycerides and glucose, has been reliably linked to new cases of CMM across various groups, including those in the CHARLS[9–11]. Other studies comparing IR-related indices have similarly supported the association of TyG with CMM risk and its dose-response relationship[12]. However, TyG primarily captures metabolic dysfunction and may not fully reflect the broader physiological deterioration that accompanies aging.

Biological aging represents a complementary dimension of cardiometabolic vulnerability by reflecting cumulative physiological deterioration across multiple organ systems[13]. Unlike chronological age, biological age integrates molecular and clinical biomarkers reflecting inflammation, metabolic regulation, and organ function[14–16]. Accelerated biological aging, which refers to the difference between biological and chronological age, has been linked to cardiovascular incidents and death[17]. Among available approaches, Light biological age (Light-BA), based on creatinine, glucose, and C-reactive protein, and the Klemera-Doubal method biological age (KDM-BA), which utilizes various clinical biomarkers, has been used in population studies due to its reliance on routinely measured biomarkers[18–22]. Because biological-age estimates depend on the algorithm and biomarker composition used, we evaluated both approaches to test whether a metabolic-aging framework holds across complementary measures of biological aging.

Metabolic dysfunction and biological aging are biologically interconnected but represent distinct dimensions of cardiometabolic vulnerability. Metabolic stress might speed up biological aging by causing oxidative stress, mitochondrial dysfunction, and chronic low-grade inflammation[23, 24], while age-related loss of physiological reserve may increase susceptibility to metabolic and vascular insults. Nonetheless, their joint significance to CMM has not been sufficiently defined. Specifically, it is still uncertain whether integrating TyG and biological age can identify individuals at particularly high risk, whether the resulting metabolic-aging burden shows dose-response and longitudinal associations with CMM, and whether it is associated with progression across successive cardiometabolic disease states. Although multiplicative TyG-BA measures have been used as summary indicators of concurrent metabolic and aging burden, their incremental prognostic information beyond conventional clinical factors and beyond models retaining TyG and biological age as separate predictors remains uncertain.

To address these gaps, we utilized data from CHARLS, which is a cohort study representing middle-aged and older adults in China. We proposed that an increased combined burden of metabolic and biological aging would correlate with a higher risk of CMM and its progression. Specifically, we aimed to: (1) examine the independent and joint associations of TyG and biological age acceleration(BAA), assessed using Light-BA and KDM-BA, with incident CMM; (2) construct continuous TyG-BA composite measures and characterize their dose-response and longitudinal associations with CMM; (3) examine whether metabolic-aging burden was associated with transitions across the cardiometabolic disease continuum; and (4) evaluate the incremental prognostic value and potential clinical utility of the composites beyond conventional clinical factors and beyond models incorporating TyG and biological age as separate predictors.

## 2. Methods

### 2.1 Study population

The prospective analysis drew on data from CHARLS, a nationwide, multi-center community cohort study of middle-aged and older adults in China. The baseline survey was conducted in 2011 (Wave 1), with follow-up surveys conducted in 2013 (Wave 2), 2015 (Wave 3), 2018 (Wave 4), and 2020 (Wave 5)[25].

Among the 17,708 participants enrolled at Wave 1, participants were excluded according to the following criteria: (1) age <45 years (n=777); (2) missing biomarkers required for triglyceride-glucose (TyG) index(n = 5,717) or biological age estimation, including triglyceride, glucose, creatinine, C-reactive protein (CRP), or blood urea nitrogen(BUN) (n=1,815); (3) prevalent cardiometabolic multimorbidity (CMM) at or before baseline(n=371); and (4) absence of available CMM outcome information during follow-up (n=261). After these exclusions, 8,767 participants free of CMM at baseline were included in the primary analytical cohort. Participants with missing or unclassifiable cardiovascular-kidney-metabolic (CKM) syndrome stage (n=205) were retained in the analytic cohort and analyzed as a separate category in CKM subgroup analyses.

To investigate longitudinal patterns of metabolic-aging burden, a separate cumulative exposure and two-wave trajectory analysis cohort was derived from the baseline analytical cohort. Participants with available TyG and biological age measurements at both Wave 1 (2011) and Wave 3 (2015), and without prevalent CMM at or before Wave 3 were included. This resulted in a final cohort of 5,708 participants. In this cohort, cumulative TyG-biological age (TyG-BA) composite exposure and two-wave trajectory patterns were established using repeated measurements from Wave 1 to Wave 3, and incident CMM was assessed from Wave 3 through the end of follow-up in 2020. The study flow diagram is presented in Figure 1.

**Figure 1.**
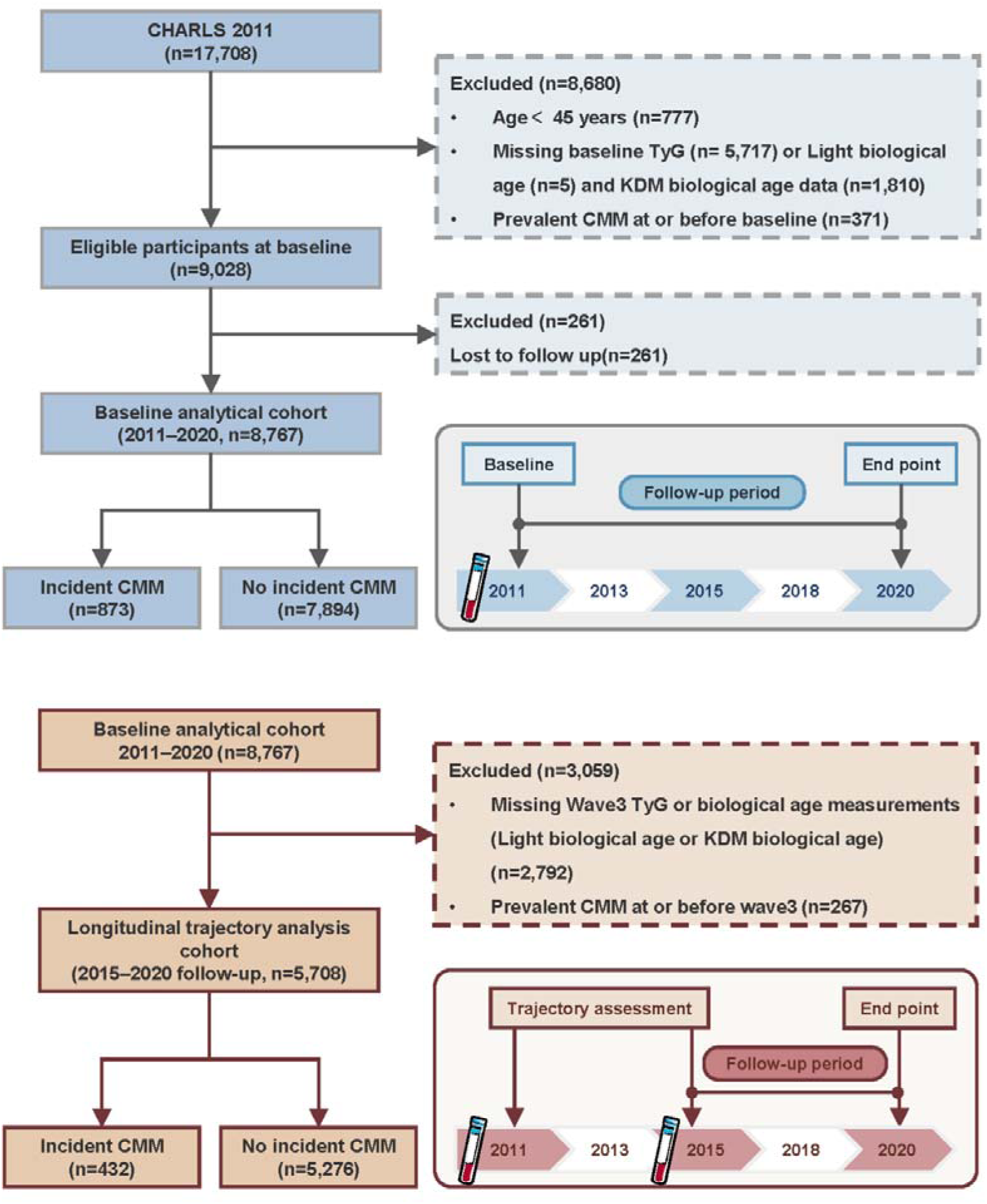
Flowchart of participant selection and follow-up in the China health and retirement longitudinal study (CHARLS), 2011–2020. *Abbreviations:* CHARLS, China Health and Retirement Longitudinal Study; TyG, triglyceride-glucose index; KDM, Klemera–Doubal method; CMM, cardiometabolic multimorbidity.

Approval for the CHARLS study was granted by Peking University’s Biomedical Ethics Review Committee (IRB00001052-11015), with all participants signing written informed consent forms.

### 2.2 CMM ascertainment

Diabetes ascertainment varied across waves because laboratory and medication data were not uniformly available. In Wave 1 and 3, diabetes was defined by self-report, medication use, HbA1c ≥ 6.5%, or fasting plasma glucose (FPG) ≥ 126 mg/dL; in Waves 2 and 4, diabetes was defined by self-report or medication use, whereas in Wave 5 it was defined by self-report only. Within each wave, missing disease components were treated conservatively as absent when at least one component was observed and as missing when all relevant components were unavailable.

Incident CMM was identified as the initial wave where the participant satisfied the CMM criteria. The event time was determined by estimating the midpoint between the last wave without CMM and the first wave with CMM. In the primary Cox proportional hazards analyses, participants who died before developing CMM were censored at the date of death; in the multi-state models, death was instead modeled as an absorbing competing event.

### 2.3 Assessment of TyG index

The TyG index served as an indirect indicator of insulin resistance and metabolic issues. It is calculated using fasting TG and FPG concentrations as follows:

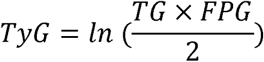

TG and FPG were expressed in mg/dL. For categorical analyses, participants were classified into low and high TyG groups according to the median TyG value in the study population (8.59).

### 2.4 Assessment of biological aging

Using a published algorithm, Light-BA was computed based on chronological age, serum creatinine, fasting glucose, and CRP[18].

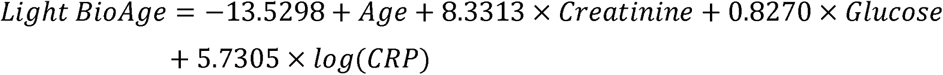

The KDM-BA was calculated using the BioAge R package [18], with previously published NHANES III training parameters and seven biomarkers: systolic blood pressure(SBP), HbA1c, total cholesterol, BUN, creatinine, platelet count, and log-transformed CRP.

Both biological age measures were retained on their original biological-age scale for analyses of biological-age level and construction of the continuous TyG-BA composite indices. BAA was derived separately to characterize the deviation of biological age from chronological-age expectations, as described below.

### 2.5 Estimation of BAA

BAA was described as the leftover from a linear regression of biological age on chronological age, specific to each sex. Specifically, for each sex, biological age was regressed on chronological age:

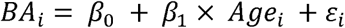

where *BA*_*i*_ represents biological age, *Age*_*i*_ represents chronological age, and *ε*_*i*_ represents the residual. The residual *ε*_*i*_ was defined as the individual’s BAA.

Positive BAA values denote a biological age greater than the expectation given an individual’s chronological age, representing accelerated biological aging; conversely, negative BAA values reflect relatively slower biological aging. Participants were dichotomized into accelerated aging (BAA□>□0) and non□accelerated aging (BAA□≤□ 0).

This residualObased method, widely adopted in epidemiological research, quantifies biological aging relative to chronologicalO age expectations instead of absolute biologicalO age levels[26]. We assessed whether BAA was independent of chronological age by inspecting their correlation (Supplementary Figure 1). The resulting BAA metrics were further implemented in categorical jointO exposure analyses.

### 2.6 Joint classification of TyG and BAA

Participants were grouped based on their baseline TyG levels and BAA status to assess the joint link between metabolic dysfunction, accelerated biological aging, and the occurrence of CMM. TyG was dichotomized using the population median, whereas BAA was classified as accelerated (BAA >0) or non-accelerated (BAA ≤0). Four mutually exclusive groups were generated: (1) low TyG with non-accelerated aging; (2) low TyG with accelerated aging; (3) high TyG with non-accelerated aging; and (4) high TyG with accelerated aging. Participants with low TyG and non-accelerated aging were used as the reference group.

This categorical framework was specifically intended to identify individuals with metabolic dysfunction accompanied by biological aging that was accelerated relative to their chronological age.

### 2.7 Construction of metabolic-aging composite indices

TyG and biological age capture complementary but distinct dimensions of cardiometabolic vulnerability. The TyG index mainly indicates insulin resistance and metabolic imbalance, whereas biological age represents an individual’s overall biological-age level based on multiple physiological biomarkers.

Two complementary biological-age metrics were therefore used for different analytical purposes. BAA was used for categorical joint classification because it quantifies deviation from age-expected biological aging and is therefore suited to identifying individuals with accelerated biological aging relative to their chronological age. In contrast, raw biological age was used to construct the continuous TyG-BA composite indices because it represents biological-age level and permits a directly interpretable multiplicative burden score.

We did not construct a multiplicative TyG×BAA score. BAA is a signed residual centered around zero; therefore, multiplication by TyG would generate both positive and negative values and would not provide a straightforward monotonic measure in which higher values consistently represent greater combined metabolic-aging burden. Accordingly, BAA was retained for categorical joint classification, whereas raw biological age was used for continuous composite construction.

Consistent with previous methodological applications of TyG × biological age[27], TyG-Light-BA and TyG-KDM-BA were calculated as the product of baseline TyG and the corresponding raw biological age measure:

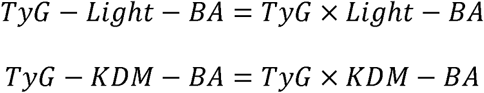

The multiplicative formulation was used as a summary measure of concurrent metabolic and biological-age burden rather than as a formal statistical interaction term. The composite indices were subsequently winsorized and standardized before regression analyses, with higher values indicating greater combined metabolic-aging burden.

Because raw biological age was expected to retain information related to chronological age, chronological age was prespecified as a covariate in the primary composite analyses. The potential influence of chronological-age adjustment on the observed associations was additionally evaluated by repeating the composite analyses without chronological-age adjustment as a sensitivity analysis.

### 2.8 Cumulative exposure and two-wave trajectory analysis of metabolic-aging composite indices (2011–2015)

Cumulative exposure to the TyG-BA composite indices was calculated from measurements obtained at Wave 1 (2011) and Wave 3 (2015) as the mean of the two composite values multiplied by the time interval between examinations:

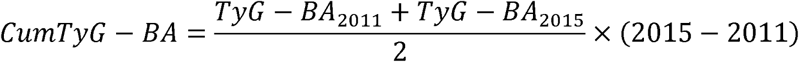

This measure followed an approach used in previous CHARLS-based analyses [25]. Because only two measurement time points were available, the resulting quantity was interpreted as an approximation of the average TyG-BA burden over 2011–2015 rather than a complete long-term exposure history. It was therefore treated as a sensitivity analysis within the Wave 3 landmark framework rather than as evidence of a causal cumulative-exposure effect.

For two-wave trajectory analyses, K-means clustering with Euclidean distance was applied to the Wave 1 and Wave 3 composite values. Three clusters (k=3) were prespecified to characterize observed two-wave exposure levels and were labeled low, moderate, and high level. Because only two measurement time points were available, these labels describe observed exposure levels rather than persistent longitudinal trajectories. The trajectory analysis was conducted as a Wave 3 landmark analysis. Cluster stability was evaluated using 200 bootstrap resamples, and k=2 and k=4 solutions were examined in sensitivity analyses.

### 2.9 Covariates

Covariates were chosen beforehand due to their possible influence as demographic, lifestyle, and clinical confounders. Demographic factors considered were age, gender, level of education (elementary or lower, middle school, college or higher), and living area (rural or urban). Lifestyle factors considered were smoking habits and alcohol use.Clinical covariates included waist circumference and HDL cholesterol. Three sequential adjustment models were specified. Model 1 took into account chronological age, sex, education, and place of residence. Model 2 further included adjustments for smoking and alcohol use. Model 3, the main model, also considered waist circumference and HDL cholesterol.

Sensitivity analyses further considered adjustment for the Frailty Index (FI), SBP, platelet count (PLT), BUN, uric acid (UA), and estimated glomerular filtration rate (eGFR), as specified for the corresponding analyses. For the continuous TyG-BA composite analyses, chronological age was retained in the primary adjustment set because chronological age is an established determinant of CMM risk and because the objective was to evaluate associations of the composite beyond chronological age. To evaluate the stability of the composite associations when adjusted for age, models excluding chronological age were also tested.

### 2.10 Missing Data

To handle missing covariate data, multiple imputation by chained equations (MICE) was used, assuming the data were missing at random. Five datasets were imputed using predictive mean matching for continuous data and logistic regression for categorical data. Each imputed dataset was analyzed separately, and the results were combined using Rubin’s rules. Complete-case analyses were used as a form of sensitivity analysis.

### 2.11 Statistical Analysis

Baseline characteristics were described using means and standard deviations for continuous variables, and frequencies and percentages for categorical ones. The characteristics were compared between joint exposure groups using either Kruskal-Wallis tests or chi-squared tests, depending on suitability.

Cox proportional hazards models were applied to calculate hazard ratios (HRs) and 95% confidence intervals (CIs) for the occurrence of CMM.TyG and raw biological age were entered separately and mutually adjusted to assess their independent associations with incident CMM. Joint exposure groups were analyzed using the low-TyG & non-accelerated aging group as the reference. In the joint exposure analyses, the multiplicative interaction was assessed by incorporating the interaction term of TyG status and BAA status into the Cox model. To assess additive interaction, the relative excess risk due to interaction (RERI), attributable proportion (AP), and synergy index (S) were used.

Cox models were employed to assess the relationships between baseline TyG-Light-BA and TyG-KDM-BA composites and the occurrence of CMM, considering the ongoing metabolic-aging burden. Composite indices were examined as standardized continuous variables and divided into quartiles.The primary composite models included chronological age as a prespecified covariate. To assess whether the associations were materially influenced by the age-related information retained in raw biological age, sensitivity analyses repeated the same models after removing chronological age from the adjustment set. The corresponding HRs and 95% CIs were compared between age-adjusted and non-age-adjusted models. The proportional hazards assumption was assessed using Schoenfeld residuals.

To explore potential nonlinear dose-response relationships, restricted cubic spline (RCS)models with three knots at the 10th, 50th, and 90th percentiles were employed. Likelihood ratio tests were used to evaluate overall and nonlinear associations. Kaplan-Meier plots and log-rank analyses were employed to illustrate and contrast CMM-free survival among combined exposure groups, composite quartiles, and two-wave trajectory groups.

Cumulative TyG-BA exposure and two-wave trajectory membership were evaluated within the Wave 3 landmark cohort. Cumulative exposure was modeled using the mean of the Wave 1 and Wave 3 composite values multiplied by the interval between examinations, as described above. Because only two measurements were available, cumulative exposure was interpreted as an approximation of average exposure over 2011-2015. K-means-derived two-wave exposure-level groups were similarly evaluated using Cox models.

Multi-state models were used to examine associations across the observed cardiometabolic disease continuum using a full illness-death state space comprising four states (CMD-free, first CMD, CMM, and death) and five transitions: CMD-free to first CMD, CMD-free to death, first CMD to CMM, first CMD to death, and CMM to death. Death was modeled as an absorbing competing event from each non-absorbing state rather than as censoring. Participants with one prevalent CMD at baseline entered the first CMD state at time 0, and first CMD event time was assigned using the same wave-to-wave midpoint imputation as CMM. Multi-morbid onset contributed to each involved subtype in subtype-specific analyses. The Markov assumption was assessed by including the global entry time into the current state as a covariate in transition-specific Cox models; transitions from the initial CMD-free state were not testable because all participants entered that state at time 0. Clock-reset (semi-Markov) models were fitted as sensitivity analyses; the corresponding results are reported in the Supplementary Materials.

Fine-Gray subdistribution hazard models were employed to consider all-cause mortality as a competing event. Analyses of subgroups were performed based on age, sex, body mass index (BMI), residence, education, smoking, alcohol consumption, hypertension, hyperlipidemia, CKM stage, and FI category. Interaction tests were performed using pooled Wald or likelihood ratio tests. Frailty effect modification was assessed by stratifying participants into robust (FI <0.10), pre-frail (FI 0.10–0.25), and frail (FI ≥0.25) categories[28], using Fine-Gray models without FI adjustment. FI was evaluated as a potential effect modifier rather than a conventional confounder because of substantial conceptual and component overlap with the exposure. Due to the numerous subgroup comparisons, these analyses were viewed as exploratory and aimed at generating hypotheses rather than confirming them. CKM stage was similarly treated as a stratification variable for evaluating risk heterogeneity rather than as a mechanistic indicator.

The incremental predictive value of the TyG-BA composites was assessed by comparing models with and without the composite measures using IPCW-adjusted ΔC-index, continuous NRI, and IDI at 96 months of follow-up. The 96-month horizon was prespecified as the primary prediction time point, with ΔC-index analyses at 60 and 108 months performed as sensitivity analyses. Clinical net benefit was assessed using decision curve analysis across various threshold probabilities throughout the entire 108-month follow-up period. Time-dependent ROC analyses were additionally performed at 3, 5, and 8 years using the timeROC package, with AUCs and 95% CIs estimated using marginal IPCW weighting. Pairwise comparisons between each marker and TyG were conducted using the iid-based comparison procedure implemented in timeROC, with *P* values adjusted using the Benjamini–Hochberg (BH) procedure. Detailed procedures for IPCW estimation, multiple-imputation pooling, model comparisons, and Kernel SHAP analyses are provided in the Supplementary Methods.

Multicollinearity was assessed in the final regression models, including the TyG-BA composite and chronological age, using variance inflation factors (VIFs). All VIFs were <5, indicating no substantial multicollinearity.

Every analysis was conducted in R version 4.5.2 using the mice, survival, cmprsk, mstate, rms, survIDINRI, kernelshap, and related packages. All statistical analyses were conducted as two-tailed tests with a significance threshold of 0.05. A complete list of analysis scripts is provided in the Supplementary Materials.

## 3. Results

### 3.1 Study Population and Baseline Characteristics

The primary analytical cohort included 8,767 participants free of CMM at baseline, among whom 873 (10.0%) developed incident CMM during a median follow-up of 108 months (IQR, 96.5-108.0). The baseline characteristics of participants according to TyG and BAA status are presented in Table 1 for Light-BA and Supplementary Table 1 for KDM-BA.

**Table 1.**
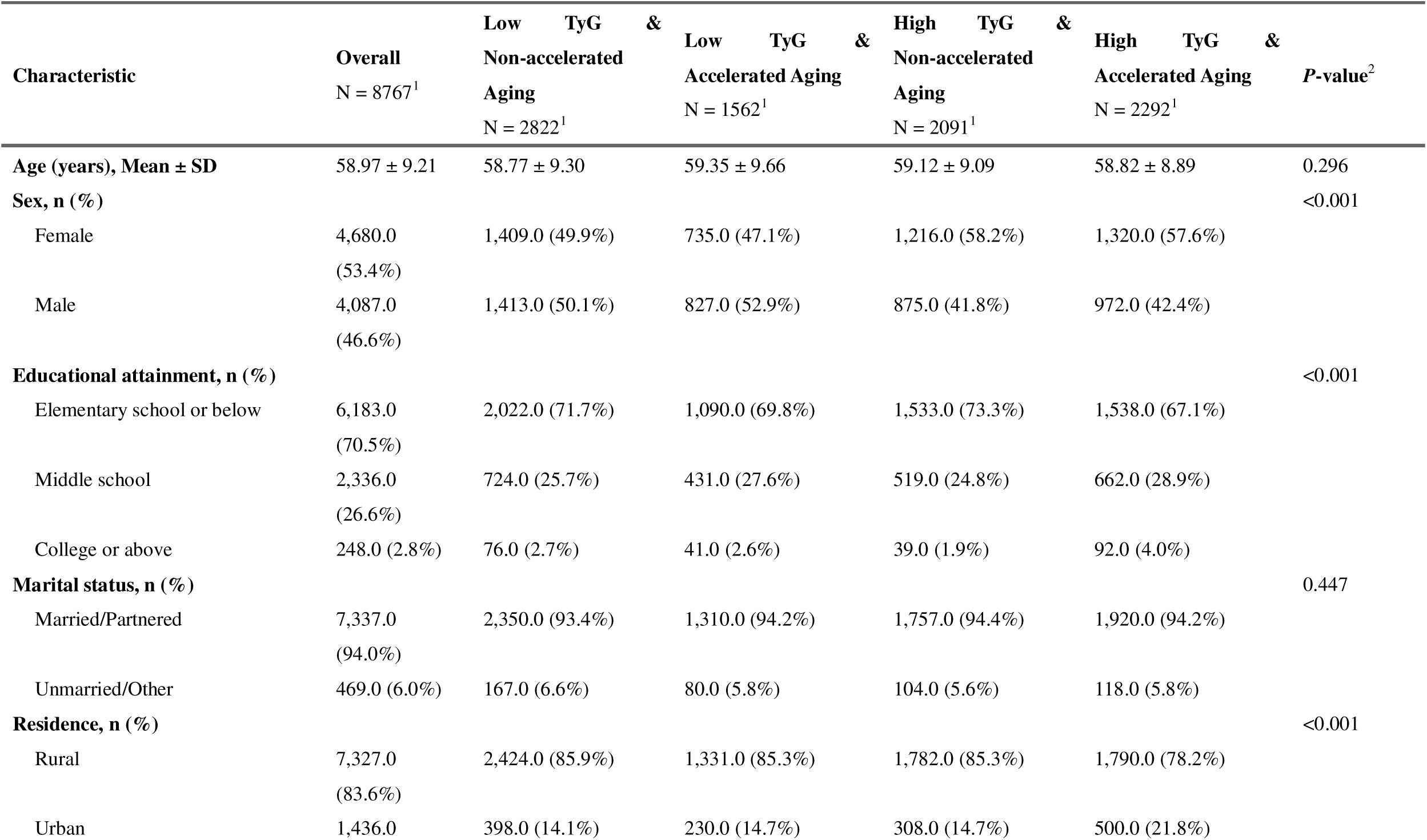
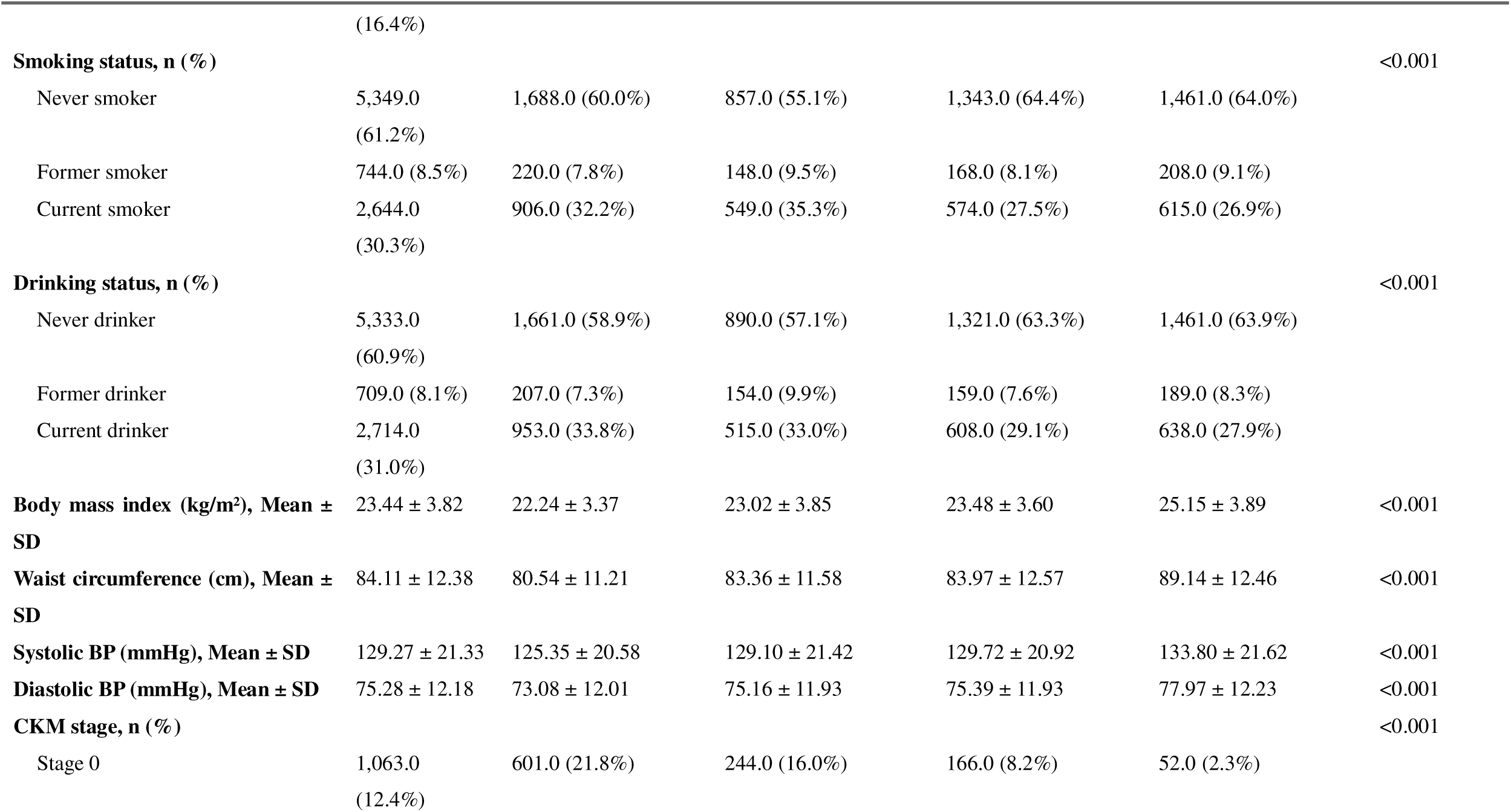
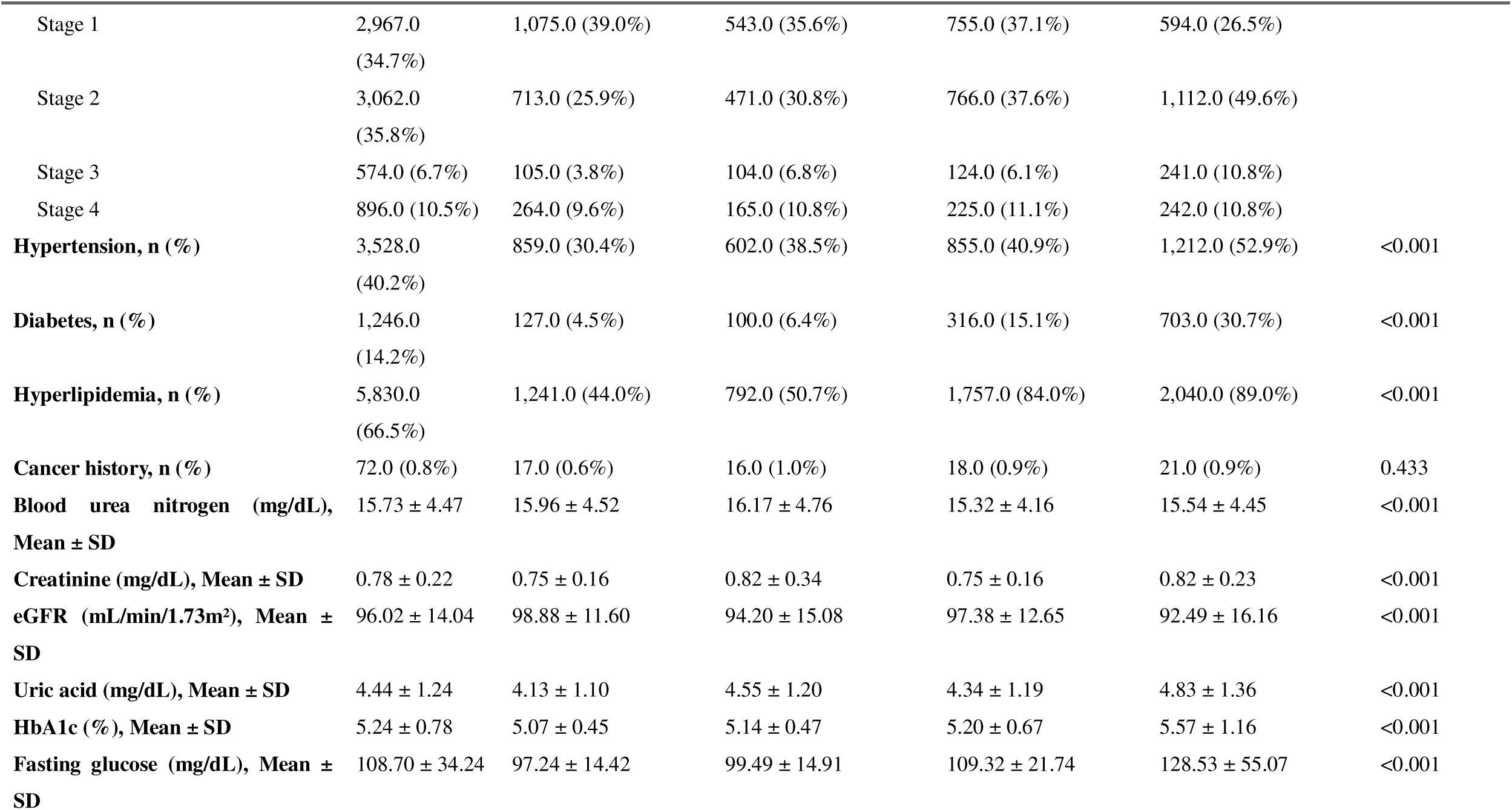
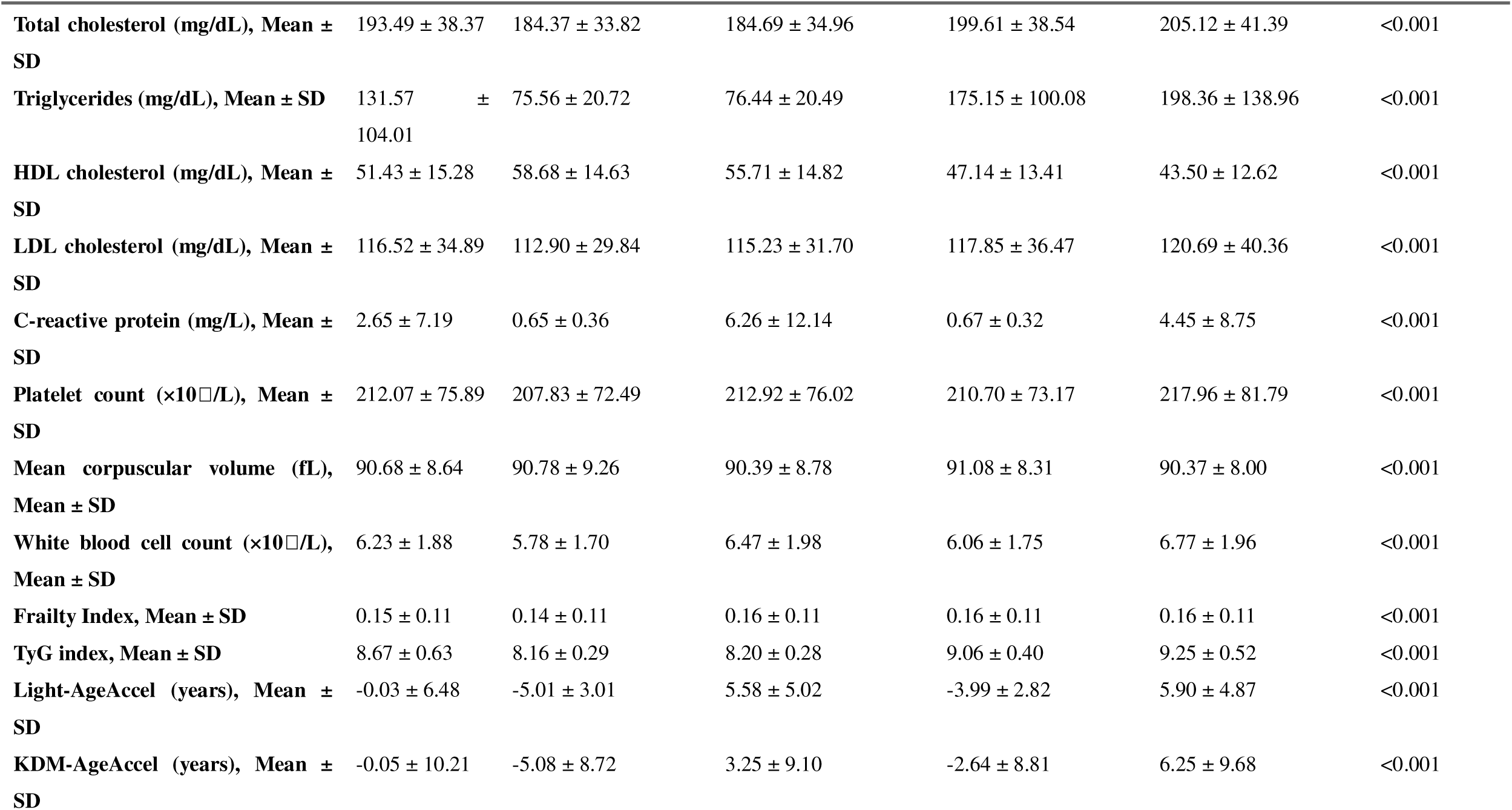
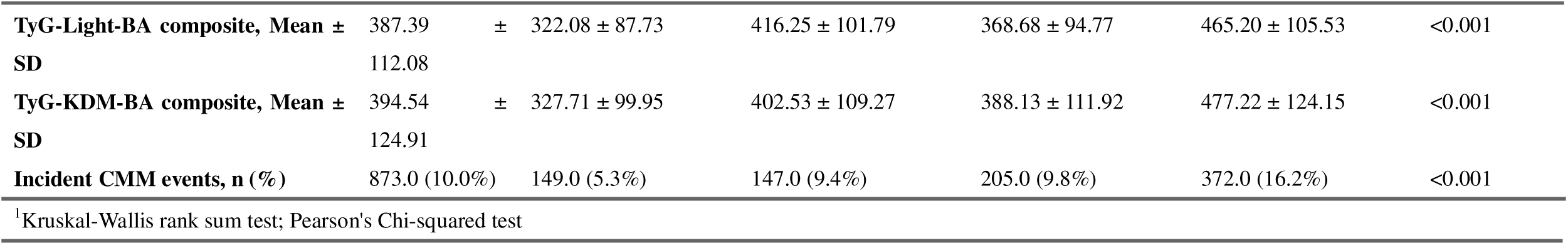
Baseline characteristics of the study population according to joint TyG index and Light biological age acceleration groups. Data are presented as mean (SD) for continuous variables and n (%) for categorical variables. Joint groups were defined by median TyG (8.67) and Light□age acceleration (cutoff: 0 years). P values from Kruskal–Wallis or chi□squared tests across groups. *Abbreviations:* TyG, triglyceride-glucose index; Light-BA, Light-method biological age; Light-AgeAccel, Light-method biological age acceleration; CKM, cardiovascular-kidney-metabolic; eGFR, estimated glomerular filtration rate; HbA1c, glycated hemoglobin; HDL, high-density lipoprotein; LDL, low-density lipoprotein; SD, standard deviation.

Biological age showed substantial age-related information, with strong positive correlations between chronological age and both KDM-BA and Light-BA (KDM-BA: R = 0.62; Light-BA: R = 0.834; both *P* < 0.001; Supplementary Figure 1). In contrast, residual-derived BAA measures showed essentially no linear correlation with chronological age (R ≈ 0, both *P* < 0.001), confirming that the residualization procedure effectively removed the linear component attributable to chronological age. These findings supported the use of BAA to characterize aging relative to age-expected biological age, while retaining BA as a measure of biological-age level for continuous composite analyses.

Individuals with elevated TyG and rapid biological aging showed the worst cardiometabolic characteristics, such as increased BMI, waist size, blood pressure, fasting glucose, HbA1c, triglycerides, and CRP levels, as well as higher prevalences of diabetes, hypertension, and hyperlipidemia (all *P* < 0.001). This group also had more advanced CKM stages at baseline. During follow-up, the incidence of CMM increased progressively across the four joint categories, from 5.3% among participants with low TyG and non-accelerated aging to 16.2% among those with high TyG and accelerated aging (*P* < 0.001).

### 3.2 Independent contributions of metabolic dysfunction and biological age to incident CMM

We first examined whether metabolic dysfunction and biological aging were independently associated with incident CMM when considered simultaneously. In mutually adjusted Cox models, both TyG and biological age remained positively associated with incident CMM (Table 2).

**Table 2.** Independent contributions of TyG index and biological aging to incident cardiometabolic multimorbidity after mutual adjustment. HRs and 95% CIs per 1□SD increase, estimated from Cox models with mutual adjustment for TyG and biological age in the same model. Crude model: no covariates. Model 1: age, sex, education, residence. Model 2: Model 1 + smoking, alcohol. Model 3 (primary): Model 2 + waist circumference, HDL□C. *Abbreviations:* HR, hazard ratio; CI, confidence interval; TyG, triglyceride-glucose index; Light-BA, Light-method biological age; KDM-BA, Klemera–Doubal method biological age; SD, standard deviation.

| Variable | Crude Model |  | Model 1 |  | Model 2 |  | Model 3 |  |
| --- | --- | --- | --- | --- | --- | --- | --- | --- |
|  | HR (95% CI) | *P* value | HR (95% CI) | *P* value | HR (95% CI) | *P* value | HR (95% CI) | *P* value |
| **TyG index and Light-BA mutually adjusted** |  |  |  |  |  |  |  |  |
| TyG index (per SD) | 1.40 (1.32, 1.49) | < 0.001 | 1.36 (1.28, 1.45) | < 0.001 | 1.36 (1.28, 1.45) | < 0.001 | 1.27 (1.18, 1.37) | < 0.001 |
| Light-BA (per SD) | 1.39 (1.30, 1.48) | < 0.001 | 1.55 (1.38, 1.74) | < 0.001 | 1.54 (1.37, 1.73) | < 0.001 | 1.44 (1.28, 1.63) | < 0.001 |
| **TyG index and KDM-BA mutually adjusted** |  |  |  |  |  |  |  |  |
| TyG index (per SD) | 1.35 (1.27, 1.44) | < 0.001 | 1.33 (1.25, 1.42) | < 0.001 | 1.33 (1.25, 1.42) | < 0.001 | 1.23 (1.14, 1.32) | < 0.001 |
|  | HR (95% CI) | P value | HR (95% CI) | P value | HR (95% CI) | P value | HR (95% CI) | P value |
| KDM-BA (per SD) | 1.46 (1.37, 1.56) | < 0.001 | 1.48 (1.36, 1.61) | < 0.001 | 1.48 (1.36, 1.61) | < 0.001 | 1.39 (1.27, 1.52) | < 0.001 |

Abbreviations: HR, Hazard Ratio; CI, Confidence Interval; TyG, Triglyceride-Glucose index; BA, Biological Age.
Crude Model: Mutually adjusted for TyG and Biological Age without additional covariates.
Model 1: Adjusted for age, sex, education attainment, and residence.
Model 2: Model 1 + smoking status and alcohol consumption.
Model 3: Model 2 + waist circumference, HDL-C.

In Model 3, each 1-SD increase in TyG was associated with a 27% higher risk of incident CMM when adjusted for Light-BA (HR, 1.27; 95% CI, 1.18–1.37) and a 23% higher risk when adjusted for KDM-BA (HR, 1.23; 95% CI, 1.14–1.32). Conversely, each 1-SD increase in Light-BA was associated with an HR of 1.44 (95% CI, 1.28–1.63), while each 1-SD increase in KDM-BA was associated with an HR of 1.39 (95% CI, 1.27–1.52). These findings indicated that metabolic dysfunction and biological-age level each contributed to CMM risk after accounting for the other dimension, providing the basis for subsequently evaluating their combined phenotypes and continuous composite burden.

### 3.3 Joint association of metabolic dysfunction and accelerated biological aging with incident CMM

Next, we explored whether the combination of metabolic dysfunction and accelerated biological aging, as opposed to chronological age, was related to a notably high risk of CMM. Using the low TyG & non-accelerated aging group as the reference, CMM risk increased across the four joint categories (Table 3).

**Table 3.**
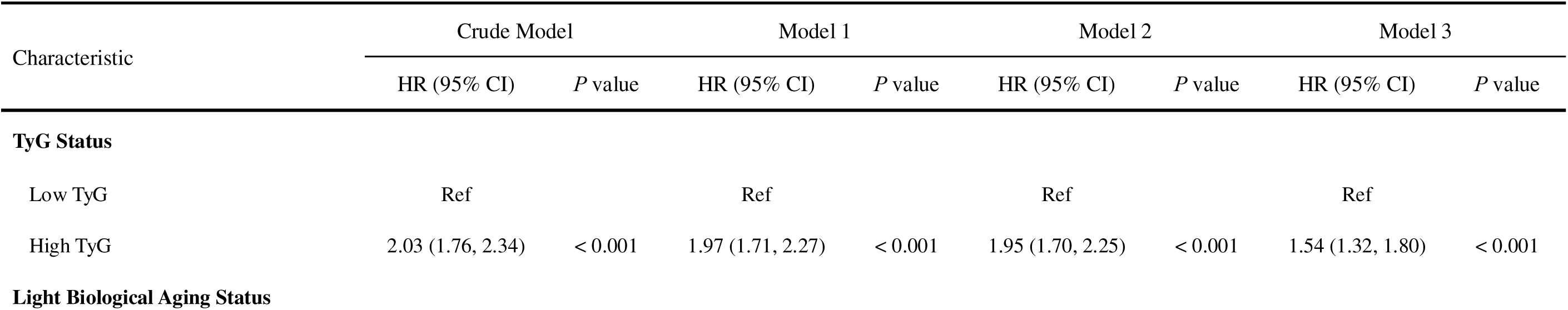
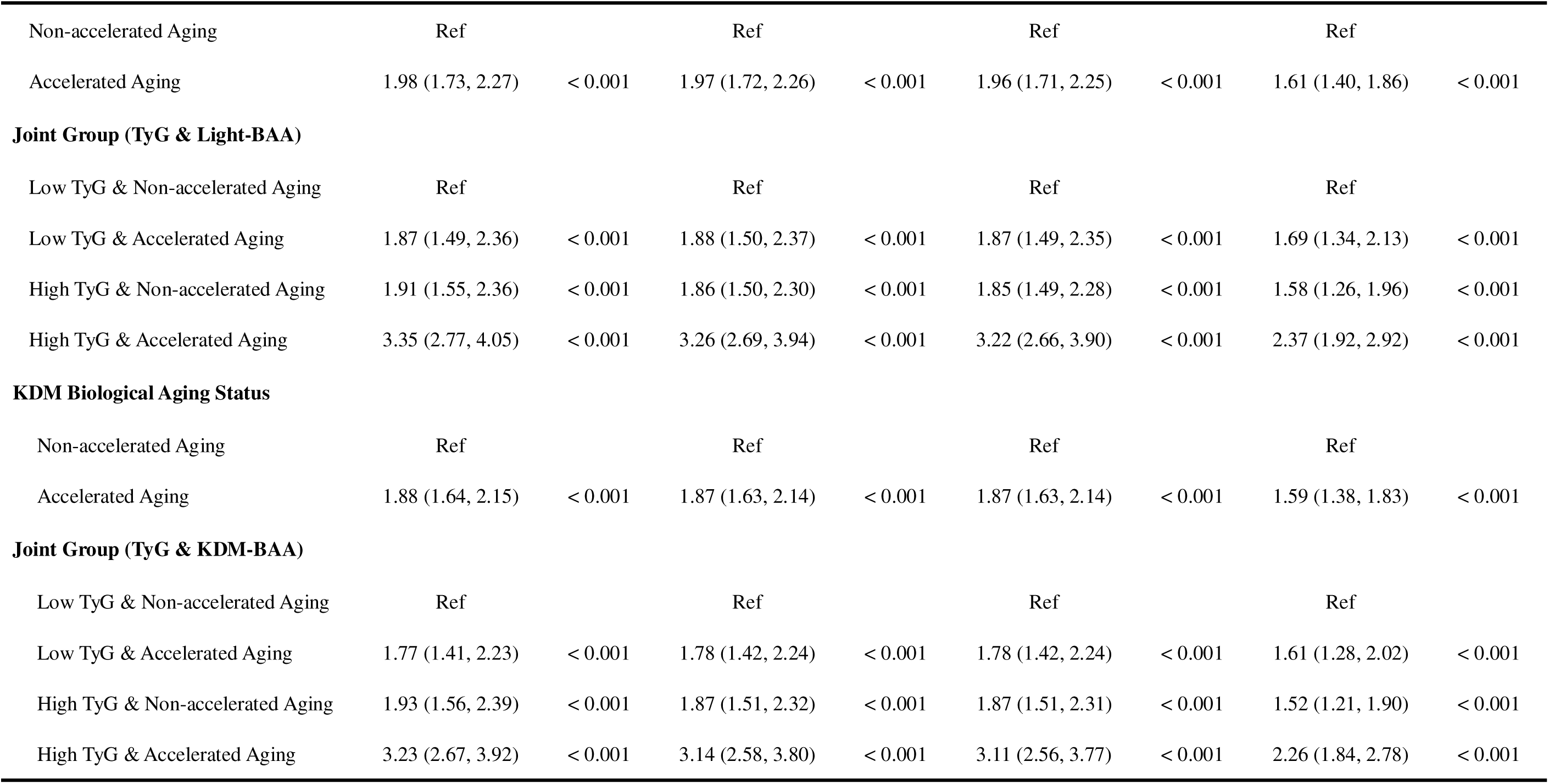
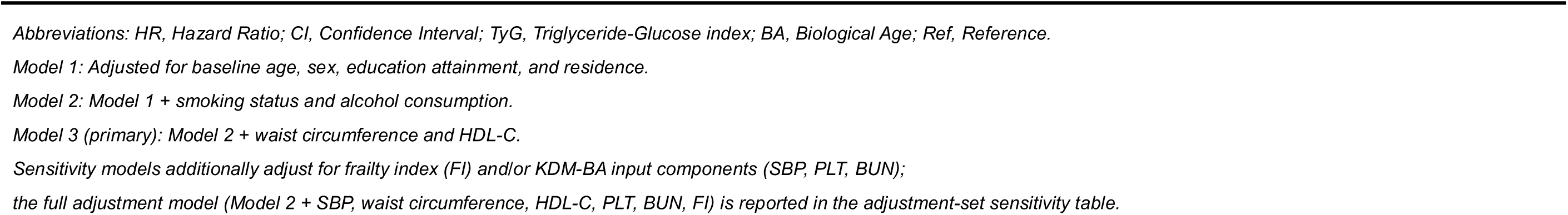
Joint associations of TyG index and biological age acceleration groups with incident cardiometabolic multimorbidity. HRs and 95% CIs from Cox models, with the “Low TyG & Non-accelerated Aging” group as reference. TyG cutoff: median (8.67); age acceleration: accelerated (>0 years). Results pooled across 5 multiply imputed datasets. Crude model: no covariates. Model 1: age, sex, education, residence. Model 2: Model 1 + smoking, alcohol. Model 3 (primary): Model 2 + waist circumference, HDL□C. *Abbreviations:* HR, hazard ratio; CI, confidence interval; TyG, triglyceride-glucose index; Light-BA, Light-method biological age; KDM-BA, Klemera–Doubal method biological age; BAA, biological age acceleration; CMM, cardiometabolic multimorbidity; Ref, reference; SBP, systolic blood pressure; PLT, platelet count; BUN, blood urea nitrogen; eGFR, estimated glomerular filtration rate.

| Characteristic | Crude Model |  | Model 1 |  | Model 2 |  | Model 3 |  |
| --- | --- | --- | --- | --- | --- | --- | --- | --- |
|  | HR (95% CI) | P value | HR (95% CI) | P value | HR (95% CI) | P value | HR (95% CI) | P value |
| TyG Status |  |  |  |  |  |  |  |  |
| Low TyG | Ref |  | Ref |  | Ref |  | Ref |  |
| High TyG | 2.03 (1.76, 2.34) | < 0.001 | 1.97 (1.71, 2.27) | < 0.001 | 1.95 (1.70, 2.25) | < 0.001 | 1.54 (1.32, 1.80) | < 0.001 |
| Light Biological Aging Status |  |  |  |  |  |  |  |  |
|  | HR (95% CI) | *P* value | HR (95% CI) | *P* value | HR (95% CI) | *P* value | HR (95% CI) | *P* value |
| Non-accelerated Aging | Ref |  | Ref |  | Ref |  | Ref |  |
| Accelerated Aging | 1.98 (1.73, 2.27) | < 0.001 | 1.97 (1.72, 2.26) | < 0.001 | 1.96 (1.71, 2.25) | < 0.001 | 1.61 (1.40, 1.86) | < 0.001 |
| **Joint Group (TyG & Light-BAA)** |  |  |  |  |  |  |  |  |
| Low TyG & Non-accelerated Aging | Ref |  | Ref |  | Ref |  | Ref |  |
| Low TyG & Accelerated Aging | 1.87 (1.49, 2.36) | < 0.001 | 1.88 (1.50, 2.37) | < 0.001 | 1.87 (1.49, 2.35) | < 0.001 | 1.69 (1.34, 2.13) | < 0.001 |
| High TyG & Non-accelerated Aging | 1.91 (1.55, 2.36) | < 0.001 | 1.86 (1.50, 2.30) | < 0.001 | 1.85 (1.49, 2.28) | < 0.001 | 1.58 (1.26, 1.96) | < 0.001 |
| High TyG & Accelerated Aging | 3.35 (2.77, 4.05) | < 0.001 | 3.26 (2.69, 3.94) | < 0.001 | 3.22 (2.66, 3.90) | < 0.001 | 2.37 (1.92, 2.92) | < 0.001 |
| **KDM Biological Aging Status** |  |  |  |  |  |  |  |  |
| Non-accelerated Aging | Ref |  | Ref |  | Ref |  | Ref |  |
| Accelerated Aging | 1.88 (1.64, 2.15) | < 0.001 | 1.87 (1.63, 2.14) | < 0.001 | 1.87 (1.63, 2.14) | < 0.001 | 1.59 (1.38, 1.83) | < 0.001 |
| **Joint Group (TyG & KDM-BAA)** |  |  |  |  |  |  |  |  |
| Low TyG & Non-accelerated Aging | Ref |  | Ref |  | Ref |  | Ref |  |
| Low TyG & Accelerated Aging | 1.77 (1.41, 2.23) | < 0.001 | 1.78 (1.42, 2.24) | < 0.001 | 1.78 (1.42, 2.24) | < 0.001 | 1.61 (1.28, 2.02) | < 0.001 |
| High TyG & Non-accelerated Aging | 1.93 (1.56, 2.39) | < 0.001 | 1.87 (1.51, 2.32) | < 0.001 | 1.87 (1.51, 2.31) | < 0.001 | 1.52 (1.21, 1.90) | < 0.001 |
| High TyG & Accelerated Aging | 3.23 (2.67, 3.92) | < 0.001 | 3.14 (2.58, 3.80) | < 0.001 | 3.11 (2.56, 3.77) | < 0.001 | 2.26 (1.84, 2.78) | < 0.001 |
|  | HR (95% CI) | P value | HR (95% CI) | P value | HR (95% CI) | P value | HR (95% CI) | P value |

Abbreviations: HR, Hazard Ratio; CI, Confidence Interval; TyG, Triglyceride-Glucose index; BA, Biological Age; Ref, Reference.

In the fully adjusted model, participants with elevated TyG and accelerated aging exhibited the highest risk of incident CMM, with an HR of 2.37 (95% CI, 1.92–2.92; *P* < 0.001) for the Light-BA definition and 2.26 (95% CI, 1.84–2.78; *P* < 0.001) for the KDM-BA definition. The intermediate joint groups also showed elevated risks compared with the reference group. The cumulative incidence curves diverged early and stayed distinct during the entire follow-up period for both biological-age algorithms (both log-rank *P* < 0.0001; Figure 2A and B).

**Figure 2.**
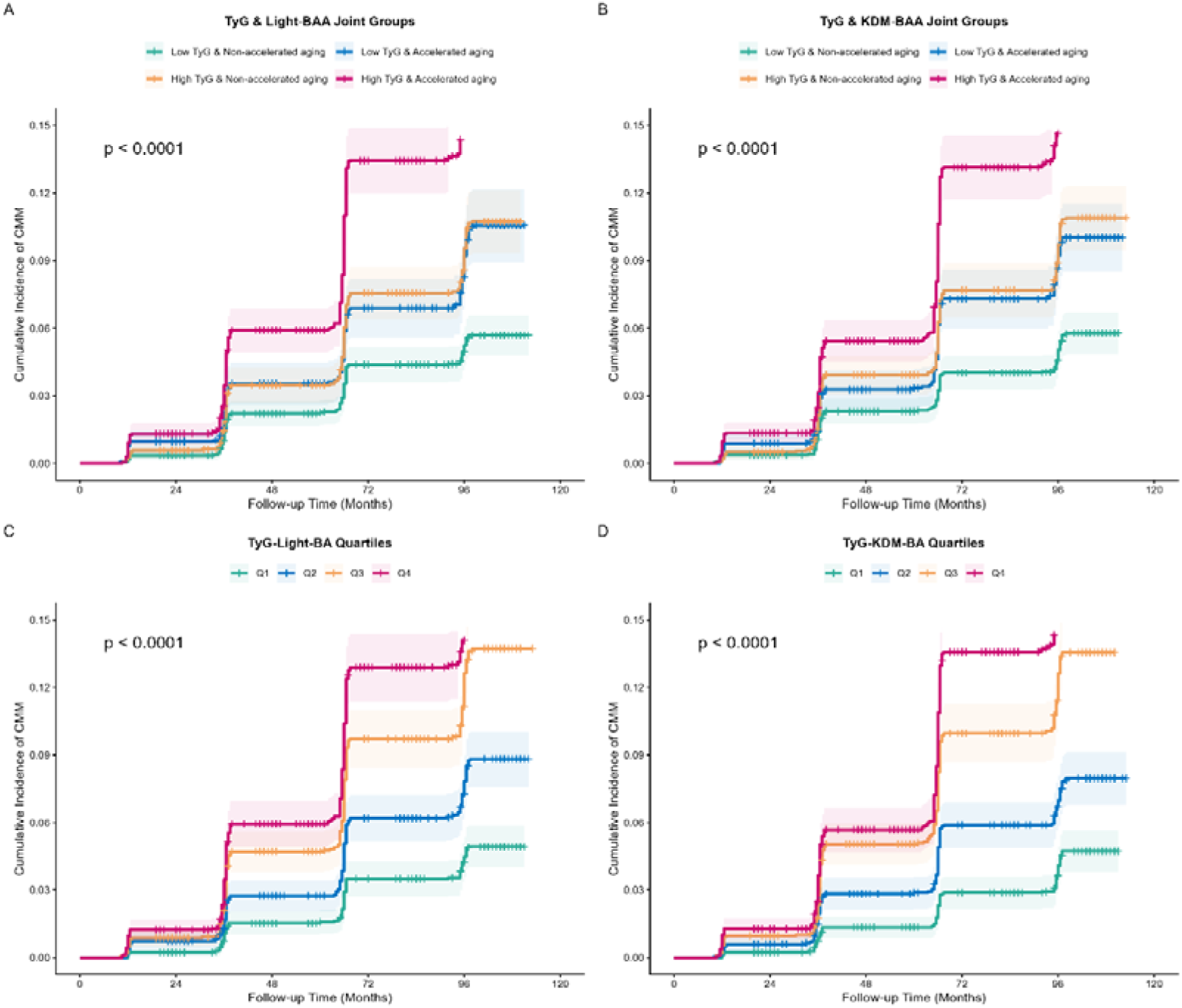
Cumulative incidence of CMM stratified by joint TyG and biological age acceleration (BAA) groups and by quartiles of composite TyG-BA indices. Cumulative incidence functions accounting for competing risks. (A) Four joint groups defined by TyG index and Light-BAA status; (B) four joint groups defined by TyG index and KDM-BAA status; (C) quartile strata of TyG-Light-BA; (D) quartile strata of TyG-KDM-BA. Shaded bands indicate 95% confidence intervals. Global log-rank *P* values are shown for between-group comparisons *Abbreviations:* TyG, triglyceride-glucose index; Light-BA, Light-method biological age; KDM-BA, Klemera-Doubal method biological age; CMM, composite cardiometabolic multimorbidity.

However, formal tests did not support a statistical interaction between TyG status and BAA on either the multiplicative or additive scale. The multiplicative interaction terms were not statistically significant for either Light-BA (HR, 0.89; 95% CI, 0.67–1.18; *P* = 0.418) or KDM-BA (HR, 0.93; 95% CI, 0.70–1.23; *P* = 0.593). Although RERI and AP estimates were positive in crude analyses, their CIs crossed the null after full adjustment (Supplementary Table 2). Therefore, the increased risk seen in the high TyG & accelerated aging group was considered a combined risk phenotype instead of proof of statistical interaction.

### 3.4 Continuous metabolic-aging burden and dose-response relationships

We next evaluated whether the combined metabolic-aging burden could be represented as a continuous measure using the multiplicative TyG × BA composites. Unlike the categorical joint analysis, which used BAA to identify accelerated aging relative to chronological age, the continuous composites used raw biological age to retain information on biological age level. In the fully adjusted Model 3, each 1-SD increase in TyG-Light-BA was associated with an HR of 1.62 (95% CI, 1.46–1.80; *P* < 0.001), while each 1-SD increase in TyG-KDM-BA was associated with an HR of 1.49 (95% CI, 1.38–1.61; *P* < 0.001) for incident CMM (Table 4).

**Table 4.**
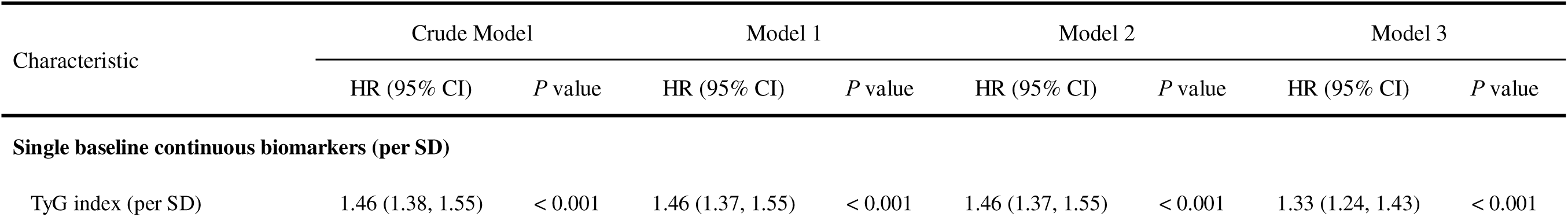
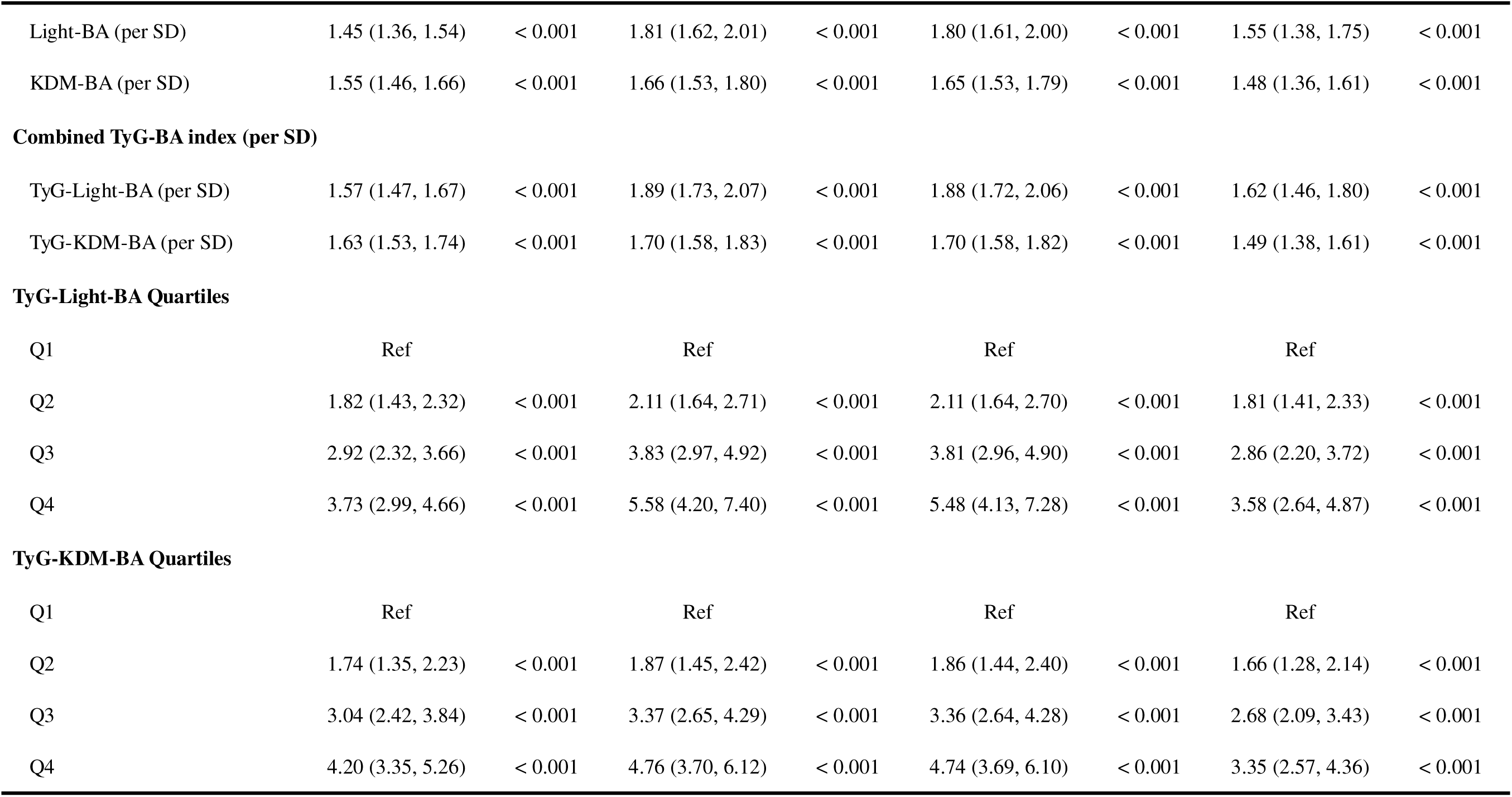
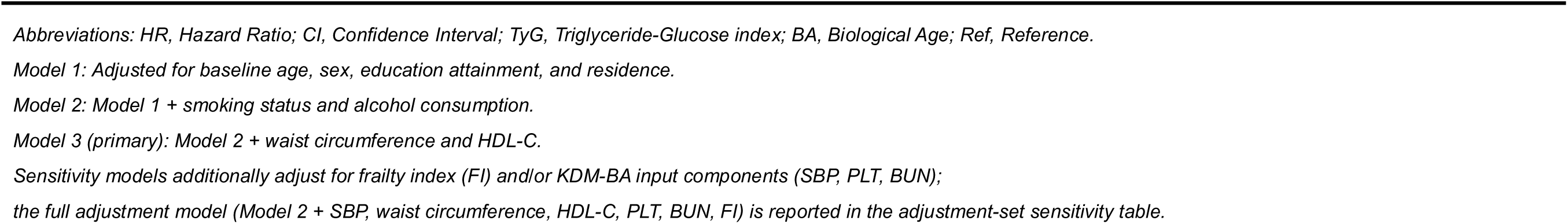
Associations of single and composite TyG–biological age indices with incident cardiometabolic multimorbidity. HRs and 95% CIs from Cox models. Single biomarkers and composite indices (TyG × biological age in years) were modeled per 1-SD increase; quartile analyses used Q1 as reference. Crude model: no covariates. Model 1: age, sex, education, residence. Model 2: Model 1 + smoking, alcohol. Model 3 (primary): Model 2 + waist circumference, HDL□ C. *Abbreviations:* HR, hazard ratio; CI, confidence interval; TyG, triglyceride-glucose index; Light-BA, Light-method biological age; KDM-BA, Klemera–Doubal method biological age; BA, biological age; Ref, reference; SD, standard deviation.

The results were consistent across quartile analyses.Compared with participants in the lowest quartile, those in the highest quartile had significantly greater risks of CMM for both TyG-Light-BA (HR, 3.58; 95% CI, 2.64–4.87) and TyG-KDM-BA (HR, 3.35; 95% CI, 2.57–4.36; both *P* < 0.001). Kaplan-Meier curves showed early and persistent separation across composite quartiles, with the highest quartile consistently exhibiting the greatest cumulative incidence (Figure 2C and D; both log-rank *P* < 0.0001).

RCS analyses provided more details on the dose-response relationships(Figure 3). The association of TyG with incident CMM was approximately linear (overall *P* < 0.001; nonlinear *P* = 0.288), whereas nonlinear associations were observed for Light-BA, KDM-BA, TyG-Light-BA, and TyG-KDM-BA (all overall and nonlinear *P* < 0.001). For Light-BA and KDM-BA, inflection points were estimated at 43.58 and 44.10 years, respectively. For the composite indices, the estimated thresholds were 3.77 for TyG-Light-BA/100 and 3.82 for TyG-KDM-BA/100, above which the hazard of CMM increased progressively.

**Figure 3.**
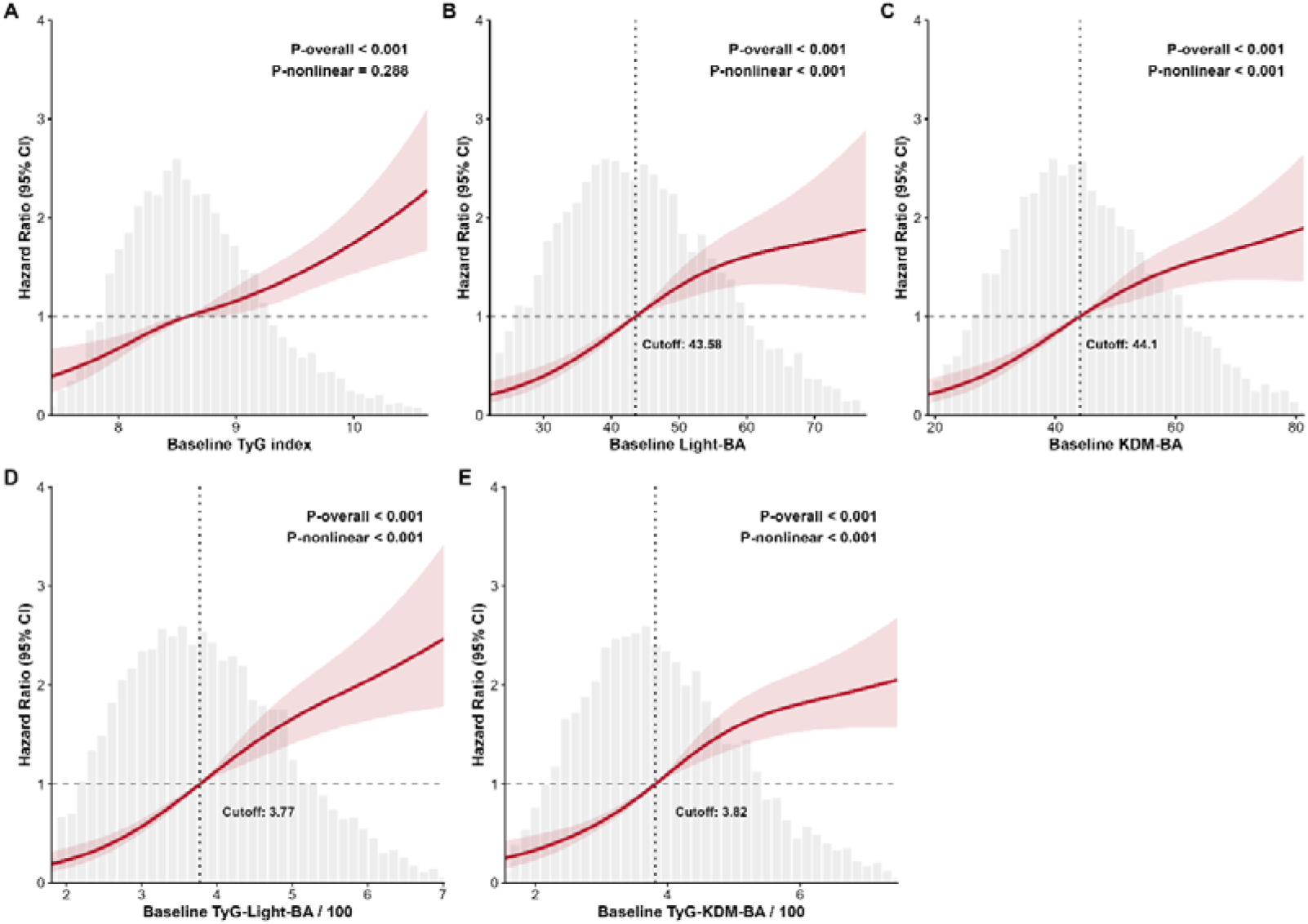
Nonlinear associations of baseline TyG index, biological aging metrics, and composite TyG-BA indices with incident CMM. Restricted cubic spline curves displaying adjusted hazard ratios (HRs) with 95% confidence intervals (shaded areas). The grey dashed line indicates HR = 1. Histograms show variable distributions; vertical dotted lines denote selected cutoff values. (A) Baseline TyG index; (B) baseline Light-BA; (C) baseline KDM-BA; (D) baseline TyG-Light-BA (/100); (E) baseline TyG-KDM-BA (/100). *P-overall* for global association; *P-nonlinear* for nonlinearity test. Models adjusted for age, sex, education, residence, smoking, alcohol use, waist circumference, and HDL-cholesterol. *Abbreviations:* TyG, triglyceride-glucose; Light-BA, Light biological age; KDM-BA, KDM biological age; CMM, cardiometabolic multimorbidity; HR, hazard ratio; CI, confidence interval.

### 3.5 Longitudinal metabolic-aging burden from 2011 to 2015

We next examined whether the association observed at baseline was also reflected in metabolic-aging burden accumulated across repeated measurements. Within the Wave 3 landmark cohort, 5,708 participants were followed for a median of 60 months after Wave 3, during which 432 incident CMM events occurred. Because only two exposure measurements were available, the cumulative index was interpreted as an approximation of average TyG-BA burden during 2011–2015 rather than a complete measure of lifelong cumulative exposure.

Each 1-SD increase in cumulative TyG-Light-BA was associated with an HR of 1.94 (95% CI, 1.66–2.25), while the corresponding HR for cumulative TyG-KDM-BA was 1.79 (95% CI, 1.60–2.00; Supplementary Table 3). Compared with the lowest quartile, participants in the highest quartile had markedly higher CMM risks for cumulative TyG-Light-BA (HR, 5.17; 95% CI, 3.40–7.87) and cumulative TyG-KDM-BA (HR, 4.75; 95% CI, 3.31–6.82; both *P* < 0.001). Cumulative incidence curves showed consistent separation across cumulative-exposure quartiles (Supplementary Figure 2A and B; both log-rank *P* < 0.0001). RCS analyses similarly showed progressively increasing CMM risk at higher cumulative exposure levels (Supplementary Figure 3). These findings suggested that a higher average metabolic-aging burden across 2011-2015 was associated with subsequent CMM risk within the Wave 3 landmark framework, although the limited number of repeated measurements precluded interpretation as a complete long-term cumulative exposure.

### 3.6 Two-wave trajectory patterns of TyG-BA

To further characterize whether consistently higher metabolic-aging burden across the two available measurements was associated with subsequent CMM, K-means clustering was applied to TyG-BA composite values measured at Waves 1 and 3. For TyG-Light-BA, 37.3% of participants were classified into the low-level group, 43.8% into the moderate-level group, and 18.9% into the high-level group. The corresponding proportions for TyG-KDM-BA were 36.7%, 43.4%, and 19.8%, respectively (Figure 4).

**Figure 4.**
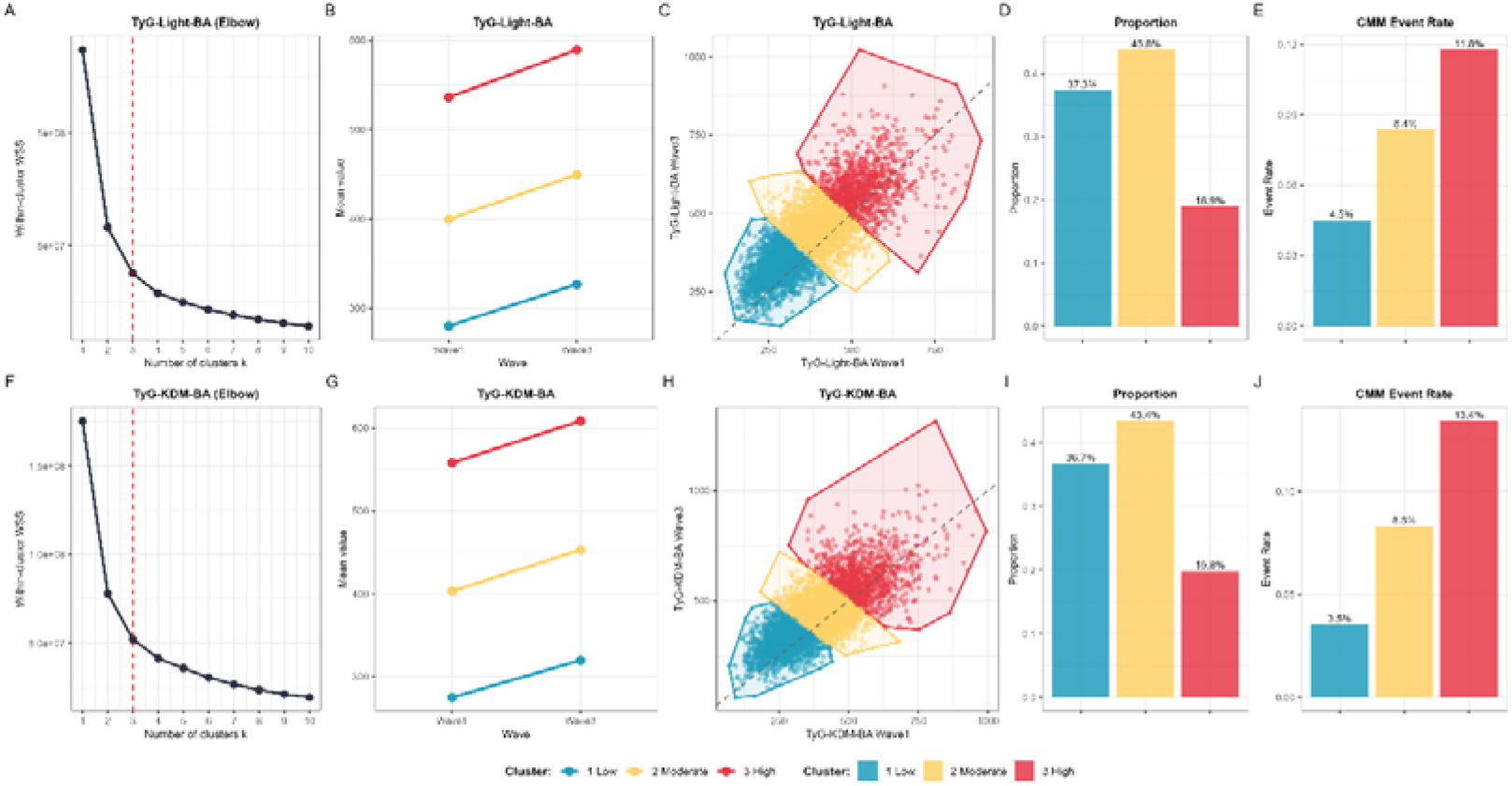
Longitudinal trajectory clustering of TyG-Light-BA and TyG-KDM-BA and incidence of CMM. (A, F) Elbow plots of within-cluster sum of squares (WSS) for K-means clustering, with vertical red dashed lines marking the optimal number of clusters (k = 3). (B, G) Mean longitudinal trajectories of the three clusters across Waves 1–3. (C, H) Scatter plots of Wave 1 versus Wave 3 biomarker values by cluster; grey dashed line represents the identity line. (D, I) Distribution proportions of participants in each trajectory cluster. (E, J) Crude CMM event rates stratified by trajectory cluster. *Abbreviations:* TyG, triglyceride-glucose index; Light-BA, Light biological age; KDM-BA, KDM biological age; CMM, cardiometabolic multimorbidity; WSS, within-cluster sum of squares.

Compared with the low-level group, participants in the high-level group had substantially higher risks of incident CMM in Model 3, with HRs of 3.93 (95% CI, 2.65–5.82) for TyG-Light-BA and 4.51 (95% CI, 3.23–6.30) for TyG-KDM-BA (both *P* < 0.001; Supplementary Table 3). Cluster stability remained high across bootstrap resampling. Cumulative incidence curves separated early and remained distinct throughout follow-up (Supplementary Figure 2C and D; both log-rank *P* < 0.0001). Because only two measurement points were available, these clusters were interpreted as two-wave exposure-level patterns rather than persistent biological trajectories.

### 3.7 Associations of TyG-BA composites across the cardiometabolic disease continuum

We further investigated whether the associations of the TyG-BA composites persisted across the cardiometabolic disease continuum using a full illness-death multi-state model (Table 5 and Figure 5). The model incorporated five transitions: from a cardiometabolic-disease-free state to incident cardiometabolic disease (n = 2,167), from a cardiometabolic-disease-free state to death (n = 476), from incident cardiometabolic disease to cardiometabolic multimorbidity (CMM) (n=711), from incident cardiometabolic disease to death (n = 431), and from CMM to death (n = 69).

**Figure 5.**
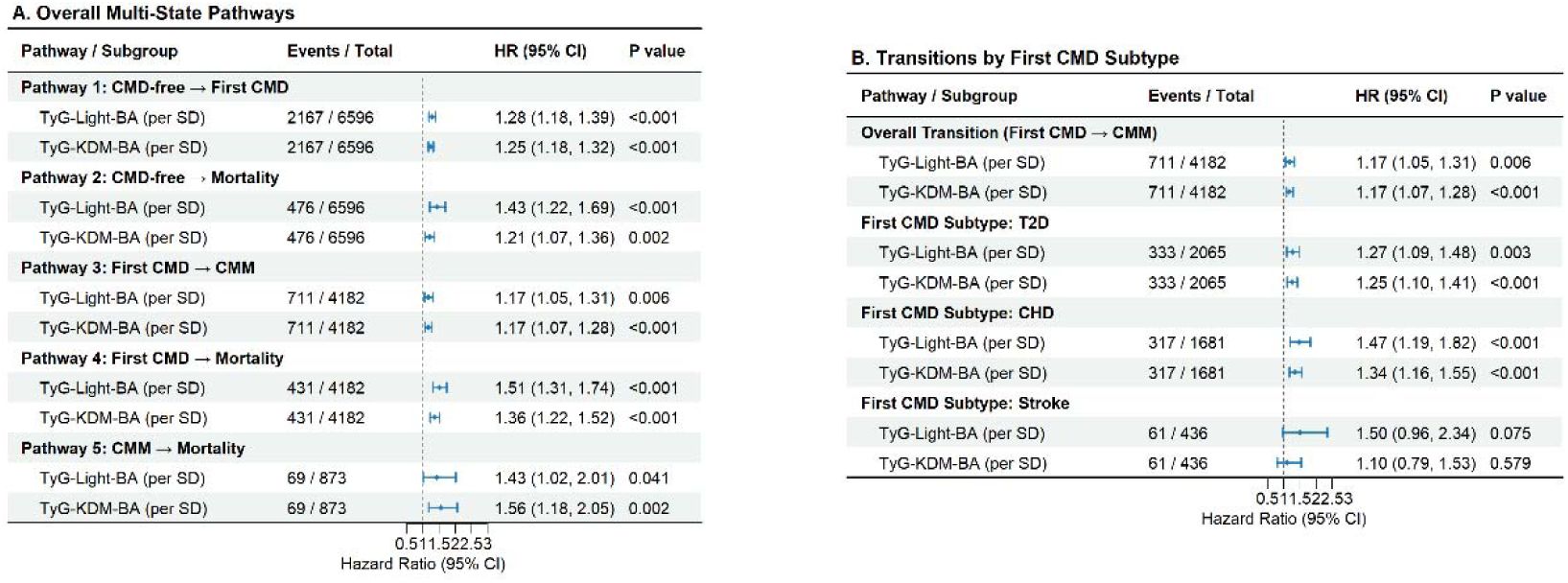
Multi-state Cox regression analyses of TyG-BA composite indices for the progression of CMM and mortality. (A) Hazard ratios (HRs) and 95% confidence intervals (CIs) are shown for TyG-Light-BA and TyG-KDM-BA (per standard deviation increase) across five pre-specified transition pathways in the overall multi-state model. (B) The transition from first CMD to CMM stratified by the subtype of the first cardiometabolic disease. *Abbreviations:* TyG, triglyceride-glucose index; Light-BA, Light biological age; KDM-BA, KDM biological age; CMD, cardiometabolic disease; CMM, cardiometabolic multimorbidity; T2D, type 2 diabetes; CHD, coronary heart disease; HR, hazard ratio; CI, confidence interval; SD, standard deviation.

**Table 5.**
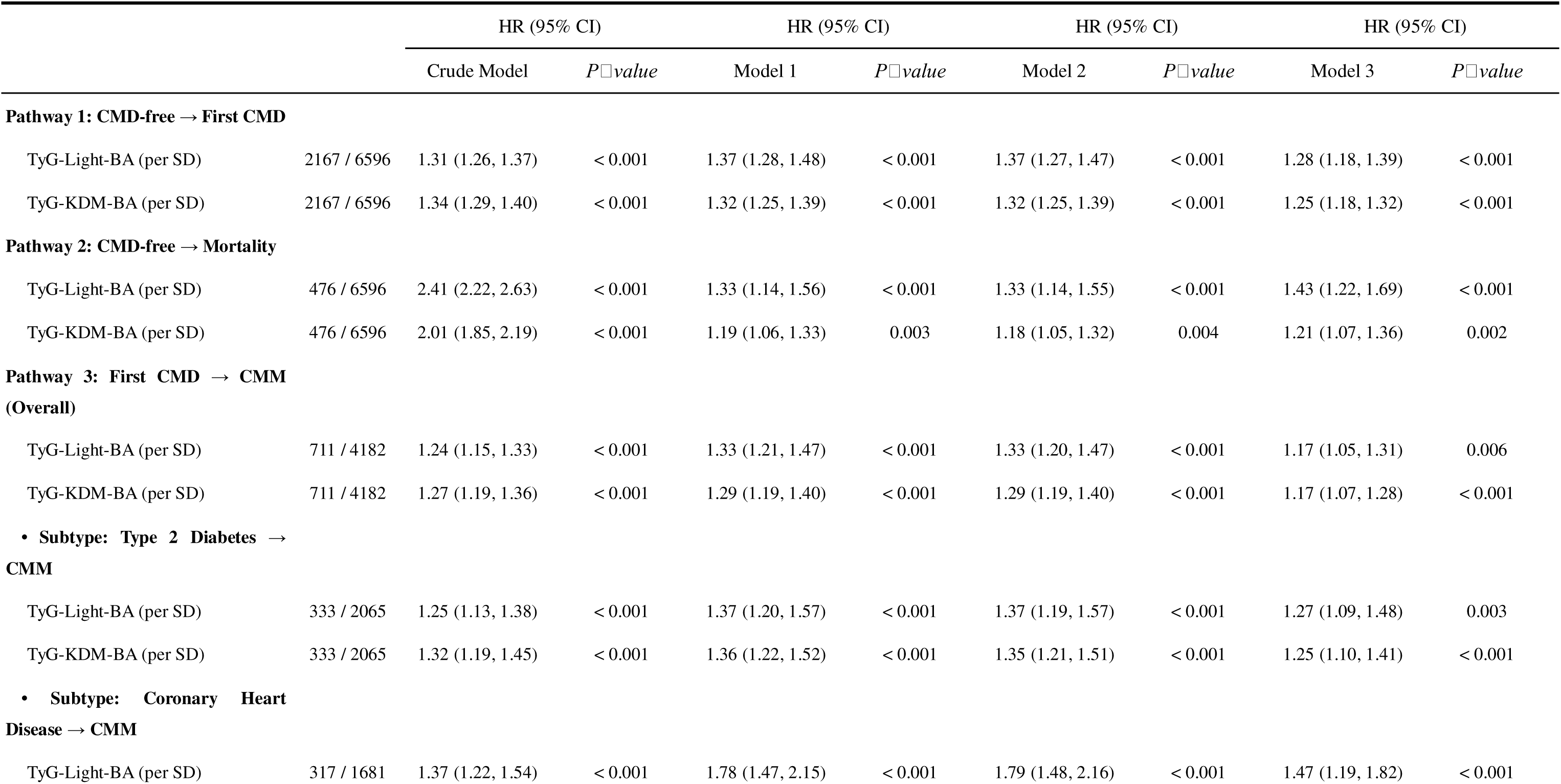
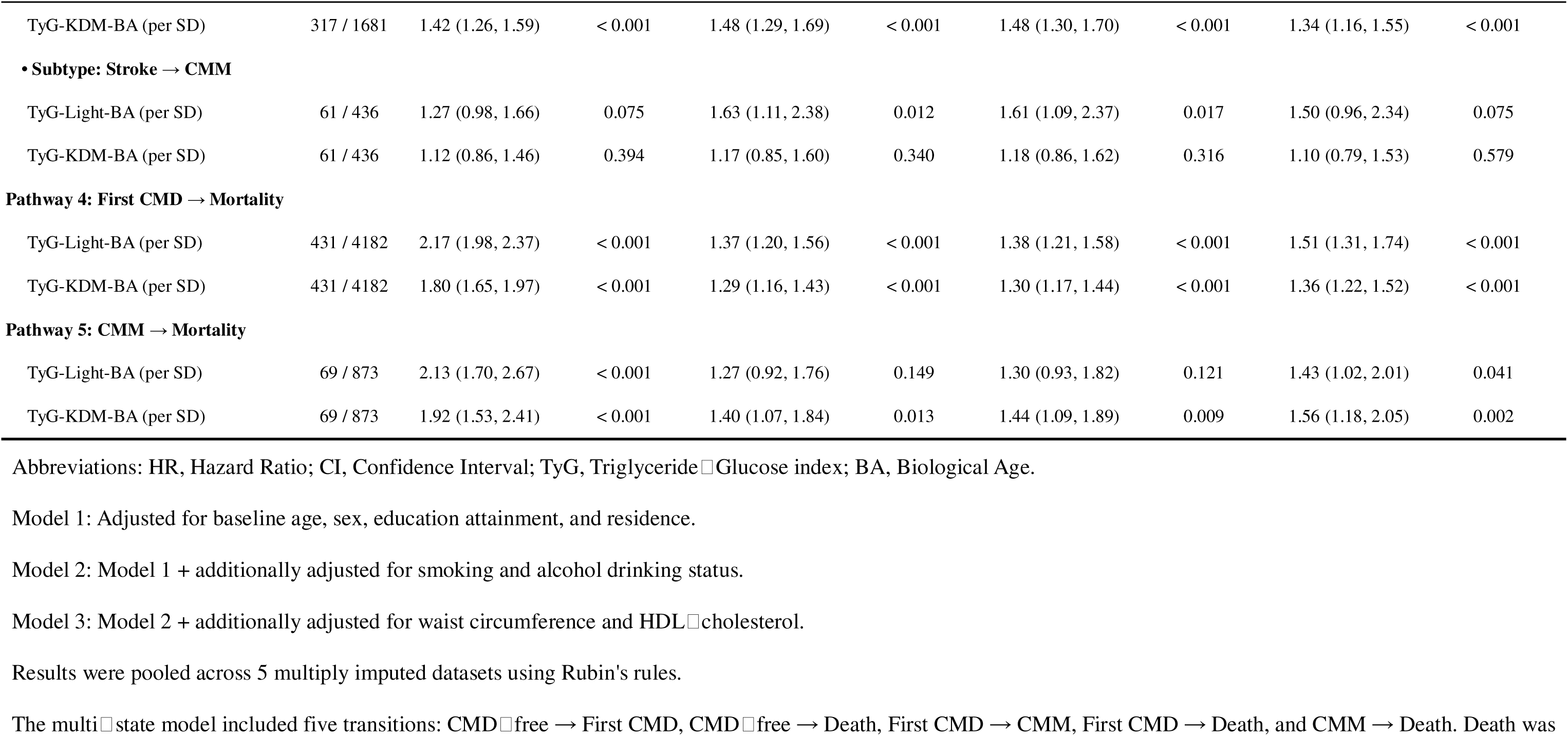
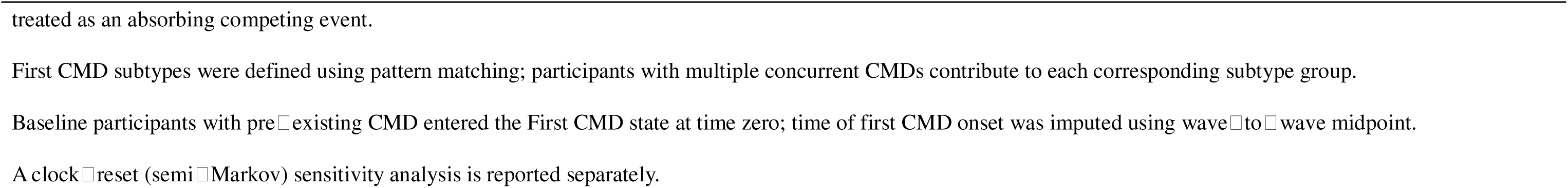
Multi-state Cox regression of TyG-BA composite indices for progression from CMD-free to first CMD, CMM, and mortality. HRs and 95% CIs per 1-SD increase in composite indices, estimated from a multi-state Cox model with five transitions: CMD-free → first CMD, CMD-free → death, first CMD → CMM, first CMD → death, and CMM → death (death treated as an absorbing competing event). First CMD subtypes (T2D, CHD, stroke) were defined by pattern matching; participants with multiple concurrent CMDs contributed to each corresponding subtype group. Participants with pre-existing CMD at baseline entered the first CMD state at time zero; time of first CMD onset was imputed using wave-to-wave midpoint. Models adjusted as described in Table 2. Results pooled across 5 multiply imputed datasets. *Abbreviations:* HR, hazard ratio; CI, confidence interval; TyG, triglyceride-glucose index; Light-BA, Light biological age; KDM-BA, Klemera–Doubal biological age; CMD, cardiometabolic disease; CMM, cardiometabolic multimorbidity; T2D, type 2 diabetes; CHD, coronary heart disease.

Higher TyG-BA values were associated with increased hazards across multiple transitions. For the transition from a cardiometabolic-disease-free state to incident cardiometabolic disease, the HRs per 1-standard-deviation increment were 1.28 (95% CI, 1.18–1.39) for TyG-Light-BA and 1.25 (95% CI, 1.18–1.32) for TyG-KDM-BA, with both *P* values < 0.001. Among individuals who developed incident cardiometabolic disease, both composite indices were associated with subsequent CMM, with HRs of 1.17 (95% CI, 1.05–1.31) for TyG-Light-BA and 1.17 (95% CI, 1.07–1.28) for TyG-KDM-BA. The two composites were also associated with death before CMM, with HRs of 1.51 (95% CI, 1.31–1.74) and 1.36 (95% CI, 1.22–1.52), respectively; all corresponding *P* values were ≤ 0.006. Higher TyG-BA was additionally associated with mortality from the cardiometabolic-disease-free state, with HRs of 1.43 (95% CI, 1.22–1.69) for TyG-Light-BA and 1.21 (95% CI, 1.07–1.36) for TyG-KDM-BA, and with mortality among participants with CMM, with corresponding HRs of 1.43 (95% CI, 1.02–2.01) and 1.56 (95% CI, 1.18–2.05), respectively.

Subtype-specific analyses of progression from incident cardiometabolic disease to CMM revealed the strongest associations among participants whose initial cardiometabolic disease was coronary heart disease, with HRs of 1.47 (95% CI, 1.19–1.82) for TyG-Light-BA and 1.34 (95% CI, 1.16–1.55) for TyG-KDM-BA. Significant associations were also observed when type 2 diabetes was the initial disease, with corresponding HRs of 1.27 (95% CI, 1.09–1.48) and 1.25 (95% CI, 1.10–1.41), respectively. In contrast, associations were weaker and not statistically significant when stroke was the initial disease, with HRs of 1.50 (95% CI, 0.96–2.34; *P* = 0.075) for TyG-Light-BA and 1.10 (95% CI, 0.79–1.53; *P* = 0.579) for TyG-KDM-BA.

The Markov assumption was not supported for the transitions from incident cardiometabolic disease to CMM and from CMM to death, whereas no evidence of violation was observed for the transition from incident cardiometabolic disease to death (Supplementary Table 4). Clock-reset semi-Markov sensitivity analyses yielded comparable associations for the transition from incident cardiometabolic disease to CMM, with HRs of 1.15 (95% CI, 1.03–1.29) for TyG-Light-BA and 1.16 (95% CI, 1.07–1.27) for TyG-KDM-BA, and for the transition from CMM to death, with corresponding HRs of 1.43 (95% CI, 1.03–1.99) and 1.47 (95% CI, 1.12–1.93), respectively. Effect estimates for the remaining transitions were directionally consistent with the primary analysis (Supplementary Table 5).

### 3.8 Exploratory Subgroup and Heterogeneity Analyses

Associations between the TyG-BA composites and incident CMM remained directionally consistent across the evaluated subgroups (Figure 7; Supplementary Tables 6 and 7). Significant heterogeneity by age was observed for both composites (both *P* for interaction <0.001). For TyG-Light-BA, the association was stronger among participants aged <65 years (HR, 1.74; 95% CI, 1.57–1.92) than among those aged ≥65 years (HR, 1.23; 95% CI, 1.04–1.47). A similar pattern was observed for TyG-KDM-BA, with HRs of 1.64 (95% CI, 1.51–1.79) and 1.25 (95% CI, 1.09–1.44), respectively.

Hypertension also modified the associations for both composites (*P* for interaction = 0.017 for TyG-Light-BA and <0.001 for TyG-KDM-BA), with stronger associations observed among participants without hypertension. Interaction by hyperlipidemia was marginal for TyG-KDM-BA (*P* = 0.054) but not evident for TyG-Light-BA (*P* = 0.305). CKM stage showed evidence of heterogeneity for both composites (*P* for interaction = 0.019 and 0.034, respectively), with attenuated associations among participants with stage 4 disease. There were no statistically significant interactions found for sex, BMI, residence, education, smoking, or alcohol consumption (all *P* for interaction >0.05).

Frailty-stratified Fine-Gray models showed positive associations across all three FI categories, although effect estimates decreased with increasing frailty (Supplementary Tables 8). For TyG-Light-BA, sHRs decreased from 1.93 (95% CI, 1.49–2.49) among robust participants to 1.54 (95% CI, 1.32–1.78) among pre-frail and 1.41 (95% CI, 1.18–1.68) among frail participants. A similar gradient was observed for TyG-KDM-BA, with sHRs of 1.71 (95% CI, 1.41–2.07), 1.43 (95% CI, 1.28–1.59), and 1.34 (95% CI, 1.17–1.55), respectively (all *P* < 0.001; *P* for interaction <0.001). Given the exploratory nature and conceptual overlap between FI and several cardiometabolic conditions, these findings were interpreted as evidence of heterogeneity rather than causal effect modification.

### 3.9 Incremental predictive performance and comparison with alternative integration strategies

We next evaluated whether the TyG-BA composites improved risk discrimination beyond the prespecified clinical model. At 96 months, adding TyG-Light-BA and TyG-KDM-BA increased the IPCW-adjusted C-index by 0.015 and 0.024, respectively, compared with the base model, with both improvements reaching statistical significance. The corresponding optimism-corrected C-indices were 0.671 and 0.677, compared with 0.656 for the base model. The continuous IPCW-weighted NRI and IDI further confirmed the additional predictive value of both composites(Table 6). Decision curve analysis showed that models including the TyG-BA composites provided a higher net clinical benefit over a wide range of threshold probabilities(Figure 6).

**Figure 6.**
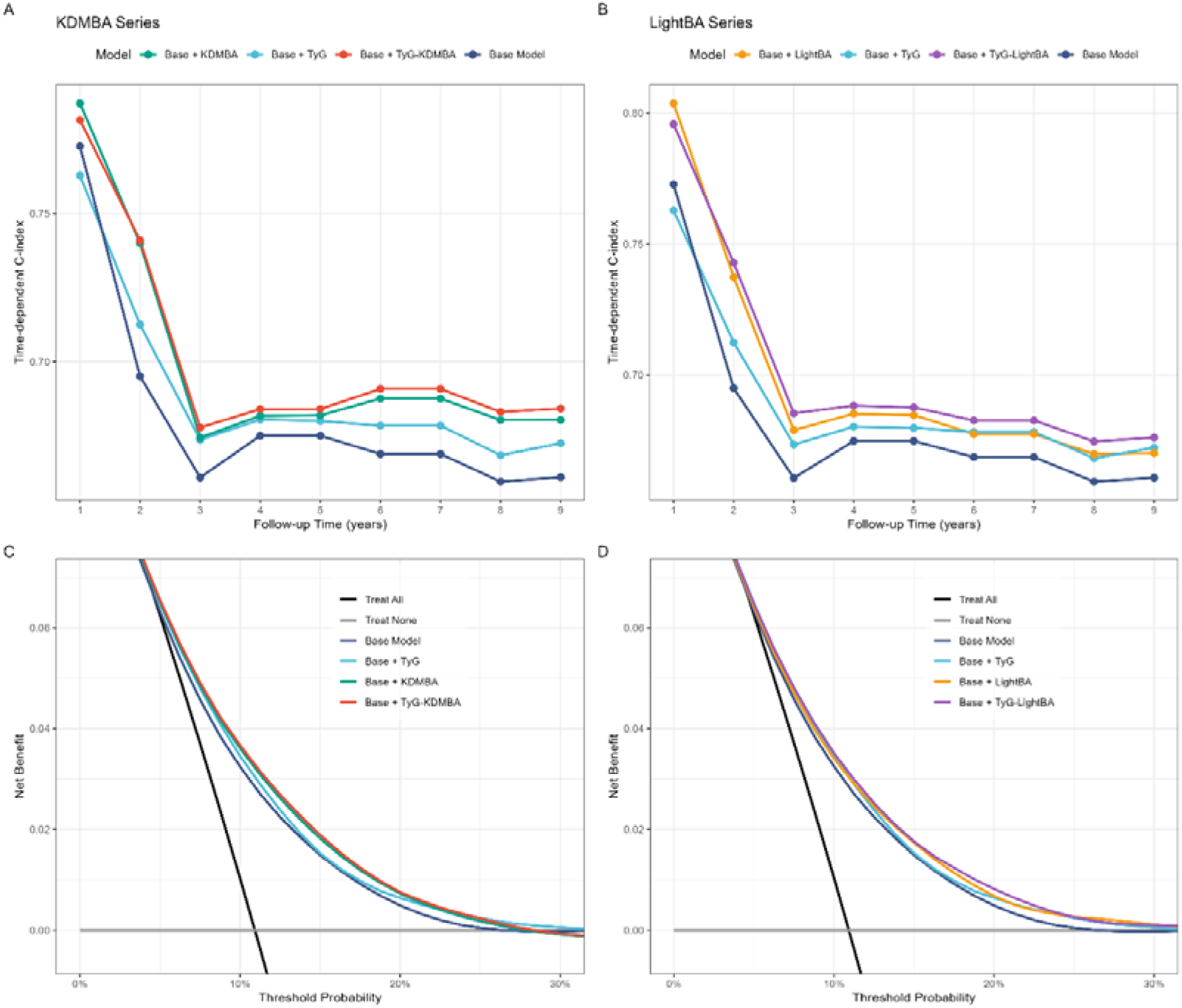
Incremental predictive performance of metabolic-aging composite indices. (A) Time-dependent C-index curves for models adding KDM-BA and TyG-KDM-BA compared with the base model and base model plus TyG. (B) Corresponding curves for Light-BA and TyG-Light-BA. (C, D) Decision curve analysis at 108 months showing net benefit across risk thresholds for models with KDM-BA and TyG–KDM-BA (C) and with Light-BA and TyG–Light-BA (D). *Abbreviations:* TyG, triglyceride-glucose index; KDM-BA, Klemara-Doubal method biological age; Light-BA, Light method biological age.

**Figure 7.**
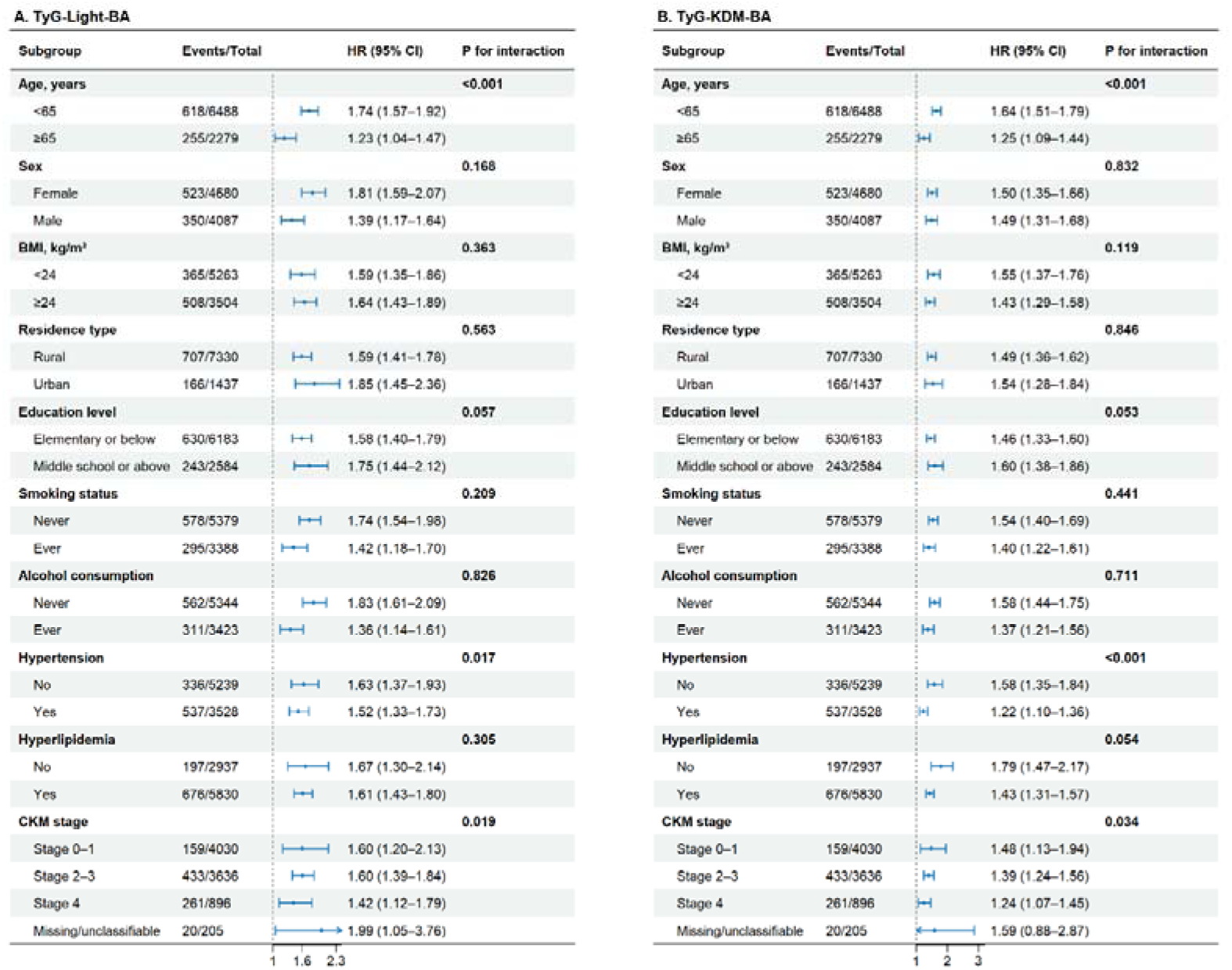
Subgroup analyses of TyG-Light-BA and TyG-KDM-BA for the incident CMM. Forest plots displaying HRs (95% CIs) per SD increase in each composite index. (A) TyG-Light-BA; (B) TyG-KDM-BA. *P* for interaction tests heterogeneity across subgroups. *Abbreviations:* TyG, triglyceride-glucose index; Light-BA, Light biological age; KDM-BA, KDM biological age; CMD, cardiometabolic disease; CMM, cardiometabolic multimorbidity; BMI, body mass index; CKM, cardiometabolic multimorbidity stage; HR, hazard ratio; CI, confidence interval.

**Table 6.**
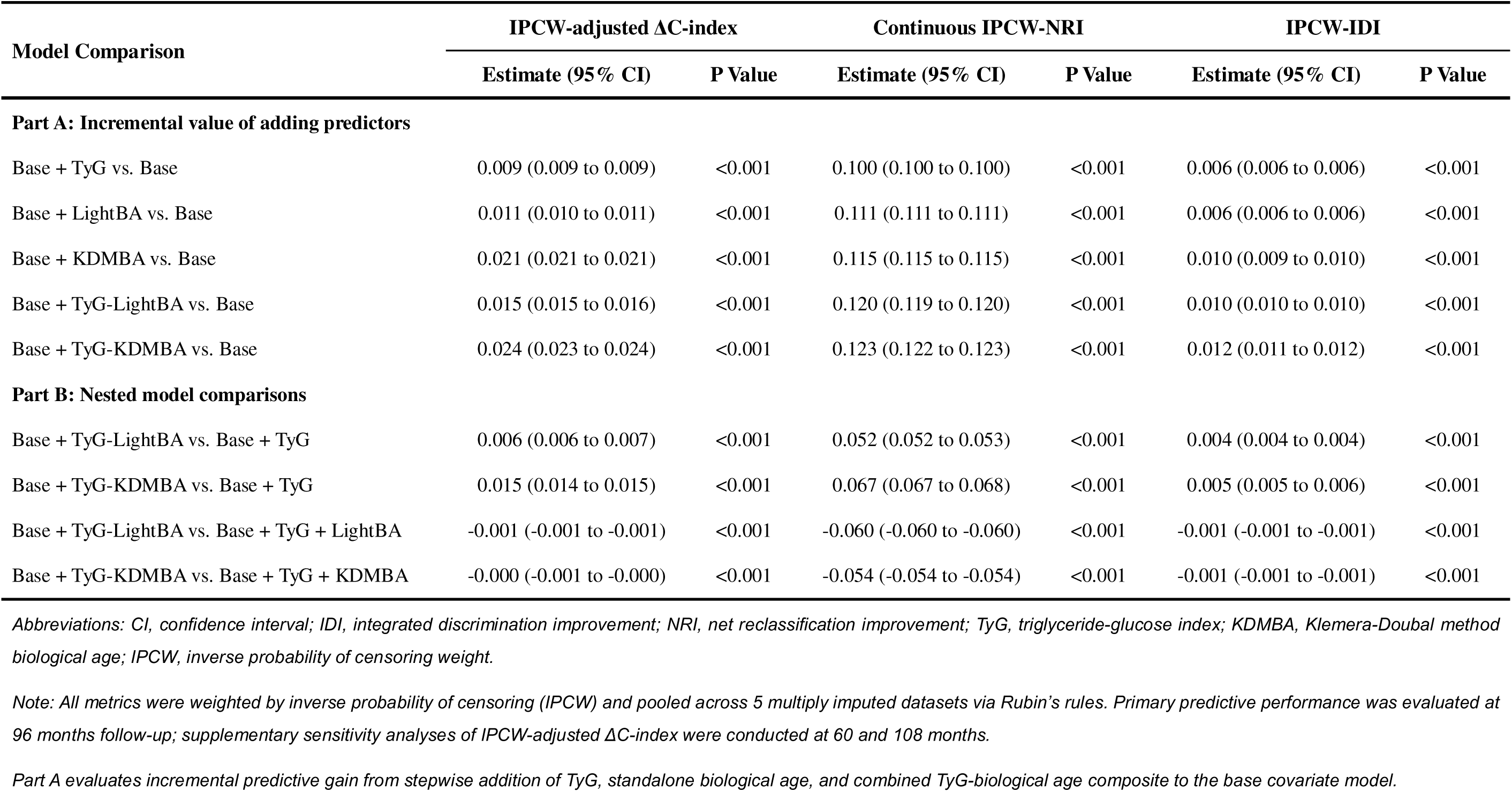
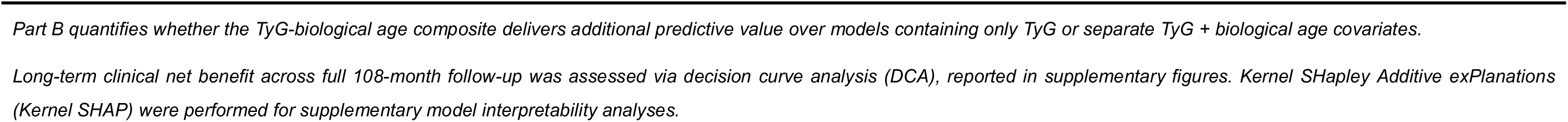
Incremental predictive performance of adding TyG index, biological age, and composite TyG-BA indices to the base model. All metrics were IPCW-adjusted and pooled across 5 multiply imputed datasets using Rubin’s rules. Part A: incremental gain from stepwise addition of each predictor to the base model (age, sex, education, residence, smoking, alcohol, waist circumference, HDL-C). Part B: nested model comparisons evaluating whether composite indices add value beyond TyG alone or TyG + biological age separately. Primary evaluation at 96 months; sensitivity analyses at 60 and 108 months are reported in supplementary table 10. *Abbreviations:* CI, confidence interval; IDI, integrated discrimination improvement; NRI, net reclassification improvement; IPCW, inverse probability of censoring weighting; TyG, triglyceride-glucose index; Light-BA, Light biological age; KDM-BA, Klemera–Doubal biological age.

Nested comparisons further clarified the role of the composites. Compared with the TyG-only model, TyG-KDM-BA provided a significant improvement in discrimination, whereas the improvement associated with TyG-Light-BA was not statistically significant. In contrast, neither composite improved discrimination beyond models incorporating TyG and the corresponding biological age measure as separate covariates. Consistently, models including TyG and biological age as separate predictors showed slightly better overall model fit than the corresponding multiplicative composite models (Table 6). These findings indicate that the TyG-BA composites primarily provide a parsimonious summary of concurrent metabolic and biological-aging burden rather than a statistically superior alternative to modeling TyG and biological age separately.

Time-dependent ROC analyses further assessed discrimination across different prediction horizons. At 3 and 5 years, TyG-Light-BA yielded the highest AUCs among the evaluated markers, at 0.689 and 0.697, respectively, compared with 0.676 and 0.688 for TyG. At 8 years, TyG-KDM-BA showed the highest AUC at 0.699, compared with 0.683 for TyG. However, after Benjamini–Hochberg adjustment for multiple comparisons, only the difference between TyG-KDM-BA and TyG at 8 years remained statistically significant (adjusted *P* = 0.007), whereas the corresponding comparisons at 3 and 5 years were not significant (Supplementary Table 9; Supplementary Figure 4).

The incremental discrimination of the composites remained statistically significant at both 60 and 108 months, supporting the robustness of the primary findings across alternative follow-up horizons (Supplementary Table 10). Exploratory Kernel SHAP analyses similarly identified the TyG-BA composites as important predictors within the corresponding models (Supplementary Figure 5).

### 3.10 Competing-risk and other sensitivity analyses

Fine-Gray models, which consider death as a competing event, produced estimates similar to those from the main Cox analyses. The subdistribution HRs were 1.56 (95% CI, 1.41–1.73) for TyG-Light-BA and 1.45 (95% CI, 1.34–1.56) for TyG-KDM-BA. Joint-group, cumulative-exposure, and two-wave exposure-level analyses similarly remained consistent in competing-risk models (Supplementary Tables 11–13).

The associations were also robust across multiple analytical specifications, including exclusion of early incident CMM events, complete-case analyses, exclusion of participants with baseline CKD or hypertension, exclusion of baseline medication users, additional adjustment for medication use, replacement of waist circumference with BMI, omission of the HbA1c criterion from the CMM definition, and additional adjustment for UA or eGFR. When follow-up was truncated at Wave 3, effect estimates were moderately attenuated but remained statistically significant (TyG-Light-BA: HR, 1.50; 95% CI, 1.26–1.78; TyG-KDM-BA: HR, 1.26; 95% CI, 1.10–1.44; Supplementary Table 14). Interval-censored Weibull models similarly supported shorter CMM-free survival with increasing composite exposure (time ratio, 0.74; 95% CI, 0.69–0.79 for TyG-Light-BA and 0.78; 95% CI, 0.74–0.82 for TyG-KDM-BA; Supplementary Table 15).

Sensitivity analyses focusing specifically on chronological-age adjustment showed that the continuous composite associations were not materially dependent on inclusion of chronological age in the covariate set. For TyG-Light-BA, the HR per SD was 1.46 without age adjustment versus 1.62 in the age-adjusted model; for TyG-KDM-BA, the corresponding estimates were 1.49 and 1.49, respectively. Quartile-based analyses showed the same overall pattern (Supplementary Table 16).

Additional covariate adjustment had little influence on TyG-Light-BA, with a maximum relative change in the HR of 4.7%. For TyG-KDM-BA, adjustment for SBP, PLT, and BUN increased the HR from 1.49 to 1.86. Because SBP and BUN are components of the KDM-BA algorithm itself, this increase was considered compatible with overadjustment rather than evidence against the primary association; therefore, the prespecified primary model was retained for main inference (Supplementary Figure 6).

## 4. Discussion

Metabolic dysfunction and biological aging represent two complementary but biologically distinct dimensions of cardiometabolic vulnerability. The TyG index primarily captures insulin resistance and metabolic dysregulation, which are implicated in hyperglycemia[29], dyslipidemia[30], endothelial dysfunction[31], and systemic inflammation [32]. Chronic metabolic stress contributes to vascular injury, atherogenesis, and the development of individual cardiometabolic diseases, including T2D and coronary heart disease[33] [34]. In parallel, biological aging reflects cumulative physiological deterioration across multiple organ systems, encompassing cellular senescence [35], mitochondrial dysfunction[36], impaired proteostasis [37], and the chronic low-grade inflammation that characterizes aging-related immune dysregulation [38]. These age-related changes may reduce physiological reserve and resilience, increasing susceptibility to additional metabolic and vascular insults. Although metabolic dysfunction and biological aging are biologically interconnected, they capture distinct aspects of cardiometabolic vulnerability. Metabolic dysfunction reflects an active metabolic burden, whereas biological aging reflects the cumulative loss of physiological reserve. Thus, neither dimension alone may fully characterize the systemic vulnerability underlying cardiometabolic multimorbidity. Previous studies have largely examined metabolic dysfunction and biological aging separately, leaving their combined relevance to the development and progression of CMM insufficiently characterized. Accordingly, we evaluated whether combining the two dimensions better characterizes cardiometabolic vulnerability, examining their independent and joint associations with CMM, the dose-response and longitudinal patterns of the composite burden, associations with progression across the cardiometabolic disease continuum, and incremental predictive value.

In mutually adjusted Cox models, TyG and biological age were independently associated with incident CMM, each contributing risk beyond the other dimension. Each 1-SD increase in TyG was associated with a 23–27% higher risk when adjusted for biological age, while each 1-SD increase in biological age was associated with a 39–44% higher risk when adjusted for TyG. These results back the perspective that metabolic dysfunction and biological aging capture partially distinct but complementary dimensions of CMM risk. For joint classification, we used two complementary biological-age metrics: BAA (the residual) to identify individuals whose biological age exceeds chronological expectations [19], and raw biological age to quantify cumulative physiological burden[39]. Through categorical joint classification, participants with concurrent high TyG and accelerated biological aging had the highest risk of CMM, with HRs of 2.37 and 2.26 for the Light-BA and KDM-BA definitions, respectively. Although formal interaction analyses did not provide evidence of multiplicative or additive interaction, the substantially higher risk observed among individuals with both elevated TyG and accelerated biological aging indicates that their co-occurrence may still be clinically informative as a joint risk phenotype.

The continuous TyG-BA composite indices further quantified the overall burden on a continuous scale, showing that each 1-SD increase in TyG-Light-BA and TyG-KDM-BA was associated with 62% and 49% higher risks of incident CMM, respectively, with the highest quartile showing more than threefold increased risks. RCS analyses showed nonlinear dose-response relationships, with inflection points above which the hazard of CMM rose progressively; this finding has not been reported previously and may inform threshold-based screening. The continuous composites provided a convenient way to summarize the concurrent burden of metabolic dysfunction and biological aging on a single scale. Their stronger associations relative to the individual components should be interpreted cautiously, as effect-size differences across differently constructed predictors do not by themselves establish superior predictive information.

One baseline measurement might not completely reflect a person’s long-term exposure history, as metabolic and aging-related biomarkers can fluctuate over time due to lifestyle changes[40], disease progression [41], or therapeutic interventions [42]. To address this, we conducted cumulative exposure and two-wave trajectory analyses using repeated measurements from 2011 and 2015 within a Wave 3 landmark framework. With only two measurement time points available, these analyses provide an approximation of the average burden over 2011–2015 rather than a complete measure of lifelong cumulative exposure; we therefore interpret these findings as sensitivity analyses that complement the baseline results. Within this framework, participants with higher cumulative TyG-BA exposure experienced substantially elevated CMM risks, with HRs of 5.17 and 4.75 for the highest versus lowest quartiles of cumulative TyG-Light-BA and TyG-KDM-BA, respectively. K-means clustering identified two-wave exposure-level groups, with the high-level groups showing HRs of 3.93 and 4.51 for the two composites. These estimates were generally larger than those observed in the baseline quartile analyses, suggesting that repeated assessment of metabolic-aging burden may capture temporal exposure patterns that are not fully represented by a single baseline measurement. Although the limited repeated measurements preclude strong causal inferences about cumulative effects, these results provide preliminary evidence that the temporal dimension of metabolic-aging burden warrants consideration in risk stratification.

The TyG-BA composites were also associated with multiple transitions across the cardiometabolic disease continuum in a full illness-death multi-state model. As far as we know, this is the initial study to explore the link between a metabolic-aging composite and the full progression of disease, from being disease-free to the first cardiometabolic disease, CMM, and death. Elevated TyG-BA levels were linked to greater risks in all five transitions: from a healthy state to developing cardiometabolic disease, from being disease-free to death, from initial cardiometabolic disease to CMM, and from the first disease to death, and from CMM to death. These results indicate that the burden of metabolic aging is linked to the risk of transitioning between disease stages, not just the initial onset of the disease. Individuals with greater combined metabolic stress and reduced physiological reserve may follow a faster clinical course once an initial cardiometabolic disease has occurred. Subtype-specific analyses revealed that the association with transition from first cardiometabolic disease to CMM was strongest when coronary heart disease[43–45] was the initial disease, with more modest associations observed for T2D and non-significant associations for stroke[46–49]. These subtype differences may reflect the distinct pathophysiological pathways underlying different cardiometabolic diseases, suggesting that the clinical utility of the TyG-BA composite may vary depending on the specific disease context.

We systematically evaluated the incremental predictive value of the TyG-BA composites through multiple discrimination metrics and nested model comparisons. At 96 months, adding TyG-Light-BA and TyG-KDM-BA to the base clinical model increased the IPCW-adjusted C-index by 0.015 and 0.024, respectively, with both improvements reaching statistical significance. Decision curve analysis showed that models including the composites provided a higher net clinical benefit over a wide range of threshold probabilities. In time-dependent ROC analyses, both composites showed numerically higher AUCs than TyG at the evaluated time points. However, these numerical differences were generally not statistically significant after multiplicity adjustment, except for TyG-KDM-BA at 8 years. We therefore refrain from claiming that any composite is clinically preferred for either short-term or long-term prediction. This timeOdependent variation, while observed numerically, should be considered hypothesis-generating and may inform the selection of composite indices for different clinical applications in future confirmatory studies.

Nested model comparisons clarified the role of the composites. Neither composite improved discrimination beyond models containing TyG and biological age as separate covariates, and models including both individual components showed slightly better overall model fit than the corresponding multiplicative composite models. These findings indicate that the TyG-BA composites primarily provide a parsimonious and interpretable summary of concurrent metabolic-aging burden rather than a statistically superior alternative to modeling TyG and biological age separately. That is, the incremental value of the composite over the base model should not be overinterpreted as superiority over a separate-component model that includes its constituent elements as distinct predictors. This distinction is important for clinical translation: the composite offers a single value that clinicians can interpret as an overall burden, which facilitates risk communication and may be useful for risk stratification. However, for precise statistical estimation, the separate components may retain advantages because they allow differential weighting of each dimension’s contribution. Thus, the composite may be best viewed as a parsimonious risk-stratification measure rather than a replacement for more comprehensive multivariable risk models.

From a clinical perspective, both TyG and biological age are derived from routinely available clinical biomarkers, making the proposed framework potentially accessible in primary care and population-based screening settings. The TyG index requires only fasting triglyceride(TG) and glucose measurements [50], while the simplified Light-BA algorithm uses creatinine, glucose, and C-reactive protein in addition to chronological age [51], all commonly measured in clinical practice. Evaluating both metabolic dysfunction and biological aging together could help pinpoint individuals at high risk for CMM, especially those who might be overlooked if only one factor is considered. In exploratory subgroup analyses, we examined heterogeneity by CKM stage, age, and frailty status. CKM stage was treated as a stratification variable for assessing risk heterogeneity rather than as a mechanistic indicator, given its conceptual overlap with components of the metabolic-aging framework. The attenuated associations observed among participants with CKM stage 4 may reflect advanced disease status, where the additional prognostic information provided by TyG-BA composites is reduced. For age and frailty, associations were stronger among younger adults (<65 years) and non-frail individuals, with effect estimates attenuating progressively from robust through pre-frail to frail subgroups. We interpret these findings as hypothesis-generating heterogeneity rather than as definitive clinical conclusions or evidence of causal effect modification. If confirmed in future research, these patterns suggest that the metabolic-aging framework may be most useful for early risk identification, before late-stage disease or substantial accumulation of health deficits, consistent with prevention strategies that prioritize early intervention.

The study has several strengths. First, it integrates metabolic dysfunction and biological aging as two complementary dimensions of cardiometabolic vulnerability. By combining TyG with biological age and using two independent biological-age algorithms, it evaluates whether the two dimensions can be jointly characterized. Second, the repeated measurements in CHARLS allowed us to examine the development of CMM and whether the metabolic-aging burden remained relevant over time. A multi-state framework further characterized its associations with transitions from a disease-free state to a first cardiometabolic disease, subsequent CMM, and death. Third, extensive sensitivity analyses, including competing-risk models, multiple imputation, complete-case analyses, alternative covariate adjustment sets, exclusion of early events, and medication adjustment, yielded broadly consistent results. Systematic comparisons with separate TyG and biological-age components and with multiplicative interaction models further clarified that the TyG-BA composite should be interpreted as a parsimonious summary of concurrent metabolic-aging burden rather than as a statistically superior replacement for its individual components. Finally, time-dependent discrimination, reclassification, and decision-curve analyses provided additional evidence on the potential clinical utility of the composite in CMM risk assessment.

Several limitations should be acknowledged. First, the TyG-BA composite indices are empirically created as multiplicative products rather than being formally validated constructs, and they require external validation in independent groups before being used clinically. The multiplicative form assumes that the joint burden of TyG and biological age is appropriately represented on a product scale, which may not necessarily reflect the true functional relationship between the two dimensions. Second, like all observational studies, it is not possible to completely rule out residual confounding, even with thorough covariate adjustments. Unmeasured or imprecisely measured confounders may have influenced the observed associations; we therefore frame our findings in terms of association and risk stratification rather than causal inference. Third, heart disease and stroke were identified through self-reported diagnoses by physicians, which could lead to misclassification. While these diagnoses have demonstrated reasonable validity in CHARLS, non-differential misclassification might reduce estimates towards the null, and differential misclassification cannot be entirely ruled out. Fourth, the cumulative exposure and trajectory analyses were limited by the availability of only two repeated measurements (2011 and 2015), which do not capture the full history of metabolic-aging exposure; studies with more frequent assessments are needed. Fifth, our study population consisted exclusively of Chinese adults aged ≥45 years from CHARLS; generalizability to other populations warrants further investigation. Finally, the exploratory subgroup and heterogeneity analyses should be approached carefully due to the number of comparisons, and the relatively small number of deaths from certain states (e.g., CMM to death, n=69) limited the precision of some multi-state estimates, which should be confirmed in larger cohorts.

## 5. Conclusion

Rather than proposing a superior biomarker, our findings support a complementary metabolic-aging framework for characterizing the risk and progression of CMM. TyG and biological age captured distinct but related dimensions of cardiometabolic vulnerability, and their combined elevation identified individuals at particularly high risk of CMM. The continuous TyG-BA composites provided a parsimonious summary of concurrent metabolic and biological-aging burden and were associated with incident CMM, longitudinal exposure patterns, and multiple transitions across the cardiometabolic disease continuum. Although incorporation of the composites improved risk discrimination beyond the prespecified clinical model, they did not outperform models retaining TyG and biological age as separate predictors. Thus, the TyG-BA composites should be viewed as convenient summary measures for metabolic-aging risk stratification rather than as superior replacements for component-based models. The observed heterogeneity across age, CKM stage, and frailty status was exploratory and hypothesis-generating. External validation in diverse populations and more frequent longitudinal assessments are warranted to determine the generalizability and clinical utility of this metabolic-aging framework.

## Data Availability

Data for this study were obtained from the CHARLS and are accessible via the project website (http://charls.pku.edu.cn)

## List of abbreviations

AP: Attributable proportion
BAA: Biological age acceleration
BMI: Body mass index
BUN: Blood urea nitrogen
CHARLS: China Health and Retirement Longitudinal Study
CHD: Coronary heart disease
CI: Confidence interval
CKM: Cardiovascular-kidney-metabolic
CMD: Cardiometabolic disease
CMM: Cardiometabolic multimorbidity
CRP: C-reactive protein
DCA: Decision curve analysis
eGFR: Estimated glomerular filtration rate
FI: Frailty Index
FPG: Fasting plasma glucose
HbA1c: Glycated hemoglobin
HDL: High-density lipoprotein
HR: Hazard ratio
IDI: Integrated discrimination improvement
IPCW: Inverse probability of censoring weighting
IR: Insulin resistance
KDM-BA: Klemera-Doubal method biological age
Light-BA: Light biological age
MICE: Multiple imputation by chained equations
NRI: Net reclassification improvement
PLT: Platelet count
RCS: Restricted cubic spline
RERI: Relative excess risk due to interaction
ROC: Receiver operating characteristic
SBP: Systolic blood pressure
SD: Standard deviation
sHR: Subdistribution hazard ratio
T2D: Type 2 diabetes
TG: Triglyceride
TyG: Triglyceride-glucose
TyG-BA: Triglyceride-glucose-biological age
UA: Uric acid
VIF: Variance inflation factor

## Declarations

## Ethics approval and consent to participate

The CHARLS study has been approved by the Biomedical Ethics Review Committee of Peking University (Approval No.: IRB00001052-11015). All participants signed written informed consent forms prior to enrollment. All procedures involving human subjects were conducted in accordance with the ethical standards of the institutional research ethics committee and the principles of the Declaration of Helsinki. The reporting of this study follows the Strengthening the Reporting of Observational Studies in Epidemiology (STROBE) guidelines.

## Consent for publication

Not applicable.

## Authors’ contributions

Baishuang Yang and Shasha Yang conceived and designed the study, extracted and curated the data, performed the statistical analyses, and drafted the manuscript. Qiong Chen reviewed the data and revised the manuscript for important intellectual content. All authors read and approved the final version of the manuscript.

## Acknowledgements

We are grateful to all participants for study design, data collection and management, and invaluable contributions.

## Authors’ information

Not applicable.

