## Supplementary material for "Joint contributions of metabolic dysfunction and biological aging to cardiometabolic multimorbidity and disease progression: a prospective cohort study": N/A

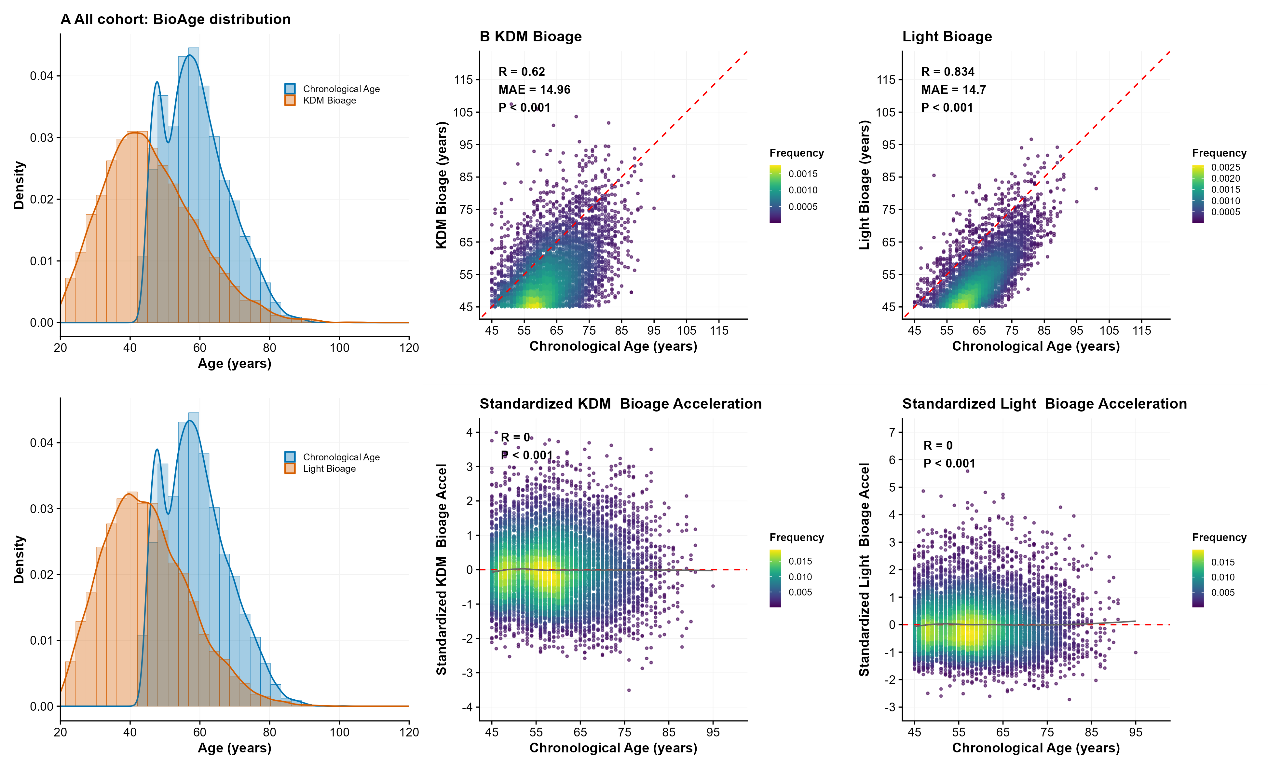

**Supplementary Figure 1. Correlation between biological age measures and chronological age, and distribution of biological age acceleration.**

Scatter colour gradients reflect participant density. (A, D) Density distributions of chronological age alongside KDM-BA (A) and Light-BA (D). (B, C) Correlations between biological age (KDM-BA, Light-BA) and chronological age. (E, F) Associations of standardized biological age acceleration (KDM, Light) with chronological age; red dashed lines represent zero acceleration. R, Pearson correlation coefficient; MAE, mean absolute error.

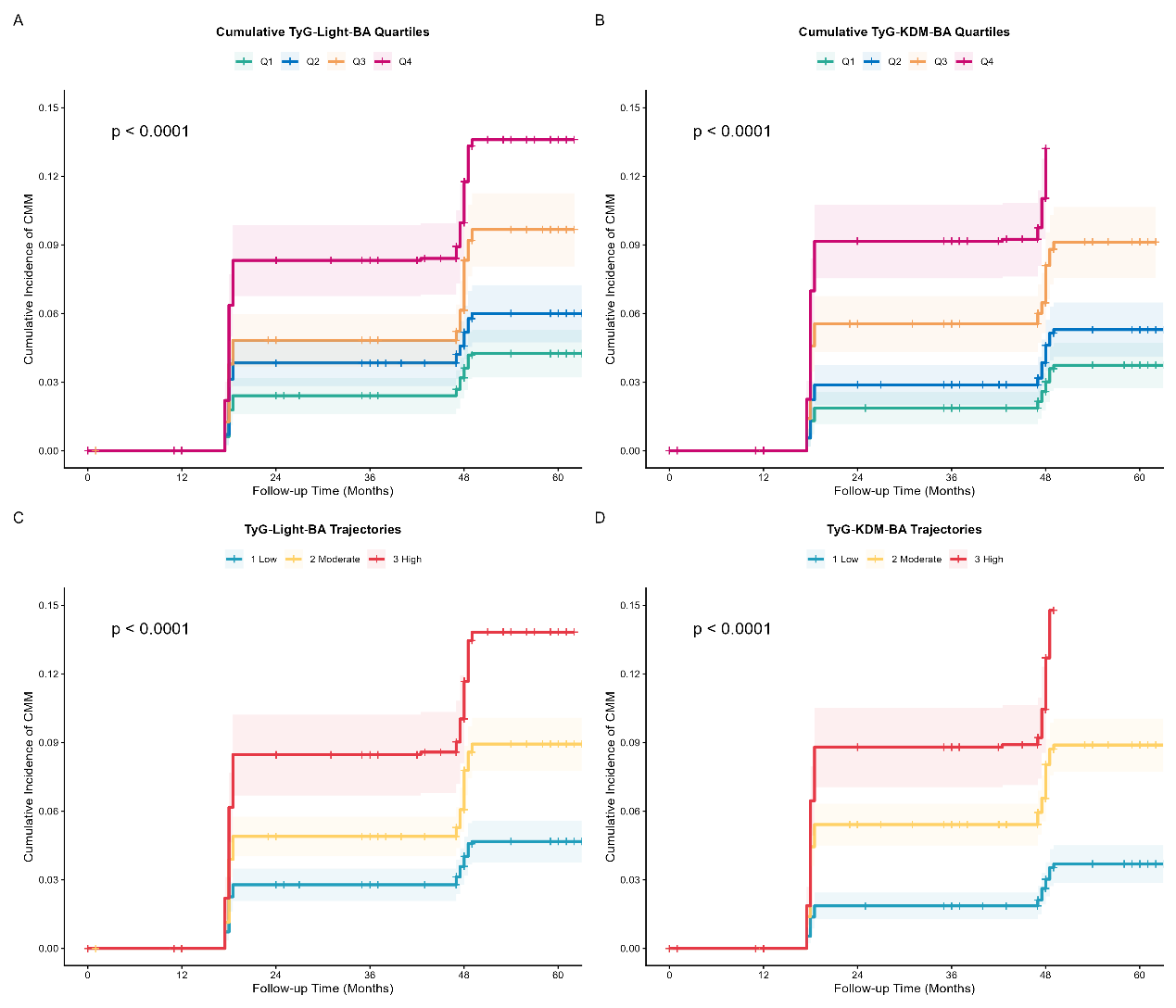

**Supplementary Figure 2. Cumulative incidence of CMM stratified by quartiles of cumTyG-BA and longitudinal trajectory clusters of TyG-BA**

Cumulative incidence curves using competing-risk functions. Shaded bands represent 95% confidence intervals; global log-rank *P* values are shown. (A) Quartiles of cumulative TyG-Light-BA; (B) quartiles of cumulative TyG-KDM-BA; (C) trajectory clusters of TyG-Light-BA (1 = Low, 2 = Moderate, 3 = High); (D) trajectory clusters of TyG-KDM-BA (1 = Low, 2 = Moderate, 3 = High).

*Abbreviations:* TyG, triglyceride-glucose index; cumTyG-BA, cumulative TyG–biological age index; Light-BA, Light-method biological age; KDM-BA, KDM biological age; CMM, composite cardiometabolic multimorbidity.

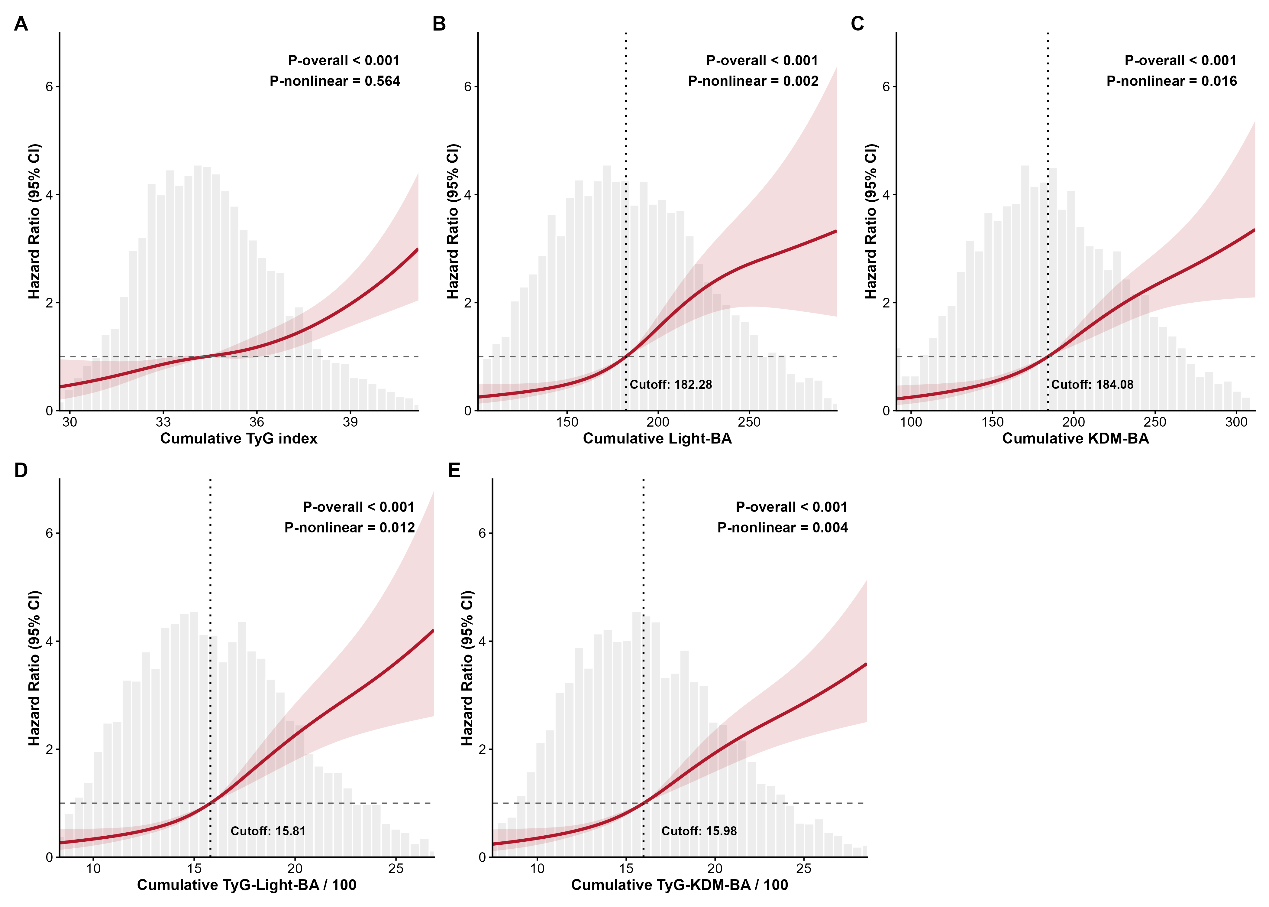

**Supplementary Figure 3. Nonlinear dose-response associations of cumulative exposure indices with CMM risk.**

Restricted cubic spline curves for adjusted HR (95% CIs). Grey dashed line: HR = 1. Histograms show variable distribution; vertical dotted lines mark cutoff values. (A) Cumulative TyG index; (B) Cumulative Light-BA; (C) Cumulative KDM-BA; (D) Cumulative TyG-Light-BA (/100); (E) Cumulative TyG-KDM-BA (/100). *P-overall*, overall association; *P-nonlinear*, test for nonlinearity.

*Abbreviations:* TyG, triglyceride-glucose index; Light-BA, Light biological age; KDM-BA, KDM biological age; CMM, composite cardiometabolic multimorbidity; HR, hazard ratio; CI, confidence interval.

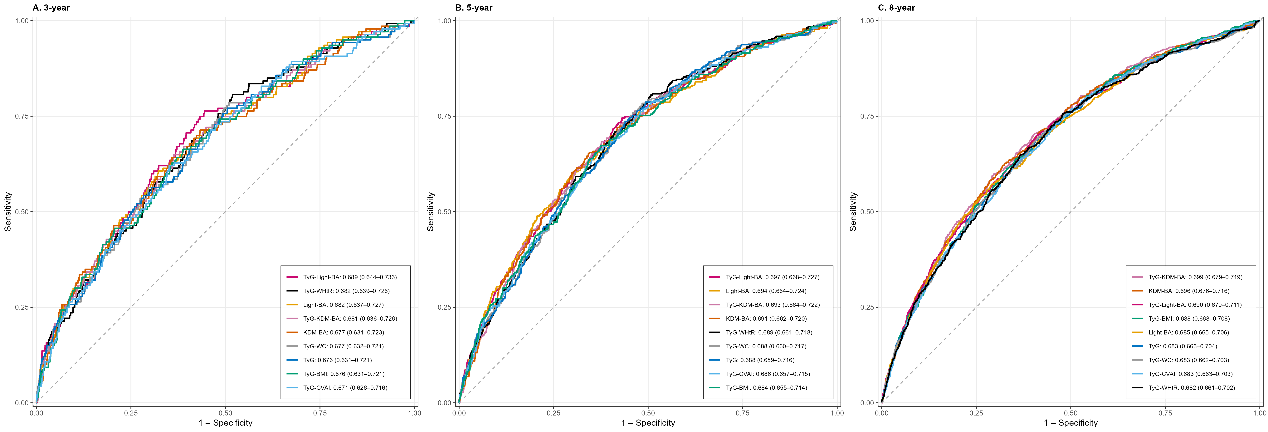
**Supplementary Figure 4. Time-dependent ROC curves for TyG, TyG-Light-BA, and TyG-KDM-BA at different prediction horizons.**

ROC curves display sensitivity versus 1-specificity for 3‑year (A), 5‑year (B) and 8‑year (C) follow‑up periods. Area under the curve (AUC) values are displayed in each panel.

*Abbreviations:* TyG, triglyceride-glucose index; Light-BA, Light-method biological age; KDM-BA, Klemara-Doubal method biological age; ROC, receiver operating characteristic; AUC, area under the curve.

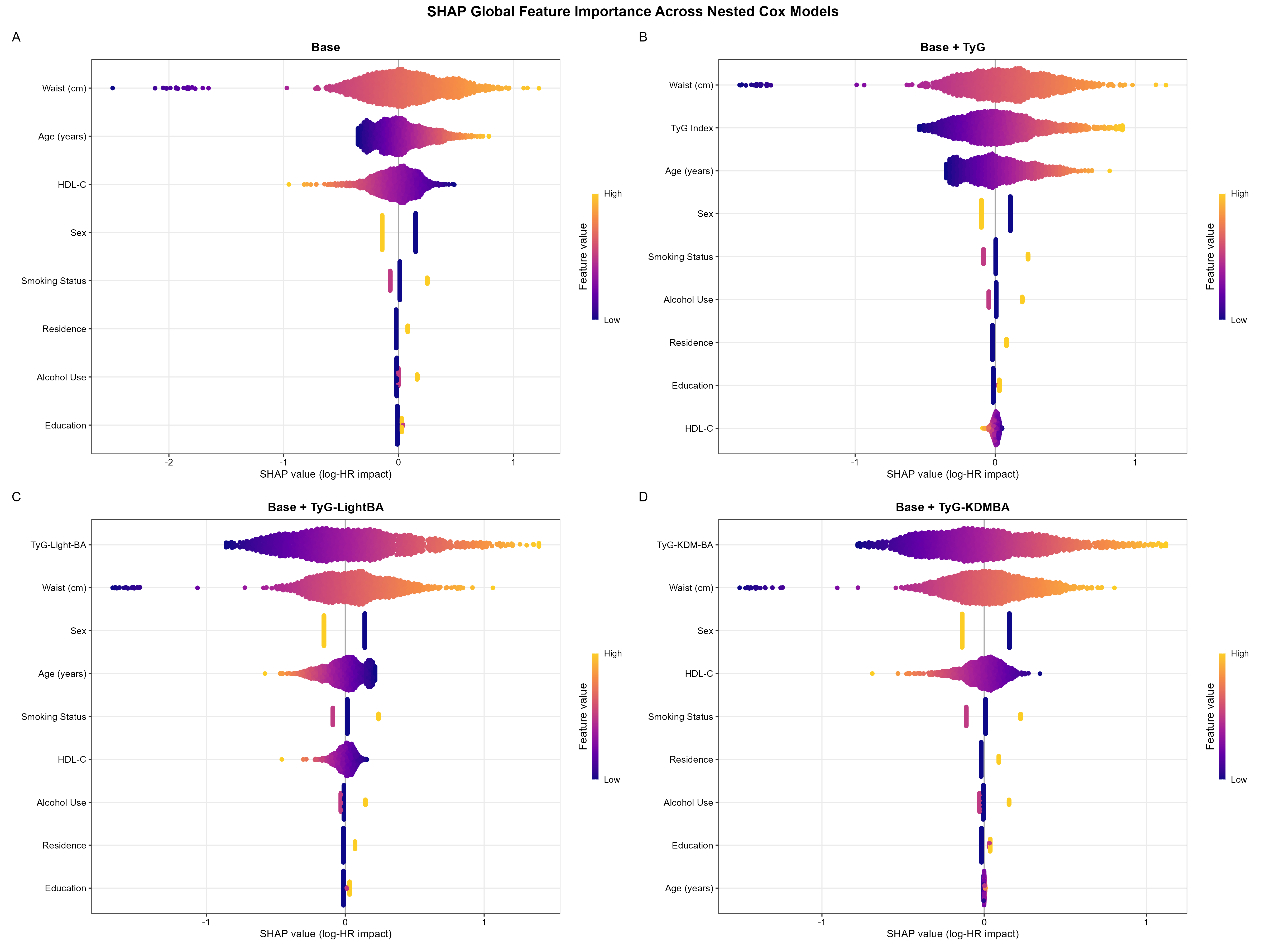

**Supplementary Figure 5. SHAP summary plots showing feature contributions to risk prediction across nested Cox models.**

(A) Base model; (B) base model plus TyG index; (C) base model plus TyG-Light-BA composite index; (D) base model plus TyG-KDM-BA composite index. SHAP values correspond to log hazard ratio contributions; color gradient indicates low (purple) to high (yellow) feature values.

Abbreviations: SHAP, SHapley Additive exPlanations; TyG, triglyceride-glucose index; KDM-BA, Klemara-Doubal method biological age; Light-BA, Light method biological age.

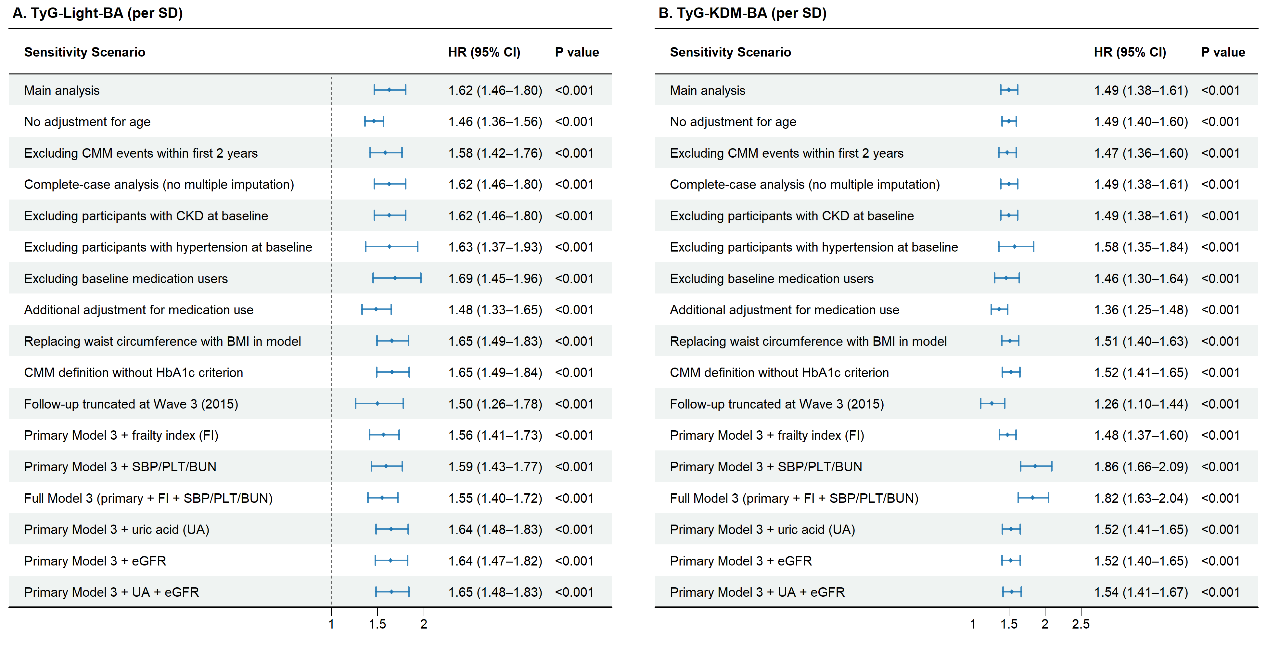

**Supplementary Figure 6. Sensitivity analyses for associations between TyG-BA composite indices and incident CMM.**

Forest plots display hazard ratios and 95% confidence intervals under alternative analytical scenarios. The primary model result is shown as the reference.

*Abbreviations:* TyG, triglyceride-glucose index; Light-BA, Light-method biological age; KDM-BA, Klemara-Doubal method biological age; CMM, composite cardiometabolic multimorbidity; CKD, chronic kidney disease; SBP, systolic blood pressure; PLT, platelet; BUN, blood urea nitrogen; UA, uric acid; eGFR, estimated glomerular filtration rate; HR, hazard ratio; CI, confidence interval.
