## Supplementary material for "Joint contributions of metabolic dysfunction and biological aging to cardiometabolic multimorbidity and disease progression: a prospective cohort study": N/A

**Supplementary Table 1. Baseline characteristics by joint TyG and KDM‑method biological age acceleration groups.**

| **Characteristic** | **Overall** N = 8767^1^ | **Low TyG & Non-accelerated Aging** N = 2693^1^ | **Low TyG & Accelerated Aging** N = 1691^1^ | **High TyG & Non-accelerated Aging** N = 2002^1^ | **High TyG & Accelerated Aging** N = 2381^1^ | ***P*-value**^2^ |
| --- | --- | --- | --- | --- | --- | --- |
| **Age (years), Mean ± SD** | 58.97 ± 9.21 | 58.85 ± 9.31 | 59.17 ± 9.62 | 58.93 ± 9.00 | 58.99 ± 8.97 | 0.849 |
| **Sex, n (%)** |  |  |  |  |  | <0.001 |
| Female | 4,680.0 (53.4%) | 1,345.0 (49.9%) | 799.0 (47.3%) | 1,171.0 (58.5%) | 1,365.0 (57.3%) |  |
| Male | 4,087.0 (46.6%) | 1,348.0 (50.1%) | 892.0 (52.7%) | 831.0 (41.5%) | 1,016.0 (42.7%) |  |
| **Educational attainment, n (%)** |  |  |  |  |  | 0.669 |
| Elementary school or below | 6,183.0 (70.5%) | 1,914.0 (71.1%) | 1,198.0 (70.8%) | 1,422.0 (71.0%) | 1,649.0 (69.3%) |  |
| Middle school | 2,336.0 (26.6%) | 703.0 (26.1%) | 452.0 (26.7%) | 525.0 (26.2%) | 656.0 (27.6%) |  |
| College or above | 248.0 (2.8%) | 76.0 (2.8%) | 41.0 (2.4%) | 55.0 (2.7%) | 76.0 (3.2%) |  |
| **Marital status, n (%)** |  |  |  |  |  | 0.440 |
| Married/Partnered | 7,337.0 (94.0%) | 2,274.0 (93.9%) | 1,386.0 (93.4%) | 1,687.0 (94.7%) | 1,990.0 (94.0%) |  |
| Unmarried/Other | 469.0 (6.0%) | 149.0 (6.1%) | 98.0 (6.6%) | 94.0 (5.3%) | 128.0 (6.0%) |  |
| **Residence, n (%)** |  |  |  |  |  | <0.001 |
| Rural | 7,327.0 (83.6%) | 2,304.0 (85.6%) | 1,451.0 (85.9%) | 1,657.0 (82.9%) | 1,915.0 (80.5%) |  |
| Urban | 1,436.0 (16.4%) | 389.0 (14.4%) | 239.0 (14.1%) | 343.0 (17.2%) | 465.0 (19.5%) |  |
| **Smoking status, n (%)** |  |  |  |  |  | <0.001 |
| Never smoker | 5,349.0 (61.2%) | 1,607.0 (59.9%) | 938.0 (55.7%) | 1,278.0 (64.1%) | 1,526.0 (64.3%) |  |
| Former smoker | 744.0 (8.5%) | 215.0 (8.0%) | 153.0 (9.1%) | 164.0 (8.2%) | 212.0 (8.9%) |  |
| Current smoker | 2,644.0 (30.3%) | 861.0 (32.1%) | 594.0 (35.3%) | 552.0 (27.7%) | 637.0 (26.8%) |  |
| **Drinking status, n (%)** |  |  |  |  |  | <0.001 |
| Never drinker | 5,333.0 (60.9%) | 1,614.0 (60.0%) | 937.0 (55.4%) | 1,275.0 (63.8%) | 1,507.0 (63.3%) |  |
| Former drinker | 709.0 (8.1%) | 206.0 (7.7%) | 155.0 (9.2%) | 150.0 (7.5%) | 198.0 (8.3%) |  |
| Current drinker | 2,714.0 (31.0%) | 870.0 (32.3%) | 598.0 (35.4%) | 572.0 (28.6%) | 674.0 (28.3%) |  |
| **Body mass index (kg/m²), Mean ± SD** | 23.44 ± 3.82 | 22.12 ± 3.38 | 23.15 ± 3.76 | 23.48 ± 3.62 | 25.09 ± 3.88 | <0.001 |
| **Waist circumference (cm), Mean ± SD** | 84.11 ± 12.38 | 80.13 ± 11.29 | 83.78 ± 11.28 | 84.20 ± 12.61 | 88.74 ± 12.54 | <0.001 |
| **SBP (mmHg), Mean ± SD** | 129.27 ± 21.33 | 117.11 ± 13.81 | 141.95 ± 21.36 | 118.74 ± 13.48 | 142.88 ± 20.56 | <0.001 |
| **DBP (mmHg), Mean ± SD** | 75.28 ± 12.18 | 69.22 ± 9.61 | 81.15 ± 11.83 | 70.33 ± 9.59 | 82.14 ± 11.44 | <0.001 |
| **CKM stage, n (%)** |  |  |  |  |  | <0.001 |
| Stage 0 | 1,063.0 (12.4%) | 709.0 (27.0%) | 136.0 (8.2%) | 178.0 (9.1%) | 40.0 (1.7%) |  |
| Stage 1 | 2,967.0 (34.7%) | 1,195.0 (45.5%) | 423.0 (25.5%) | 933.0 (47.8%) | 416.0 (17.9%) |  |
| Stage 2 | 3,062.0 (35.8%) | 399.0 (15.2%) | 785.0 (47.3%) | 528.0 (27.0%) | 1,350.0 (58.1%) |  |
| Stage 3 | 574.0 (6.7%) | 87.0 (3.3%) | 122.0 (7.4%) | 92.0 (4.7%) | 273.0 (11.7%) |  |
| Stage 4 | 896.0 (10.5%) | 237.0 (9.0%) | 192.0 (11.6%) | 221.0 (11.3%) | 246.0 (10.6%) |  |
| **Hypertension, n (%)** | 3,528.0 (40.2%) | 447.0 (16.6%) | 1,014.0 (60.0%) | 483.0 (24.1%) | 1,584.0 (66.5%) | <0.001 |
| **Diabetes, n (%)** | 1,246.0 (14.2%) | 118.0 (4.4%) | 109.0 (6.4%) | 309.0 (15.4%) | 710.0 (29.8%) | <0.001 |
| **Hyperlipidemia, n (%)** | 5,830.0 (66.5%) | 1,118.0 (41.5%) | 915.0 (54.1%) | 1,655.0 (82.7%) | 2,142.0 (90.0%) | <0.001 |
| **Cancer history, n (%)** | 72.0 (0.8%) | 22.0 (0.8%) | 11.0 (0.7%) | 20.0 (1.0%) | 19.0 (0.8%) | 0.703 |
| **Blood urea nitrogen (mg/dL), Mean ± SD** | 15.73 ± 4.47 | 14.87 ± 3.78 | 17.88 ± 5.18 | 14.16 ± 3.47 | 16.51 ± 4.65 | <0.001 |
| **Creatinine (mg/dL), Mean ± SD** | 0.78 ± 0.22 | 0.74 ± 0.16 | 0.83 ± 0.33 | 0.74 ± 0.16 | 0.82 ± 0.23 | <0.001 |
| **eGFR (mL/min/1.73m²), Mean ± SD** | 96.02 ± 14.04 | 99.39 ± 11.43 | 93.75 ± 14.83 | 97.86 ± 12.27 | 92.27 ± 16.18 | <0.001 |
| **Uric acid (mg/dL), Mean ± SD** | 4.44 ± 1.24 | 4.11 ± 1.08 | 4.56 ± 1.21 | 4.32 ± 1.15 | 4.83 ± 1.38 | <0.001 |
| **HbA1c (%), Mean ± SD** | 5.24 ± 0.78 | 5.04 ± 0.40 | 5.18 ± 0.54 | 5.14 ± 0.51 | 5.60 ± 1.20 | <0.001 |
| **Fasting glucose (mg/dL), Mean ± SD** | 108.70 ± 34.24 | 97.17 ± 13.74 | 99.43 ± 15.87 | 110.44 ± 27.88 | 126.87 ± 52.22 | <0.001 |
| **Total cholesterol (mg/dL), Mean ± SD** | 193.49 ± 38.37 | 178.65 ± 31.78 | 193.77 ± 35.90 | 190.28 ± 34.55 | 212.76 ± 41.63 | <0.001 |
| **Triglycerides (mg/dL), Mean ± SD** | 131.57 ± 104.01 | 75.34 ± 20.55 | 76.73 ± 20.76 | 170.75 ± 88.85 | 201.19 ± 143.43 | <0.001 |
| **HDL cholesterol (mg/dL), Mean ± SD** | 51.43 ± 15.28 | 57.17 ± 14.28 | 58.34 ± 15.48 | 45.45 ± 13.15 | 45.06 ± 13.11 | <0.001 |
| **LDL cholesterol (mg/dL), Mean ± SD** | 116.52 ± 34.89 | 108.69 ± 28.18 | 121.74 ± 32.37 | 110.99 ± 33.54 | 126.37 ± 41.06 | <0.001 |
| **C-reactive protein (mg/L), Mean ± SD** | 2.65 ± 7.19 | 1.32 ± 2.87 | 4.75 ± 11.61 | 1.45 ± 3.72 | 3.66 ± 8.16 | <0.001 |
| **Platelet count (×10⁹/L), Mean ± SD** | 212.07 ± 75.89 | 209.17 ± 74.47 | 210.41 ± 72.75 | 211.74 ± 75.89 | 216.79 ± 79.42 | <0.001 |
| **Mean corpuscular volume (fL), Mean ± SD** | 90.68 ± 8.64 | 90.60 ± 9.45 | 90.70 ± 8.49 | 91.03 ± 8.26 | 90.44 ± 8.06 | 0.002 |
| **White blood cell count (×10⁹/L), Mean ± SD** | 6.23 ± 1.88 | 5.84 ± 1.74 | 6.33 ± 1.94 | 6.12 ± 1.74 | 6.69 ± 1.98 | <0.001 |
| **Frailty Index, Mean ± SD** | 0.15 ± 0.11 | 0.14 ± 0.11 | 0.16 ± 0.11 | 0.16 ± 0.11 | 0.16 ± 0.11 | <0.001 |
| **TyG index, Mean ± SD** | 8.67 ± 0.63 | 8.16 ± 0.29 | 8.20 ± 0.28 | 9.05 ± 0.39 | 9.25 ± 0.53 | <0.001 |
| **Light-AgeAccel (years), Mean ± SD** | -0.03 ± 6.48 | -3.28 ± 4.99 | 2.01 ± 6.95 | -1.81 ± 4.98 | 3.69 ± 6.33 | <0.001 |
| **KDM-AgeAccel (years), Mean ± SD** | -0.05 ± 10.21 | -8.27 ± 5.41 | 7.69 ± 6.46 | -6.82 ± 4.97 | 9.43 ± 7.28 | <0.001 |
| **TyG-Light-BA, Mean ± SD** | 387.39 ± 112.08 | 336.91 ± 95.44 | 385.45 ± 108.34 | 386.39 ± 102.47 | 446.71 ± 111.34 | <0.001 |
| **TyG-KDM-BA, Mean ± SD** | 394.54 ± 124.91 | 302.25 ± 80.61 | 437.38 ± 97.26 | 348.42 ± 85.32 | 507.28 ± 108.77 | <0.001 |
| **Incident CMM events, n (%)** | 873.0 (10.0%) | 144.0 (5.3%) | 152.0 (9.0%) | 201.0 (10.0%) | 376.0 (15.8%) | <0.001 |
| ^1^Kruskal-Wallis rank sum test; Pearson's Chi-squared test | | | | | | |

**Supplementary Table 1. Baseline characteristics by joint TyG and KDM‑method biological age acceleration groups.**

Data are presented as mean (SD) for continuous variables and n (%) for categorical variables. Joint groups were defined by median TyG (8.67) and KDM‑age acceleration (cutoff: 0 years). *P* values from Kruskal–Wallis or chi‑squared tests across groups.

*Abbreviations:* TyG, triglyceride‑glucose index; KDM‑AgeAccel, Klemera–Doubal method age acceleration; CKM, cardiovascular‑kidney‑metabolic; eGFR, estimated glomerular filtration rate; HDL, high‑density lipoprotein; LDL, low‑density lipoprotein.

| Characteristic | Crude Model | | Model 1 | | Model 2 | | Model 3 | |
| --- | --- | --- | --- | --- | --- | --- | --- | --- |
|  | HR (95% CI) | P value | HR (95% CI) | P value | HR (95% CI) | P value | HR (95% CI) | P value |
| **TyG × Light Accelerated Aging** | | | | | | | | |
| TyG status × Light Accelerated aging status (multiplicative interaction) | 0.94 (0.70, 1.24) | 0.647 | 0.93 (0.70, 1.24) | 0.620 | 0.93 (0.70, 1.24) | 0.636 | 0.89 (0.67, 1.18) | 0.418 |
| RERI | 0.566 (-0.071, 1.203) |  | 0.515 (-0.738, 1.768) |  | 0.506 (-0.735, 1.747) |  | 0.101 (-0.832, 1.034) |  |
| AP | 0.169 (0.011, 0.327) |  | 0.158 (-0.166, 0.482) |  | 0.157 (-0.168, 0.482) |  | 0.043 (-0.335, 0.420) |  |
| S (Synergy index) | 1.317 |  | 1.296 |  | 1.295 |  | 1.080 |  |
| **TyG × KDM Accelerated Aging** | | | | | | | | |
| TyG status × KDM Accelerated aging status (multiplicative interaction) | 0.95 (0.71, 1.26) | 0.698 | 0.94 (0.71, 1.25) | 0.671 | 0.94 (0.70, 1.24) | 0.645 | 0.93 (0.70, 1.23) | 0.593 |
| RERI | 0.531 (-0.090, 1.152) |  | 0.482 (-0.738, 1.701) |  | 0.462 (-0.748, 1.671) |  | 0.133 (-0.763, 1.028) |  |
| AP | 0.164 (0.004, 0.325) |  | 0.154 (-0.176, 0.483) |  | 0.148 (-0.183, 0.480) |  | 0.059 (-0.314, 0.432) |  |
| S (Synergy index) | 1.312 |  | 1.291 |  | 1.280 |  | 1.118 |  |
| *Abbreviations: HR, Hazard Ratio; CI, Confidence Interval; Multiplicative interaction: Cox model interaction term HR and P value. Additive interaction: RERI (Relative Excess Risk due to Interaction), AP (Attributable Proportion due to interaction), S (Synergy index). RERI >0, AP>0, S>1 indicate significant additive synergistic effect. Model 1: Adjusted for baseline age, sex, education attainment, and residence. Model 2: Model 1 + smoking status and alcohol consumption. Model 3: Model 2 + waist circumference, HDL-C.* | | | | | | | | |

**Supplementary Table 2. Multiplicative and additive interactions between TyG status and biological age acceleration for incident CMM.**

**Supplementary Table 2. Multiplicative and additive interactions between TyG status and biological age acceleration for incident CMM.**

HRs and 95% CIs from Cox models with multiplicative interaction terms (TyG status × BAA status). Additive interaction quantified by RERI, AP, and Synergy index (S); RERI > 0, AP > 0, and S > 1 indicate positive additive synergy. TyG status: high vs. low (median cutoff 8.67); BAA: accelerated (>0 years) vs. non‑accelerated (≤0). Models adjusted as described in Table 2.

*Abbreviations:* HR, hazard ratio; CI, confidence interval; TyG, triglyceride‑glucose index; BAA, biological age acceleration; RERI, relative excess risk due to interaction; AP, attributable proportion due to interaction.

**Supplementary Table 3. Cumulative exposure indices and two-waves trajectory clusters of TyG-BA composites in relation to incident CMM (Wave 3 landmark subcohort)**

| Characteristic | Crude Model | | Model 1 | | Model 2 | | Model 3 | |
| --- | --- | --- | --- | --- | --- | --- | --- | --- |
|  | HR (95% CI) | P value | HR (95% CI) | P value | HR (95% CI) | P value | HR (95% CI) | P value |
| **Single cumulative continuous biomarkers (per SD, Wave3 subcohort)** | | | | | | | | |
| Cumulative TyG index (per SD) | 1.53 (1.40, 1.67) | < 0.001 | 1.53 (1.40, 1.67) | < 0.001 | 1.52 (1.39, 1.67) | < 0.001 | 1.41 (1.27, 1.56) | < 0.001 |
| Cumulative Light biological age (per SD) | 1.41 (1.29, 1.55) | < 0.001 | 2.33 (1.96, 2.78) | < 0.001 | 2.33 (1.95, 2.78) | < 0.001 | 2.01 (1.66, 2.43) | < 0.001 |
| Cumulative KDM biological age (per SD) | 1.64 (1.50, 1.80) | < 0.001 | 2.02 (1.79, 2.27) | < 0.001 | 2.01 (1.79, 2.27) | < 0.001 | 1.85 (1.63, 2.10) | < 0.001 |
| **Combined cumulative TyG-BA index (per SD)** | | | | | | | | |
| Cumulative TyG-Light-BA (per SD) | 1.55 (1.42, 1.70) | < 0.001 | 2.16 (1.89, 2.48) | < 0.001 | 2.16 (1.88, 2.47) | < 0.001 | 1.94 (1.66, 2.25) | < 0.001 |
| Cumulative TyG-KDM-BA (per SD) | 1.73 (1.58, 1.89) | < 0.001 | 1.96 (1.76, 2.17) | < 0.001 | 1.95 (1.76, 2.17) | < 0.001 | 1.79 (1.60, 2.00) | < 0.001 |
| **Cumulative TyG-Light-BA Quartiles** | | | | | | | | |
| Q1 | Ref |  | Ref |  | Ref |  | Ref |  |
| Q2 | 1.43 (1.03, 1.99) | 0.035 | 1.87 (1.33, 2.63) | < 0.001 | 1.86 (1.32, 2.62) | < 0.001 | 1.67 (1.18, 2.36) | 0.004 |
| Q3 | 2.32 (1.71, 3.15) | < 0.001 | 3.79 (2.68, 5.36) | < 0.001 | 3.75 (2.65, 5.30) | < 0.001 | 3.04 (2.12, 4.34) | < 0.001 |
| Q4 | 3.37 (2.51, 4.54) | < 0.001 | 7.10 (4.80, 10.51) | < 0.001 | 6.97 (4.70, 10.33) | < 0.001 | 5.17 (3.40, 7.87) | < 0.001 |
| **Cumulative TyG-KDM-BA Quartiles** | | | | | | | | |
| Q1 | Ref |  | Ref |  | Ref |  | Ref |  |
| Q2 | 1.43 (1.00, 2.05) | 0.048 | 1.68 (1.17, 2.41) | 0.005 | 1.66 (1.16, 2.39) | 0.006 | 1.52 (1.05, 2.18) | 0.025 |
| Q3 | 2.53 (1.83, 3.50) | < 0.001 | 3.24 (2.31, 4.56) | < 0.001 | 3.19 (2.27, 4.49) | < 0.001 | 2.72 (1.92, 3.86) | < 0.001 |
| Q4 | 4.42 (3.26, 6.01) | < 0.001 | 6.20 (4.39, 8.76) | < 0.001 | 6.11 (4.32, 8.64) | < 0.001 | 4.75 (3.31, 6.82) | < 0.001 |
| **TyG-Light-BA Trajectory Clusters** | | | | | | | | |
| 1 Low | Ref |  | Ref |  | Ref |  | Ref |  |
| 2 Moderate | 1.94 (1.53, 2.48) | < 0.001 | 2.65 (2.01, 3.48) | < 0.001 | 2.63 (2.00, 3.46) | < 0.001 | 2.20 (1.66, 2.92) | < 0.001 |
| 3 High | 3.11 (2.38, 4.05) | < 0.001 | 5.50 (3.80, 7.97) | < 0.001 | 5.44 (3.76, 7.89) | < 0.001 | 3.93 (2.65, 5.82) | < 0.001 |
| **TyG-KDM-BA Trajectory Clusters** | | | | | | | | |
| 1 Low | Ref |  | Ref |  | Ref |  | Ref |  |
| 2 Moderate | 2.50 (1.91, 3.26) | < 0.001 | 2.95 (2.24, 3.91) | < 0.001 | 2.92 (2.21, 3.87) | < 0.001 | 2.57 (1.93, 3.41) | < 0.001 |
| 3 High | 4.44 (3.36, 5.87) | < 0.001 | 5.81 (4.22, 8.00) | < 0.001 | 5.78 (4.20, 7.96) | < 0.001 | 4.51 (3.23, 6.30) | < 0.001 |
| *Abbreviations: HR, Hazard Ratio; CI, Confidence Interval; TyG, Triglyceride-Glucose index; BA, Biological Age; Ref, Reference. Model 1: Adjusted for Wave 3 age, sex, education attainment, and residence. Model 2: Model 1 + smoking status and alcohol consumption. Model 3 (primary): Model 2 + Wave 3 waist circumference and HDL-C. Sensitivity models additionally adjust for frailty index (FI) and/or KDM-BA input components (SBP, PLT, BUN). Note 1: The W3 subcohort is a landmark analysis; participants had to survive and complete Wave 3 (2015) measurements, and follow-up starts at Wave 3. This conditions on survival to 2015 (selection on healthy survivors). Note 2: Trajectory labels describe two measured waves (2011 and 2015) only; cluster-number sensitivity (k = 2 and k = 4) and bootstrap cluster stability are reported in the supplementary outputs. Note 3: Cumulative-exposure models additionally adjusted for the baseline composite are reported separately (Table S3b) to formally test the 'beyond baseline assessment' claim.* | | | | | | | | |

**Supplementary Table 3. Cumulative exposure indices and two-waves trajectory clusters of TyG-BA composites in relation to incident CMM (Wave 3 landmark subcohort)**

HRs and 95% CIs per 1‑SD increase (cumulative indices) or versus cluster 1 (trajectory clusters). Trajectory clusters derived from K‑means (k = 3) using Wave 1 and Wave 3 measurements. Models adjusted as described in Table 2, using Wave 3 covariates. The Wave 3 subcohort conditions on survival to and completion of the 2015 wave (landmark analysis). Sensitivity analyses with k = 2, k = 4, and bootstrap cluster stability are reported separately.

*Abbreviations:* HR, hazard ratio; CI, confidence interval; TyG, triglyceride‑glucose index; Light‑BA, Light biological age; KDM‑BA, Klemera–Doubal biological age; CMM, cardiometabolic multimorbidity; Ref, reference.

**Supplementary Table 4. Markov assumption check of the association of global state entry time with each transition hazard.**

| Transition | Events | HR of entry time (per 12 months) | P value |
| --- | --- | --- | --- |
| CMD-free → First CMD | 2,167 | NaN (NaN, NaN) |  |
| CMD-free → Death | 476 | NaN (NaN, NaN) |  |
| First CMD → CMM | 711 | 0.89 (0.85, 0.93) | < 0.001 |
| First CMD → Death | 431 | 0.99 (0.94, 1.05) | 0.857 |
| CMM → Death | 69 | 0.82 (0.69, 0.98) | 0.025 |
| A significant HR for entry_global_time indicates that the transition hazard depends on when the | | | |
| state was entered, providing evidence against the Markov assumption. | | | |
| Transitions from the initial CMD-free state cannot be tested via entry time because all participants enter that state at global time 0; the clock-reset sensitivity analysis in Supplementary Table 4 covers all five transitions. | | | |
| Models additionally include Model 3 covariates and TyG-LightBA (per SD); cluster(ID) robust SEs. | | | |
| Results were pooled across 5 multiply imputed datasets using Rubin's rules. | | | |

**Supplementary Table 4. Markov assumption check of the association of global state entry time with each transition hazard.**

HRs and 95% CIs for entry_global_time (per 12‑month increase in calendar time since study baseline at state entry), estimated from Cox models with robust cluster (ID) standard errors, adjusted for Model 3 covariates and TyG‑Light‑BA (per SD). A significant HR indicates time‑dependent transition hazards, violating the Markov assumption. Transitions from the initial CMD‑free state are not testable because all participants enter at global time 0; the clock‑reset (semi‑Markov) sensitivity analysis in Supplementary Table 5 covers all five transitions. Results pooled across 5 multiply imputed datasets.

*Abbreviations:* HR, hazard ratio; CI, confidence interval; CMD, cardiometabolic disease; CMM, cardiometabolic multimorbidity; TyG, triglyceride‑glucose index; Light‑BA, Light biological age.

**Supplementary Table 5. Clock‑reset (semi‑Markov) multi‑state Cox regression of TyG‑BA composites for CMM progression and mortality.**

| Pathway / Subgroup | Events / Total | HR (95% CI) | | HR (95% CI) | | HR (95% CI) | | HR (95% CI) | |
| --- | --- | --- | --- | --- | --- | --- | --- | --- | --- |
|  |  | Crude Model | *P‑value* | Model 1 | *P‑value* | Model 2 | *P‑value* | Model 3 | *P‑value* |
| **Pathway 1: CMD-free → First CMD** |  |  |  |  |  |  |  |  |  |
| TyG-Light-BA (per SD) | 2167 / 6596 | 1.31 (1.26, 1.37) | < 0.001 | 1.37 (1.28, 1.48) | < 0.001 | 1.37 (1.27, 1.47) | < 0.001 | 1.28 (1.18, 1.39) | < 0.001 |
| TyG-KDM-BA (per SD) | 2167 / 6596 | 1.34 (1.29, 1.40) | < 0.001 | 1.32 (1.25, 1.39) | < 0.001 | 1.32 (1.25, 1.39) | < 0.001 | 1.25 (1.18, 1.32) | < 0.001 |
| **Pathway 2: CMD-free → Mortality** |  |  |  |  |  |  |  |  |  |
| TyG-Light-BA (per SD) | 476 / 6596 | 2.41 (2.22, 2.63) | < 0.001 | 1.33 (1.14, 1.56) | < 0.001 | 1.33 (1.14, 1.55) | < 0.001 | 1.43 (1.22, 1.69) | < 0.001 |
| TyG-KDM-BA (per SD) | 476 / 6596 | 2.01 (1.85, 2.19) | < 0.001 | 1.19 (1.06, 1.33) | 0.003 | 1.18 (1.05, 1.32) | 0.004 | 1.21 (1.07, 1.36) | 0.002 |
| **Pathway 3: First CMD → CMM** |  |  |  |  |  |  |  |  |  |
| TyG-Light-BA (per SD) | 711 / 4182 | 1.22 (1.14, 1.32) | < 0.001 | 1.30 (1.18, 1.44) | < 0.001 | 1.30 (1.18, 1.43) | < 0.001 | 1.15 (1.03, 1.29) | 0.014 |
| TyG-KDM-BA (per SD) | 711 / 4182 | 1.26 (1.18, 1.35) | < 0.001 | 1.28 (1.18, 1.38) | < 0.001 | 1.28 (1.18, 1.38) | < 0.001 | 1.16 (1.07, 1.27) | < 0.001 |
| **Subtype: Type 2 Diabetes → CMM** |  |  |  |  |  |  |  |  |  |
| TyG-Light-BA (per SD) | 333 / 2065 | 1.21 (1.09, 1.34) | < 0.001 | 1.31 (1.14, 1.50) | < 0.001 | 1.30 (1.13, 1.49) | < 0.001 | 1.20 (1.03, 1.40) | 0.019 |
| TyG-KDM-BA (per SD) | 333 / 2065 | 1.28 (1.16, 1.41) | < 0.001 | 1.32 (1.18, 1.48) | < 0.001 | 1.31 (1.17, 1.47) | < 0.001 | 1.21 (1.07, 1.37) | 0.002 |
| **Subtype: Coronary Heart Disease → CMM** |  |  |  |  |  |  |  |  |  |
| TyG-Light-BA (per SD) | 317 / 1681 | 1.39 (1.23, 1.56) | < 0.001 | 1.81 (1.50, 2.19) | < 0.001 | 1.83 (1.51, 2.21) | < 0.001 | 1.51 (1.22, 1.87) | < 0.001 |
| TyG-KDM-BA (per SD) | 317 / 1681 | 1.44 (1.29, 1.61) | < 0.001 | 1.51 (1.32, 1.73) | < 0.001 | 1.52 (1.33, 1.73) | < 0.001 | 1.37 (1.19, 1.58) | < 0.001 |
| **Subtype: Stroke → CMM** |  |  |  |  |  |  |  |  |  |
| TyG-Light-BA (per SD) | 61 / 436 | 1.30 (1.00, 1.69) | 0.053 | 1.65 (1.13, 2.39) | 0.009 | 1.63 (1.11, 2.39) | 0.012 | 1.55 (1.00, 2.40) | 0.050 |
| TyG-KDM-BA (per SD) | 61 / 436 | 1.16 (0.89, 1.51) | 0.278 | 1.21 (0.88, 1.66) | 0.251 | 1.22 (0.88, 1.68) | 0.229 | 1.15 (0.82, 1.60) | 0.428 |
| **Pathway 4: First CMD → Mortality** |  |  |  |  |  |  |  |  |  |
| TyG-Light-BA (per SD) | 431 / 4182 | 2.08 (1.90, 2.27) | < 0.001 | 1.30 (1.14, 1.47) | < 0.001 | 1.30 (1.15, 1.48) | < 0.001 | 1.42 (1.24, 1.63) | < 0.001 |
| TyG-KDM-BA (per SD) | 431 / 4182 | 1.75 (1.60, 1.91) | < 0.001 | 1.25 (1.13, 1.39) | < 0.001 | 1.26 (1.13, 1.40) | < 0.001 | 1.32 (1.19, 1.48) | < 0.001 |
| **Pathway 5: CMM → Mortality** |  |  |  |  |  |  |  |  |  |
| TyG-Light-BA (per SD) | 69 / 873 | 2.13 (1.70, 2.67) | < 0.001 | 1.29 (0.94, 1.77) | 0.116 | 1.32 (0.95, 1.82) | 0.094 | 1.43 (1.03, 1.99) | 0.033 |
| TyG-KDM-BA (per SD) | 69 / 873 | 1.83 (1.46, 2.28) | < 0.001 | 1.36 (1.05, 1.77) | 0.021 | 1.37 (1.05, 1.79) | 0.021 | 1.47 (1.12, 1.93) | 0.006 |
| Abbreviations: HR, Hazard Ratio; CI, Confidence Interval; TyG, Triglyceride‑Glucose index; BA, Biological Age. | | | | | | | | | |
| Model 1: Adjusted for baseline age, sex, education attainment, and residence. | | | | | | | | | |
| Model 2: Model 1 + additionally adjusted for smoking and alcohol drinking status. | | | | | | | | | |
| Model 3: Model 2 + additionally adjusted for waist circumference and HDL‑cholesterol. | | | | | | | | | |
| Results were pooled across 5 multiply imputed datasets using Rubin's rules. | | | | | | | | | |
| The multi‑state model included five transitions: CMD‑free → First CMD, CMD‑free → Death, First CMD → CMM, First CMD → Death, and CMM → Death. Death was treated as an absorbing competing event. | | | | | | | | | |
| First CMD subtypes were defined using pattern matching; participants with multiple concurrent CMDs contribute to each corresponding subtype group. | | | | | | | | | |
| Baseline participants with pre‑existing CMD entered the First CMD state at time zero; time of first CMD onset was imputed using wave‑to‑wave midpoint. | | | | | | | | | |
| The time scale was reset to 0 at entry into each state (clock‑reset / semi‑Markov model). | | | | | | | | | |

**Supplementary Table 5. Clock‑reset (semi‑Markov) multi‑state Cox regression of TyG‑BA composites for CMM progression and mortality.**

HRs and 95% CIs per 1‑SD increase from clock‑reset (semi‑Markov) multi‑state models, where the time scale was reset to 0 at entry into each state. Transition definitions, CMD subtype grouping, and covariate adjustment are identical to Table 5 (models adjusted as described in Table 2). Results pooled across 5 multiply imputed datasets.

*Abbreviations:* HR, hazard ratio; CI, confidence interval; TyG, triglyceride‑glucose index; Light‑BA, Light biological age; KDM‑BA, Klemera–Doubal biological age; CMD, cardiometabolic disease; CMM, cardiometabolic multimorbidity.

**Supplementary Table 6. Subgroup analysis of TyG-Light-BA per SD increase for incident cardiometabolic multimorbidity.**

| **Subgroup** | **Events/Total** | **HR (95% CI)** | ***P* value** | ***P* for interaction** |
| --- | --- | --- | --- | --- |
| **Age, years** |  |  |  | **<0.001** |
| <65 | 618 / 6488 | 1.74 (1.57–1.92) | <0.001 |  |
| ≥65 | 255 / 2279 | 1.23 (1.04–1.47) | 0.017 |  |
| **Sex** |  |  |  | **0.168** |
| Female | 523 / 4680 | 1.81 (1.59–2.07) | <0.001 |  |
| Male | 350 / 4087 | 1.39 (1.17–1.64) | <0.001 |  |
| **BMI, kg/m²** |  |  |  | **0.363** |
| <24 | 365 / 5263 | 1.59 (1.35–1.86) | <0.001 |  |
| ≥24 | 508 / 3504 | 1.64 (1.43–1.89) | <0.001 |  |
| **Residence type** |  |  |  | **0.563** |
| Rural | 707 / 7330 | 1.59 (1.41–1.78) | <0.001 |  |
| Urban | 166 / 1437 | 1.85 (1.45–2.36) | <0.001 |  |
| **Education level** |  |  |  | **0.057** |
| Elementary or below | 630 / 6183 | 1.58 (1.40–1.79) | <0.001 |  |
| Middle school or above | 243 / 2584 | 1.75 (1.44–2.12) | <0.001 |  |
| **Smoking status** |  |  |  | **0.209** |
| Never | 578 / 5379 | 1.74 (1.54–1.98) | <0.001 |  |
| Ever | 295 / 3388 | 1.42 (1.18–1.70) | <0.001 |  |
| **Alcohol consumption** |  |  |  | **0.826** |
| Never | 562 / 5344 | 1.83 (1.61–2.09) | <0.001 |  |
| Ever | 311 / 3423 | 1.36 (1.14–1.61) | <0.001 |  |
| **Hypertension** |  |  |  | **0.017** |
| No | 336 / 5239 | 1.63 (1.37–1.93) | <0.001 |  |
| Yes | 537 / 3528 | 1.52 (1.33–1.73) | <0.001 |  |
| **Hyperlipidemia** |  |  |  | **0.305** |
| No | 197 / 2937 | 1.67 (1.30–2.14) | <0.001 |  |
| Yes | 676 / 5830 | 1.61 (1.43–1.80) | <0.001 |  |
| **CKM stage** |  |  |  | **0.019** |
| Stage 0–1 | 159 / 4030 | 1.60 (1.20–2.13) | 0.001 |  |
| Stage 2–3 | 433 / 3636 | 1.60 (1.39–1.84) | <0.001 |  |
| Stage 4 | 261 / 896 | 1.42 (1.12–1.79) | 0.003 |  |
| Missing/unclassifiable | 20 / 205 | 1.99 (1.05–3.76) | 0.034 |  |
| *Abbreviations: HR, hazard ratio; CI, confidence interval.* | | | | |
| *Models were adjusted for baseline covariates excluding the stratifying subgroup variable.* | | | | |
| *P for interaction derived from pooled D1/D3 likelihood ratio test or Wald test.* | | | | |
| *Missing data were handled using multiple imputation by chained equations (MICE).* | | | | |

**Supplementary Table 6. Subgroup analysis of TyG-Light-BA per SD increase for incident cardiometabolic multimorbidity.**

HRs and 95% CIs per 1‑SD increase in TyG‑Light‑BA, estimated from Cox models adjusted for baseline covariates excluding the stratifying variable in each subgroup. *P* for interaction from pooled likelihood ratio or Wald tests across multiply imputed datasets (MICE).

*Abbreviations:* HR, hazard ratio; CI, confidence interval; TyG, triglyceride‑glucose index; Light‑BA, Light biological age; BMI, body mass index; CKM, cardiovascular‑kidney‑metabolic.

**Supplementary Table 7. Subgroup analysis of TyG-KDM-BA per SD increase for incident cardiometabolic multimorbidity.**

| **Subgroup** | **Events/Total** | **HR (95% CI)** | ***P* value** | ***P* for interaction** |
| --- | --- | --- | --- | --- |
| **Age, years** |  |  |  | **<0.001** |
| <65 | 618 / 6488 | 1.64 (1.51–1.79) | <0.001 |  |
| ≥65 | 255 / 2279 | 1.25 (1.09–1.44) | 0.002 |  |
| **Sex** |  |  |  | **0.832** |
| Female | 523 / 4680 | 1.50 (1.35–1.66) | <0.001 |  |
| Male | 350 / 4087 | 1.49 (1.31–1.68) | <0.001 |  |
| **BMI, kg/m²** |  |  |  | **0.119** |
| <24 | 365 / 5263 | 1.55 (1.37–1.76) | <0.001 |  |
| ≥24 | 508 / 3504 | 1.43 (1.29–1.58) | <0.001 |  |
| **Residence type** |  |  |  | **0.846** |
| Rural | 707 / 7330 | 1.49 (1.36–1.62) | <0.001 |  |
| Urban | 166 / 1437 | 1.54 (1.28–1.84) | <0.001 |  |
| **Education level** |  |  |  | **0.053** |
| Elementary or below | 630 / 6183 | 1.46 (1.33–1.60) | <0.001 |  |
| Middle school or above | 243 / 2584 | 1.60 (1.38–1.86) | <0.001 |  |
| **Smoking status** |  |  |  | **0.441** |
| Never | 578 / 5379 | 1.54 (1.40–1.69) | <0.001 |  |
| Ever | 295 / 3388 | 1.40 (1.22–1.61) | <0.001 |  |
| **Alcohol consumption** |  |  |  | **0.711** |
| Never | 562 / 5344 | 1.58 (1.44–1.75) | <0.001 |  |
| Ever | 311 / 3423 | 1.37 (1.21–1.56) | <0.001 |  |
| **Hypertension** |  |  |  | **<0.001** |
| No | 336 / 5239 | 1.58 (1.35–1.84) | <0.001 |  |
| Yes | 537 / 3528 | 1.22 (1.10–1.36) | <0.001 |  |
| **Hyperlipidemia** |  |  |  | **0.054** |
| No | 197 / 2937 | 1.79 (1.47–2.17) | <0.001 |  |
| Yes | 676 / 5830 | 1.43 (1.31–1.57) | <0.001 |  |
| **CKM stage** |  |  |  | **0.034** |
| Stage 0–1 | 159 / 4030 | 1.48 (1.13–1.94) | 0.005 |  |
| Stage 2–3 | 433 / 3636 | 1.39 (1.24–1.56) | <0.001 |  |
| Stage 4 | 261 / 896 | 1.24 (1.07–1.45) | 0.005 |  |
| Missing/unclassifiable | 20 / 205 | 1.59 (0.88–2.87) | 0.124 |  |
| *Abbreviations: HR, hazard ratio; CI, confidence interval.* | | | | |
| *Models were adjusted for baseline covariates excluding the stratifying subgroup variable.* | | | | |
| *P for interaction derived from pooled D1/D3 likelihood ratio test or Wald test.* | | | | |
| *Missing data were handled using multiple imputation by chained equations (MICE).* | | | | |

**Supplementary Table 7. Subgroup analysis of TyG-KDM-BA per SD increase for incident cardiometabolic multimorbidity.**

HRs and 95% CIs per 1‑SD increase in TyG‑KDM‑BA, estimated from Cox models adjusted for baseline covariates excluding the stratifying variable in each subgroup. *P* for interaction from pooled likelihood ratio or Wald tests across multiply imputed datasets (MICE).

*Abbreviations:* HR, hazard ratio; CI, confidence interval; TyG, triglyceride‑glucose index; KDM‑BA, Klemera–Doubal biological age; BMI, body mass index; CKM, cardiovascular‑kidney‑metabolic.

**Supplementary Table 8. Effect modification by frailty index on the association of TyG‑BA composites with incident cardiometabolic multimorbidity.**

| **Exposure / Subgroup** | **FI Category** | **sHR (95% CI)** | ***P* value** | ***P* for interaction** |
| --- | --- | --- | --- | --- |
| **TyG-Light-BA (per SD)** | | | | |
|  | Robust (<0.10) | 1.93 (1.49-2.49) | <0.001 | <0.001 |
|  | Pre-frail (0.10-0.25) | 1.54 (1.32-1.78) | <0.001 |  |
|  | Frail (>=0.25) | 1.41 (1.18-1.68) | <0.001 |  |
| **TyG-KDM-BA (per SD)** | | | | |
|  | Robust (<0.10) | 1.71 (1.41-2.07) | <0.001 | <0.001 |
|  | Pre-frail (0.10-0.25) | 1.43 (1.28-1.59) | <0.001 |  |
|  | Frail (>=0.25) | 1.34 (1.17-1.55) | <0.001 |  |
| *Abbreviations: sHR, subdistribution hazard ratio; CI, Confidence Interval; FI, Frailty Index. FI categories: Robust (FI < 0.10), Pre-frail (0.10 ≤ FI < 0.25), Frail (FI ≥ 0.25). Models adjusted for age, sex, education attainment, residence, smoking status, alcohol consumption, waist circumference, HDL-C. P for interaction was derived from a pooled Wald test (D1) comparing nested models with and without the exposure × FI category interaction term. Results were pooled across 5 multiply imputed datasets using Rubin's rules.* | | | | |

**Supplementary Table 8. Effect modification by frailty index on the association of TyG‑BA composites with incident cardiometabolic multimorbidity.**

Subdistribution HRs (sHR) and 95% CIs per 1‑SD increase from Fine‑Gray models treating death as competing risk. FI categories: robust (<0.10), pre‑frail (0.10–0.25), frail (≥0.25). Models adjusted for age, sex, education, residence, smoking, alcohol, waist circumference, and HDL‑C. *P* for interaction from pooled Wald test comparing models with and without exposure × FI category interaction term. Results pooled across 5 multiply imputed datasets.

*Abbreviations:* sHR, subdistribution hazard ratio; CI, confidence interval; FI, frailty index; TyG, triglyceride‑glucose index; Light‑BA, Light biological age; KDM‑BA, Klemera–Doubal biological age; CMM, cardiometabolic multimorbidity.

**Supplementary Table 9. Predefined pairwise comparisons of time‑dependent AUCs with Benjamini–Hochberg false discovery rate correction.**

| Time (years) | Marker A | Marker B | Raw *P*-value | BH-FDR adjusted *P* |
| --- | --- | --- | --- | --- |
| 3 | TyG-Light-BA | TyG | 0.179 | 0.607 |
|  | TyG-KDM-BA | TyG | 0.715 | 0.814 |
|  | TyG-Light-BA | TyG-BMI | 0.258 | 0.646 |
|  | TyG-Light-BA | TyG-WC | 0.182 | 0.607 |
|  | TyG-Light-BA | TyG-WHtR | 0.601 | 0.814 |
|  | TyG-Light-BA | TyG-CVAI | 0.125 | 0.607 |
|  | TyG-KDM-BA | TyG-BMI | 0.733 | 0.814 |
|  | TyG-KDM-BA | TyG-WC | 0.724 | 0.814 |
|  | TyG-KDM-BA | TyG-WHtR | 0.897 | 0.897 |
|  | TyG-KDM-BA | TyG-CVAI | 0.462 | 0.814 |
| 5 | TyG-Light-BA | TyG | 0.167 | 0.464 |
|  | TyG-KDM-BA | TyG | 0.490 | 0.582 |
|  | TyG-Light-BA | TyG-BMI | 0.126 | 0.464 |
|  | TyG-Light-BA | TyG-WC | 0.186 | 0.464 |
|  | TyG-Light-BA | TyG-WHtR | 0.299 | 0.523 |
|  | TyG-Light-BA | TyG-CVAI | 0.155 | 0.464 |
|  | TyG-KDM-BA | TyG-BMI | 0.315 | 0.523 |
|  | TyG-KDM-BA | TyG-WC | 0.524 | 0.582 |
|  | TyG-KDM-BA | TyG-WHtR | 0.622 | 0.622 |
|  | TyG-KDM-BA | TyG-CVAI | 0.366 | 0.523 |
| 8 | TyG-Light-BA | TyG | 0.121 | 0.152 |
|  | TyG-KDM-BA | TyG | 0.002 | 0.007 |
|  | TyG-Light-BA | TyG-BMI | 0.672 | 0.672 |
|  | TyG-Light-BA | TyG-WC | 0.096 | 0.137 |
|  | TyG-Light-BA | TyG-WHtR | 0.072 | 0.119 |
|  | TyG-Light-BA | TyG-CVAI | 0.145 | 0.161 |
|  | TyG-KDM-BA | TyG-BMI | 0.059 | 0.117 |
|  | TyG-KDM-BA | TyG-WC | 0.001 | 0.007 |
|  | TyG-KDM-BA | TyG-WHtR | <0.001 | 0.007 |
|  | TyG-KDM-BA | TyG-CVAI | 0.003 | 0.007 |
| *Comparison of time-dependent AUCs using timeROC::compare() with iid-based variance estimator; Models: Model 3.* | | | | |
| *Comparisons are restricted to pre-specified pairs. BH-FDR correction was performed within each time point across all planned tests.* | | | | |

**Supplementary Table 9. Predefined pairwise comparisons of time‑dependent AUCs with Benjamini–Hochberg false discovery rate correction.**

Pairwise comparisons of time‑dependent AUCs at 3‑, 5‑, and 8‑year horizons using the iid‑based variance estimator (timeROC). All models included Model 3 covariates. BH‑FDR correction was applied within each time point across all planned comparisons.

*Abbreviations:* AUC, area under the curve; BH‑FDR, Benjamini–Hochberg false discovery rate; TyG, triglyceride‑glucose index; Light‑BA, Light biological age; KDM‑BA, Klemera–Doubal biological age; BMI, body mass index; WC, waist circumference; WHtR, waist‑to‑height ratio; CVAI, Chinese visceral adiposity index.

**Supplementary Table 10. Sensitivity analysis of pooled ΔC‑index at 60, 96, and 108 months.**

| **Model Comparison** | **ΔC-index 60months** | | **ΔC-index 96months** | | **ΔC-index 108months** | |
| --- | --- | --- | --- | --- | --- | --- |
|  | **Estimate (95% CI)** | ***P* Value** | **Estimate (95% CI)** | ***P* Value** | **Estimate (95% CI)** | ***P* Value** |
| Base + KDMBA vs. Base Model | 0.007 (0.007 to 0.007) | <0.001 | 0.021 (0.020 to 0.021) | <0.001 | 0.019 (0.019 to 0.020) | <0.001 |
| Base + LightBA vs. Base Model | 0.010 (0.010 to 0.010) | <0.001 | 0.011 (0.010 to 0.011) | <0.001 | 0.009 (0.009 to 0.010) | <0.001 |
| Base + TyG vs. Base Model | 0.005 (0.005 to 0.005) | <0.001 | 0.009 (0.009 to 0.009) | <0.001 | 0.011 (0.011 to 0.012) | <0.001 |
| Base + TyG-KDMBA vs. Base Model | 0.009 (0.009 to 0.009) | <0.001 | 0.024 (0.023 to 0.024) | <0.001 | 0.023 (0.023 to 0.023) | <0.001 |
| Base + TyG-KDMBA vs. Base + TyG | 0.004 (0.004 to 0.004) | <0.001 | 0.015 (0.014 to 0.015) | <0.001 | 0.012 (0.011 to 0.012) | <0.001 |
| Base + TyG-KDMBA vs. Base + TyG + KDMBA | -0.001 (-0.001 to -0.001) | <0.001 | -0.000 (-0.001 to -0.000) | 0.021 | -0.001 (-0.002 to -0.001) | <0.001 |
| Base + TyG-LightBA vs. Base Model | 0.013 (0.012 to 0.013) | <0.001 | 0.015 (0.015 to 0.016) | <0.001 | 0.015 (0.015 to 0.016) | <0.001 |
| Base + TyG-LightBA vs. Base + TyG | 0.008 (0.008 to 0.008) | <0.001 | 0.006 (0.006 to 0.007) | <0.001 | 0.004 (0.004 to 0.004) | <0.001 |
| Base + TyG-LightBA vs. Base + TyG + LightBA | 0.001 (0.000 to 0.001) | 0.004 | -0.001 (-0.001 to -0.001) | <0.001 | -0.002 (-0.003 to -0.002) | <0.001 |

**Supplementary Table 10. Sensitivity analysis of pooled ΔC‑index at 60, 96, and 108 months.**

IPCW‑adjusted ΔC‑index (95% CI) comparing nested models at 60‑, 96‑, and 108‑month horizons. Base model: age, sex, education, residence, smoking, alcohol, waist circumference, HDL‑C. Results pooled across 5 multiply imputed datasets.

*Abbreviations:* CI, confidence interval; IPCW, inverse probability of censoring weighting; TyG, triglyceride‑glucose index; Light‑BA, Light biological age; KDM‑BA, Klemera–Doubal biological age.

**Supplementary Table 11. Fine‑Gray competing risk analysis of TyG‑BA indices for incident CMM (baseline cohort).**

| Characteristic | Crude Model | | Model 1 | | Model 2 | | Model 3 | |
| --- | --- | --- | --- | --- | --- | --- | --- | --- |
|  | sHR (95% CI) | *P* value | sHR (95% CI) | *P* value | sHR (95% CI) | *P* value | sHR (95% CI) | *P* value |
| **Single baseline continuous biomarkers (per SD)** | | | | | | | | |
| TyG index (per SD) | 1.46 (1.38-1.54) | <0.001 | 1.46 (1.38-1.54) | <0.001 | 1.46 (1.37-1.54) | <0.001 | 1.32 (1.23-1.43) | <0.001 |
| Light biological age (per SD) | 1.33 (1.26-1.41) | <0.001 | 1.73 (1.56-1.92) | <0.001 | 1.72 (1.55-1.91) | <0.001 | 1.48 (1.32-1.66) | <0.001 |
| KDM biological age (per SD) | 1.45 (1.37-1.53) | <0.001 | 1.61 (1.48-1.74) | <0.001 | 1.60 (1.48-1.73) | <0.001 | 1.43 (1.31-1.56) | <0.001 |
| **Combined TyG-biological age index (per SD)** | | | | | | | | |
| TyG-LightBA (per SD) | 1.45 (1.37-1.53) | <0.001 | 1.83 (1.68-2.00) | <0.001 | 1.82 (1.67-1.99) | <0.001 | 1.56 (1.41-1.73) | <0.001 |
| TyG-KDMBA (per SD) | 1.53 (1.45-1.62) | <0.001 | 1.65 (1.54-1.77) | <0.001 | 1.65 (1.54-1.77) | <0.001 | 1.45 (1.34-1.56) | <0.001 |
| **TyG-Light-BA Quartiles** | | | | | | | | |
| Q1 | Ref |  | Ref |  | Ref |  | Ref |  |
| Q2 | 1.79 (1.41-2.28) | <0.001 | 2.17 (1.69-2.78) | <0.001 | 2.16 (1.69-2.78) | <0.001 | 1.85 (1.44-2.38) | <0.001 |
| Q3 | 2.77 (2.21-3.47) | <0.001 | 3.95 (3.07-5.10) | <0.001 | 3.95 (3.06-5.09) | <0.001 | 2.93 (2.25-3.83) | <0.001 |
| Q4 | 3.23 (2.59-4.03) | <0.001 | 5.56 (4.17-7.41) | <0.001 | 5.48 (4.11-7.31) | <0.001 | 3.55 (2.59-4.86) | <0.001 |
| **TyG-KDM-BA Quartiles** | | | | | | | | |
| Q1 | Ref |  | Ref |  | Ref |  | Ref |  |
| Q2 | 1.69 (1.32-2.17) | <0.001 | 1.90 (1.47-2.46) | <0.001 | 1.89 (1.46-2.44) | <0.001 | 1.68 (1.30-2.17) | <0.001 |
| Q3 | 2.90 (2.31-3.65) | <0.001 | 3.45 (2.70-4.41) | <0.001 | 3.45 (2.70-4.40) | <0.001 | 2.73 (2.12-3.51) | <0.001 |
| Q4 | 3.70 (2.96-4.62) | <0.001 | 4.69 (3.62-6.08) | <0.001 | 4.68 (3.61-6.08) | <0.001 | 3.27 (2.48-4.33) | <0.001 |
| *Abbreviations: sHR, subdistribution hazard ratio; CI, Confidence Interval; TyG, Triglyceride-Glucose index; BA, Biological Age; Ref, Reference. Model 1: Adjusted for baseline age, sex, education attainment, and residence. Model 2: Model 1 + smoking status and alcohol consumption. Model 3: Model 2 + SBP, waist circumference, HDL-C, platelet count, BUN, and frailty index (FI). Note: Fine-Gray subdistribution hazard models with all-cause mortality as a competing risk. Results pooled across 5 multiply imputed datasets using Rubin's rules.* | | | | | | | | |

**Supplementary Table 11. Fine‑Gray competing risk analysis of TyG‑BA indices for incident CMM (baseline cohort).**

Subdistribution HRs (sHR) and 95% CIs per 1‑SD increase or versus Q1, from Fine‑Gray models treating all‑cause mortality as a competing risk. Model 1: age, sex, education, residence. Model 2: Model 1 + smoking, alcohol. Model 3: Model 2 + SBP, waist circumference, HDL‑C. Results pooled across 5 multiply imputed datasets.

*Abbreviations:* sHR, subdistribution hazard ratio; CI, confidence interval; TyG, triglyceride‑glucose index; Light‑BA, Light biological age; KDM‑BA, Klemera–Doubal biological age; CMM, cardiometabolic multimorbidity; Ref, reference.

**Supplementary Table 12. Fine‑Gray competing risk analysis of joint TyG and biological age acceleration groups for incident CMM (baseline cohort).**

| Characteristic | Crude Model | | Model 1 | | Model 2 | | Model 3 | |
| --- | --- | --- | --- | --- | --- | --- | --- | --- |
|  | sHR (95% CI) | *P* value | sHR (95% CI) | *P* value | sHR (95% CI) | *P* value | sHR (95% CI) | *P* value |
| **TyG Status** | | | | | | | | |
| Low TyG | Ref |  | Ref |  | Ref |  | Ref |  |
| High TyG | 2.02 (1.76-2.32) | <0.001 | 1.96 (1.70-2.25) | <0.001 | 1.95 (1.69-2.24) | <0.001 | 1.52 (1.30-1.78) | <0.001 |
| **Light-BA Status** | | | | | | | | |
| Non-accelerated Aging | Ref |  | Ref |  | Ref |  | Ref |  |
| Accelerated Aging | 1.93 (1.69-2.21) | <0.001 | 1.91 (1.67-2.18) | <0.001 | 1.90 (1.66-2.17) | <0.001 | 1.56 (1.36-1.79) | <0.001 |
| **Joint Group (TyG & Light-BA)** | | | | | | | | |
| Low TyG & Non-accelerated Aging | Ref |  | Ref |  | Ref |  | Ref |  |
| Low TyG & Accelerated Aging | 1.81 (1.44-2.28) | <0.001 | 1.81 (1.44-2.27) | <0.001 | 1.80 (1.44-2.27) | <0.001 | 1.63 (1.30-2.05) | <0.001 |
| High TyG & Non-accelerated Aging | 1.90 (1.54-2.35) | <0.001 | 1.85 (1.50-2.28) | <0.001 | 1.84 (1.49-2.27) | <0.001 | 1.56 (1.25-1.95) | <0.001 |
| High TyG & Accelerated Aging | 3.26 (2.70-3.94) | <0.001 | 3.15 (2.60-3.81) | <0.001 | 3.12 (2.58-3.78) | <0.001 | 2.27 (1.83-2.81) | <0.001 |
| **KDM-BA Status** | | | | | | | | |
| Non-accelerated Aging | Ref |  | Ref |  | Ref |  | Ref |  |
| Accelerated Aging | 1.83 (1.60-2.09) | <0.001 | 1.82 (1.59-2.08) | <0.001 | 1.82 (1.58-2.08) | <0.001 | 1.55 (1.34-1.78) | <0.001 |
| **Joint Group (TyG & KDM-BA)** | | | | | | | | |
| Low TyG & Non-accelerated Aging | Ref |  | Ref |  | Ref |  | Ref |  |
| Low TyG & Accelerated Aging | 1.72 (1.37-2.17) | <0.001 | 1.73 (1.38-2.17) | <0.001 | 1.73 (1.38-2.17) | <0.001 | 1.56 (1.24-1.97) | <0.001 |
| High TyG & Non-accelerated Aging | 1.93 (1.56-2.39) | <0.001 | 1.87 (1.51-2.31) | <0.001 | 1.86 (1.50-2.31) | <0.001 | 1.51 (1.21-1.89) | <0.001 |
| High TyG & Accelerated Aging | 3.15 (2.60-3.81) | <0.001 | 3.04 (2.51-3.69) | <0.001 | 3.02 (2.49-3.66) | <0.001 | 2.18 (1.77-2.69) | <0.001 |
| *Abbreviations: sHR, subdistribution hazard ratio; CI, Confidence Interval; TyG, Triglyceride-Glucose index; BA, Biological Age; Ref, Reference. Model 1: Adjusted for baseline age, sex, education attainment, and residence. Model 2: Model 1 + smoking status and alcohol consumption. Model 3: Model 2 + waist circumference, HDL-C. Note: Fine-Gray subdistribution hazard models with all-cause mortality as a competing risk. Results pooled across 5 multiply imputed datasets using Rubin's rules.* | | | | | | | | |

**Supplementary Table 12. Fine‑Gray competing risk analysis of joint TyG and biological age acceleration groups for incident CMM (baseline cohort).**

sHRs and 95% CIs from Fine‑Gray models treating all‑cause mortality as a competing risk. TyG status: high vs. low (median cutoff 8.67); BAA: accelerated (>0 years) vs. non‑accelerated (≤0). Reference: Low TyG & Non‑accelerated Aging. Models adjusted as described in Supplementary Table 11 (Model 1–3). Results pooled across 5 multiply imputed datasets.

*Abbreviations:* sHR, subdistribution hazard ratio; CI, confidence interval; TyG, triglyceride‑glucose index; Light‑BA, Light biological age; KDM‑BA, Klemera–Doubal biological age; BAA, biological age acceleration; CMM, cardiometabolic multimorbidity; Ref, reference.

**Supplementary Table 13. Fine‑Gray competing risk analysis of cumulative and trajectory‑based TyG‑BA indices for incident CMM (Wave 3 landmark subcohort).**

| Characteristic | Crude Model | | Model 1 | | Model 2 | | Model 3 | |
| --- | --- | --- | --- | --- | --- | --- | --- | --- |
|  | sHR (95% CI) | P value | sHR (95% CI) | P value | sHR (95% CI) | P value | sHR (95% CI) | P value |
| **TyG-Light-BA Wave3 (per SD)** | 1.47 (1.35-1.61) | <0.001 | 1.73 (1.53-1.97) | <0.001 | 1.73 (1.52-1.97) | <0.001 | 1.53 (1.32-1.78) | <0.001 |
| **TyG-KDM-BA Wave3 (per SD)** | 1.63 (1.50-1.78) | <0.001 | 1.74 (1.58-1.92) | <0.001 | 1.74 (1.57-1.91) | <0.001 | 1.59 (1.42-1.77) | <0.001 |
| **Cumulative TyG-Light-BA (per SD)** | 1.55 (1.42-1.69) | <0.001 | 2.15 (1.88-2.46) | <0.001 | 2.14 (1.87-2.45) | <0.001 | 1.92 (1.64-2.25) | <0.001 |
| **Cumulative TyG-KDM-BA (per SD)** | 1.72 (1.59-1.87) | <0.001 | 1.95 (1.76-2.15) | <0.001 | 1.94 (1.76-2.14) | <0.001 | 1.78 (1.59-1.99) | <0.001 |
| **Cumulative TyG-Light-BA Quartiles** | | | | | | | | |
| Q1 | Ref |  | Ref |  | Ref |  | Ref |  |
| Q2 | 1.43 (1.03-1.99) | 0.034 | 1.86 (1.32-2.62) | <0.001 | 1.85 (1.31-2.61) | <0.001 | 1.67 (1.18-2.36) | 0.004 |
| Q3 | 2.31 (1.71-3.13) | <0.001 | 3.76 (2.65-5.34) | <0.001 | 3.72 (2.62-5.28) | <0.001 | 3.02 (2.09-4.37) | <0.001 |
| Q4 | 3.35 (2.50-4.50) | <0.001 | 7.01 (4.69-10.47) | <0.001 | 6.88 (4.59-10.31) | <0.001 | 5.11 (3.30-7.92) | <0.001 |
| **Cumulative TyG-KDM-BA Quartiles** | | | | | | | | |
| Q1 | Ref |  | Ref |  | Ref |  | Ref |  |
| Q2 | 1.43 (1.01-2.04) | 0.047 | 1.68 (1.17-2.40) | 0.005 | 1.66 (1.16-2.38) | 0.006 | 1.51 (1.05-2.17) | 0.024 |
| Q3 | 2.52 (1.83-3.48) | <0.001 | 3.22 (2.29-4.53) | <0.001 | 3.18 (2.26-4.47) | <0.001 | 2.71 (1.90-3.87) | <0.001 |
| Q4 | 4.39 (3.24-5.95) | <0.001 | 6.13 (4.34-8.67) | <0.001 | 6.05 (4.27-8.56) | <0.001 | 4.70 (3.23-6.84) | <0.001 |
| **TyG-Light-BA Trajectory Clusters** | | | | | | | | |
| 1 Low | Ref |  | Ref |  | Ref |  | Ref |  |
| 2 Moderate | 1.94 (1.52-2.47) | <0.001 | 2.63 (2.00-3.47) | <0.001 | 2.61 (1.98-3.44) | <0.001 | 2.19 (1.65-2.91) | <0.001 |
| 3 High | 3.09 (2.37-4.02) | <0.001 | 5.44 (3.71-7.97) | <0.001 | 5.38 (3.66-7.91) | <0.001 | 3.89 (2.57-5.87) | <0.001 |
| **TyG-KDMBA Trajectory Clusters** | | | | | | | | |
| 1 Low | Ref |  | Ref |  | Ref |  | Ref |  |
| 2 Moderate | 2.49 (1.91-3.24) | <0.001 | 2.94 (2.23-3.88) | <0.001 | 2.91 (2.20-3.85) | <0.001 | 2.56 (1.92-3.41) | <0.001 |
| 3 High | 4.41 (3.34-5.81) | <0.001 | 5.75 (4.17-7.93) | <0.001 | 5.73 (4.15-7.90) | <0.001 | 4.47 (3.16-6.33) | <0.001 |
| *Abbreviations: sHR, subdistribution hazard ratio; CI, Confidence Interval; TyG, Triglyceride-Glucose index; BA, Biological Age; Ref, Reference. Model 1: Adjusted for Wave 3 age, sex, education attainment, and residence. Model 2: Model 1 + smoking status and alcohol consumption. Model 3: Model 2 + Wave 3waist circumference, HDL-C. Note: Fine-Gray subdistribution hazard models with all-cause mortality as a competing risk. Results pooled across 5 multiply imputed datasets using Rubin's rules.* | | | | | | | | |

**Supplementary Table 13. Fine‑Gray competing risk analysis of cumulative and trajectory‑based TyG‑BA indices for incident CMM (Wave 3 landmark subcohort).**

sHRs and 95% CIs from Fine‑Gray models treating all‑cause mortality as a competing risk. Trajectory clusters derived from K‑means (k = 3). Models adjusted as described in Supplementary Table 11, using Wave 3 covariates. Results pooled across 5 multiply imputed datasets.

*Abbreviations:* sHR, subdistribution hazard ratio; CI, confidence interval; TyG, triglyceride‑glucose index; Light‑BA, Light biological age; KDM‑BA, Klemera–Doubal biological age; CMM, cardiometabolic multimorbidity; Ref, reference.

**Supplementary Table 14. Sensitivity analyses of TyG‑BA composite indices for incident cardiometabolic multimorbidity under alternative analytical scenarios.**

| **Sensitivity Analysis Scenario** | **HR (95% CI)** | ***P* value** |
| --- | --- | --- |
| **Exposure: TyG-Light-BA (per SD)** | | |
| Main analysis | 1.62 (1.46–1.80) | <0.001 |
| No adjustment for age | 1.46 (1.36–1.56) | <0.001 |
| Excluding CMM events within first 2 years | 1.58 (1.42–1.76) | <0.001 |
| Complete-case analysis (no multiple imputation) | 1.62 (1.46–1.80) | <0.001 |
| Excluding participants with CKD at baseline | 1.62 (1.46–1.80) | <0.001 |
| Excluding participants with hypertension at baseline | 1.63 (1.37–1.93) | <0.001 |
| Excluding baseline medication users | 1.69 (1.45–1.96) | <0.001 |
| Additional adjustment for medication use | 1.48 (1.33–1.65) | <0.001 |
| Replacing waist circumference with BMI in model | 1.65 (1.49–1.83) | <0.001 |
| CMM definition without HbA1c criterion | 1.65 (1.49–1.84) | <0.001 |
| Follow-up truncated at Wave 3 (2015) | 1.50 (1.26–1.78) | <0.001 |
| Primary Model 3 + frailty index (FI) | 1.56 (1.41–1.73) | <0.001 |
| Primary Model 3 + SBP/PLT/BUN | 1.59 (1.43–1.77) | <0.001 |
| Full Model 3 (primary + FI + SBP/PLT/BUN) | 1.55 (1.40–1.72) | <0.001 |
| Primary Model 3 + uric acid (UA) | 1.64 (1.48–1.83) | <0.001 |
| Primary Model 3 + eGFR | 1.64 (1.47–1.82) | <0.001 |
| Primary Model 3 + UA + eGFR | 1.65 (1.48–1.83) | <0.001 |
| **Exposure: TyG-KDM-BA (per SD)** | | |
| Main analysis | 1.49 (1.38–1.61) | <0.001 |
| No adjustment for age | 1.49 (1.40–1.60) | <0.001 |
| Excluding CMM events within first 2 years | 1.47 (1.36–1.60) | <0.001 |
| Complete-case analysis (no multiple imputation) | 1.49 (1.38–1.61) | <0.001 |
| Excluding participants with CKD at baseline | 1.49 (1.38–1.61) | <0.001 |
| Excluding participants with hypertension at baseline | 1.58 (1.35–1.84) | <0.001 |
| Excluding baseline medication users | 1.46 (1.30–1.64) | <0.001 |
| Additional adjustment for medication use | 1.36 (1.25–1.48) | <0.001 |
| Replacing waist circumference with BMI in model | 1.51 (1.40–1.63) | <0.001 |
| CMM definition without HbA1c criterion | 1.52 (1.41–1.65) | <0.001 |
| Follow-up truncated at Wave 3 (2015) | 1.26 (1.10–1.44) | <0.001 |
| Primary Model 3 + frailty index (FI) | 1.48 (1.37–1.60) | <0.001 |
| Primary Model 3 + SBP/PLT/BUN | 1.86 (1.66–2.09) | <0.001 |
| Full Model 3 (primary + FI + SBP/PLT/BUN) | 1.82 (1.63–2.04) | <0.001 |
| Primary Model 3 + uric acid (UA) | 1.52 (1.41–1.65) | <0.001 |
| Primary Model 3 + eGFR | 1.52 (1.40–1.65) | <0.001 |
| Primary Model 3 + UA + eGFR | 1.54 (1.41–1.67) | <0.001 |
| Abbreviations: HR, Hazard Ratio; CI, Confidence Interval; CKD, Chronic Kidney Disease; HTN, Hypertension; HbA1c, Hemoglobin A1c; SD, Standard Deviation. All models adjusted for primary Model 3 covariates (age, sex, education, residence, smoking, alcohol, waist circumference, and HDL-C) unless specified otherwise in scenario definition. Results were pooled across multiply imputed datasets using Rubin's rules, except for complete case analysis. The 'Follow-up truncated at 2015' scenario restricts CMM ascertainment to Waves 1-3 (where diabetes uses laboratory criteria at W1/W3) to address wave-dependent outcome definition. Adjustment-set scenarios ('+FI', '+SBP/PLT/BUN', 'full Model 3') test whether adding frailty and/or KDM-BA input components (SBP, BUN) changes the primary-model estimates. Renal-metabolic scenarios ('+UA', '+eGFR', '+UA+eGFR') test additional adjustment; eGFR is derived from creatinine, a KDM-BA input, so it is reported as sensitivity only. Interval-censored Weibull models (survreg, interval2) are reported separately as time ratios. | | |

**Supplementary Table 14. Sensitivity analyses of TyG‑BA composite indices for incident cardiometabolic multimorbidity under alternative analytical scenarios.**

HRs and 95% CIs per 1‑SD increase, estimated from Cox models. All scenarios adjusted for primary Model 3 covariates (age, sex, education, residence, smoking, alcohol, waist circumference, HDL‑C) unless otherwise specified. Results pooled across multiply imputed datasets, except for complete‑case analysis.

*Abbreviations:* HR, hazard ratio; CI, confidence interval; TyG, triglyceride‑glucose index; Light‑BA, Light biological age; KDM‑BA, Klemera–Doubal biological age; CMM, cardiometabolic multimorbidity; CKD, chronic kidney disease; SBP, systolic blood pressure; PLT, platelet count; BUN, blood urea nitrogen; UA, uric acid; eGFR, estimated glomerular filtration rate; FI, frailty index; BMI, body mass index; HbA1c, glycated hemoglobin.

**Supplementary Table 15. Interval‑censored Weibull survival models for incident cardiometabolic multimorbidity.**

| Exposure (per SD) | Time ratio (95% CI) | *P* value |
| --- | --- | --- |
| TyG-Light-BA (per SD) | 0.74 (0.69–0.79) | <0.001 |
| TyG-KDM-BA (per SD) | 0.78 (0.74–0.82) | <0.001 |
| Event time is treated as an interval between the last CMM-free wave and the first wave with CMM. | | |
| Non-events are right-censored. Time ratio > 1 indicates longer event-free time (lower hazard); | | |
| models are adjusted for primary Model 3 covariates and pooled across imputations by Rubin's rules. | | |

**Supplementary Table 15. Interval‑censored Weibull survival models for incident cardiometabolic multimorbidity.**

Time ratios (95% CI) per 1‑SD increase, estimated from interval‑censored Weibull models. Event time treated as interval between last CMM‑free wave and first wave with CMM; non‑events right‑censored. Time ratio < 1 indicates shorter event‑free time (higher hazard). Models adjusted for primary Model 3 covariates and pooled across multiply imputed datasets.

*Abbreviations:* CI, confidence interval; TyG, triglyceride‑glucose index; Light‑BA, Light biological age; KDM‑BA, Klemera–Doubal biological age; CMM, cardiometabolic multimorbidity.

**Supplementary Table 16. Associations of TyG‑BA composites with incident cardiometabolic multimorbidity with and without chronological age adjustment.**

| Exposure | Model A: No chronological age | | Model B: Age‑adjusted | |
| --- | --- | --- | --- | --- |
|  | HR (95% CI) | *P* value | HR (95% CI) | *P* value |
| **TyG‑Light‑BA composite (per SD)** | | | | |
| TyG‑Light‑BA composite (per SD) | 1.46 (1.36, 1.56) | < 0.001 | 1.62 (1.46, 1.80) | < 0.001 |
| **TyG‑Light‑BA composite (quartiles)** | | | | |
| Q1 (reference) | Ref |  | Ref |  |
| Q2 | 1.74 (1.36, 2.22) | < 0.001 | 1.81 (1.41, 2.33) | < 0.001 |
| Q3 | 2.63 (2.08, 3.33) | < 0.001 | 2.86 (2.20, 3.72) | < 0.001 |
| Q4 | 3.12 (2.47, 3.95) | < 0.001 | 3.58 (2.64, 4.87) | < 0.001 |
| **TyG‑KDM‑BA composite (per SD)** | | | | |
| TyG‑KDM‑BA composite (per SD) | 1.49 (1.40, 1.60) | < 0.001 | 1.49 (1.38, 1.61) | < 0.001 |
| **TyG‑KDM‑BA composite (quartiles)** | | | | |
| Q1 (reference) | Ref |  | Ref |  |
| Q2 | 1.68 (1.30, 2.16) | < 0.001 | 1.66 (1.28, 2.14) | < 0.001 |
| Q3 | 2.73 (2.15, 3.46) | < 0.001 | 2.68 (2.09, 3.43) | < 0.001 |
| Q4 | 3.45 (2.72, 4.36) | < 0.001 | 3.35 (2.57, 4.36) | < 0.001 |
| *Abbreviations: HR, Hazard Ratio; CI, Confidence Interval; Ref, Reference; TyG, Triglyceride‑Glucose index; BA, Biological Age. Model A (no chronological age): adjusted for sex, educational attainment, residence, smoking status, alcohol consumption, waist circumference, and HDL‑C. Model B (age‑adjusted): Model A plus chronological age at baseline. Analyses were performed on multiply imputed datasets and pooled using Rubin's rules. Quartiles were derived from the distribution of each TyG‑BA composite index; Q1 is set as the reference group.*  **Supplementary Table 16. Associations of TyG‑BA composites with incident cardiometabolic multimorbidity with and without chronological age adjustment.**  HRs and 95% CIs per 1‑SD increase or versus Q1. Model A (no chronological age): adjusted for sex, education, residence, smoking, alcohol, waist circumference, HDL‑C. Model B (age‑adjusted): Model A plus chronological age. Quartiles derived from the distribution of each composite index. Results pooled across multiply imputed datasets.  *Abbreviations:* HR, hazard ratio; CI, confidence interval; TyG, triglyceride‑glucose index; Light‑BA, Light biological age; KDM‑BA, Klemera–Doubal biological age; CMM, cardiometabolic multimorbidity; Ref, reference. | | | | |
