## Supplementary material for "Joint contributions of metabolic dysfunction and biological aging to cardiometabolic multimorbidity and disease progression: a prospective cohort study": N/A

**Supplementary Methods**

**Part 1. Baseline characteristics of included versus excluded participants and sensitivity analyses for missing baseline biomarkers**

Demographic, lifestyle, anthropometric and frailty-related baseline variables were compared between participants included in the main analytical cohort and participants whose only exclusion criterion was missing baseline biomarkers required for TyG, Light-BA, or KDM-BA. Because biomarker values were missing by definition in the excluded subgroup, only non-biomarker baseline characteristics were compared (Table S1). Stabilized inverse probability weights were additionally estimated from a logistic propensity model based on the same non-biomarker characteristics to evaluate the potential impact of selection bias on baseline comparability (Table S2). As a further sensitivity analysis, missing baseline exposure biomarkers were multiply imputed, and the fully adjusted Model 3 was re-estimated in the expanded cohort of otherwise eligible participants (Table S3).

| Characteristic | Included (N = 8,767) | Excluded (biomarker missing only) (N = 6,565) | *P* value |
| --- | --- | --- | --- |
| Age (years), Mean ± SD | 58.97 ± 9.21 | 58.25 ± 9.84 | <0.001 |
| Sex, n (%) |  |  | <0.001 |
| Female | 4,680.0 (53.4%) | 3,186.0 (48.5%) |  |
| Male | 4,087.0 (46.6%) | 3,379.0 (51.5%) |  |
| Education level, n (%) |  |  | <0.001 |
| Elementary school or below | 6,183.0 (70.5%) | 4,042.0 (61.6%) |  |
| Middle school | 2,336.0 (26.6%) | 2,118.0 (32.3%) |  |
| College or above | 248.0 (2.8%) | 405.0 (6.2%) |  |
| Residence type, n (%) |  |  | <0.001 |
| Rural | 7,327.0 (83.6%) | 4,802.0 (73.1%) |  |
| Urban | 1,436.0 (16.4%) | 1,741.0 (26.5%) |  |
| Smoking status, n (%) |  |  | 0.010 |
| Current | 2,644.0 (30.2%) | 1,734.0 (26.4%) |  |
| Former | 744.0 (8.5%) | 448.0 (6.8%) |  |
| Never | 5,349.0 (61.0%) | 3,791.0 (57.7%) |  |
| Drinking status, n (%) |  |  | 0.027 |
| Current | 2,716.0 (31.0%) | 2,135.0 (32.5%) |  |
| Former | 709.0 (8.1%) | 488.0 (7.4%) |  |
| Never | 5,333.0 (60.8%) | 3,852.0 (58.7%) |  |
| BMI (kg/m2), Mean ± SD | 23.44 ± 3.82 | 23.34 ± 4.05 | 0.245 |
| DBP (mmHg), Mean ± SD | 75.28 ± 12.18 | 75.59 ± 12.07 | 0.209 |
| Frailty Index, Mean ± SD | 0.15 ± 0.11 | 0.14 ± 0.12 | <0.001 |

***Table S1. Baseline characteristics between included participants and those excluded only due to missing biomarkers***

Note: The excluded group is restricted to participants whose only exclusion criterion was missing baseline biomarkers required for TyG, Light-BA, or KDM-BA (E2/E3/E3b); participants excluded for age, prevalent CMM, or complete loss to follow-up were not included in this comparison. Because biomarker values are missing by definition in the excluded subgroup, only non-biomarker baseline characteristics were compared. Continuous variables are presented as mean ± SD (two decimal places). Categorical variables are displayed as n (percentage, one decimal place). Two-sample t-test was applied for continuous variables; the Pearson χ² test was used for categorical variables. P-values < 0.001 are shown as '<0.001'; remaining P-values retain three decimal digits. P-values are shown only on the first line of each characteristic for visual simplicity.

| Characteristic | Included, IPW (N = 8,767) | Excluded (biomarker missing only), IPW (N = 6,565) | *P* value |
| --- | --- | --- | --- |
| Age (years), Weighted mean ± SD | 58.73 ± 9.17 | 58.76 ± 10.01 | 0.831 |
| Sex, Weighted n (%) |  |  | 0.945 |
| Female | 4502.0 (51.5%) | 3391.0 (51.6%) |  |
| Male | 4240.3 (48.5%) | 3185.0 (48.4%) |  |
| Education level, Weighted n (%) |  |  | 0.797 |
| Elementary school or below | 5849.3 (66.9%) | 4395.1 (66.8%) |  |
| Middle school | 2533.0 (29.0%) | 1895.8 (28.8%) |  |
| College or above | 360.0 (4.1%) | 285.1 (4.3%) |  |
| Residence type, Weighted n (%) |  |  | 0.750 |
| Rural | 6932.2 (79.3%) | 5183.9 (79.1%) |  |
| Urban | 1806.1 (20.7%) | 1369.1 (20.9%) |  |
| Smoking status, Weighted n (%) |  |  | <0.001 |
| Current | 2513.4 (28.9%) | 1900.6 (31.2%) |  |
| Former | 684.7 (7.9%) | 517.8 (8.5%) |  |
| Never | 5510.6 (63.3%) | 3667.7 (60.3%) |  |
| Drinking status, Weighted n (%) |  |  | 0.870 |
| Current | 2762.2 (31.6%) | 2077.5 (32.0%) |  |
| Former | 682.1 (7.8%) | 510.0 (7.9%) |  |
| Never | 5288.5 (60.6%) | 3904.2 (60.1%) |  |
| BMI (kg/m2), Weighted mean ± SD | 23.33 ± 3.72 | 23.54 ± 4.31 | 0.008 |
| Diastolic blood pressure (mmHg), Weighted mean ± SD | 75.18 ± 12.15 | 75.73 ± 12.12 | 0.024 |
| Frailty Index, Weighted mean ± SD | 0.15 ± 0.11 | 0.15 ± 0.12 | 0.597 |

***Table S2. IPW-weighted baseline characteristics between included participants and those excluded only due to missing biomarkers***

Note: Stabilized inverse probability weights were estimated from a logistic propensity model including age, sex, education, residence, smoking, drinking, BMI, diastolic blood pressure, and frailty index (median/mode imputation for missing propensity covariates). Weighted means/percentages and weighted tests (linear regression with weights for continuous variables; weighted chi-square test for categorical variables) are presented.

| Exposure (per SD) | Primary Model 3 HR (95% CI) | Exposure-MI HR (95% CI) | *P* value | Expanded N | CMM events |
| --- | --- | --- | --- | --- | --- |
| TyG-Light-BA (per SD) | 1.62 (1.46, 1.80) | 1.71 (1.54, 1.90) | < 0.001 | 15,332 | 1,410 |
| TyG-KDM-BA (per SD) | 1.49 (1.38, 1.61) | 1.55 (1.43, 1.68) | < 0.001 | 15,332 | 1,410 |

***Table S3. Exposure multiple imputation sensitivity: primary Model 3 HRs after imputing missing baseline exposure biomarkers and including participants otherwise eligible except for biomarker missingness***

Note: Missing TyG-Light-BA and TyG-KDM-BA composite exposures were multiply imputed (m = 5) using MICE with Model 3 covariates and CMM outcome as auxiliary predictors. Exposure-MI HRs were pooled by Rubin's rules; exposures were standardized using means and SDs from the primary included cohort after 1%-99% winsorization. Primary Model 3 HRs are reported in Table 4 of the main text.

*Abbreviations:* BMI, body mass index; DBP, diastolic blood pressure; SD, standard deviation; IPW, inverse probability weighting; MICE, multiple imputation by chained equations; MI, multiple imputation; HR, hazard ratio; CI, confidence interval; TyG, triglyceride-glucose index; Light-BA, Light's method biological age; KDM-BA, Klemera-Doubal method biological age; CMM, cardiometabolic multimorbidity.

**Part 2. Multiple imputation for missing data**

Multiple imputation by chained equations (MICE) was performed to handle missing values of covariates, implemented via the mice and parlmice R‑packages. Two separate multiply‑imputed datasets were generated for the baseline main cohort and Wave 3 landmark sub‑cohort, respectively. Variables representing study outcomes, derived composite exposures (including TyG-BA interaction terms, cumulative indices, delta‑change metrics, k‑means clustering labels), participant identifiers and time‑to‑event endpoints were excluded from the imputation model.

Imputation methods were specified according to variable types: predictive mean matching (pmm) for stable continuous biomarkers; normal‑distribution imputation (norm) for skewed continuous variables; logistic regression (logreg) for binary covariates; classification and regression trees (cart) for multi‑categorical predictors. The predictor matrix was constrained using quickpred (minimum correlation threshold = 0.1); self‑predictions were disabled (diagonal set to zero). Given the low proportion of missing data and the consistency between complete-case analysis and multiple imputation results, we generated five imputed datasets with 20 iterations per imputation chain. A random seed of 1234 was set to ensure reproducibility, and parallel computation was adopted to accelerate model convergence.

Missing patterns for all candidate imputed variables are summarized in Table S4. All downstream survival analyses were conducted across imputed datasets and pooled according to Rubin’s rules.

| Study cohort | Covariate | Missing (%) | MICE imputation method |
| --- | --- | --- | --- |
| Main baseline cohort | Age | 0.00 | pmm |
| Main baseline cohort | Sex | 0.00 | cart |
| Main baseline cohort | education.attainment | 0.00 | cart |
| Main baseline cohort | marital_status | 10.96 | cart |
| Main baseline cohort | residence | 0.05 | cart |
| Main baseline cohort | smoke.group | 0.34 | cart |
| Main baseline cohort | alq.group | 0.13 | cart |
| Main baseline cohort | BMI | 1.32 | pmm |
| Main baseline cohort | Waist | 0.57 | pmm |
| Main baseline cohort | HbA1c | 0.00 | pmm |
| Main baseline cohort | Glucose | 0.00 | pmm |
| Main baseline cohort | TC | 0.00 | pmm |
| Main baseline cohort | TG | 0.00 | pmm |
| Main baseline cohort | HDL | 0.02 | pmm |
| Main baseline cohort | LDL | 0.15 | pmm |
| Main baseline cohort | SBP | 0.00 | pmm |
| Main baseline cohort | DBP | 0.02 | pmm |
| Main baseline cohort | BUN | 0.00 | pmm |
| Main baseline cohort | Creatinine | 0.00 | pmm |
| Main baseline cohort | CRP | 0.00 | pmm |
| Main baseline cohort | UA | 0.00 | pmm |
| Main baseline cohort | PLT | 1.93 | norm |
| Main baseline cohort | MCV | 1.92 | norm |
| Main baseline cohort | WBC | 1.97 | norm |
| Main baseline cohort | hypertension | 0.00 | logreg |
| Main baseline cohort | diabetes | 0.00 | logreg |
| Main baseline cohort | hyperlipidemia | 0.00 | logreg |
| Main baseline cohort | Cancer | 0.54 | logreg |
| Main baseline cohort | eGFR | 0.00 | norm |
| Main baseline cohort | FI | 0.43 | norm |
| Wave 3 landmark sub-cohort | Age_wave3 | 0.00 | pmm |
| Wave 3 landmark sub-cohort | Waist_wave3 | 0.56 | pmm |
| Wave 3 landmark sub-cohort | HbA1c_wave3 | 0.00 | pmm |
| Wave 3 landmark sub-cohort | Glucose_wave3 | 0.00 | pmm |
| Wave 3 landmark sub-cohort | TC_wave3 | 0.00 | pmm |
| Wave 3 landmark sub-cohort | TG_wave3 | 0.00 | pmm |
| Wave 3 landmark sub-cohort | HDL_wave3 | 0.00 | pmm |
| Wave 3 landmark sub-cohort | LDL_wave3 | 0.00 | pmm |
| Wave 3 landmark sub-cohort | SBP_wave3 | 0.00 | pmm |
| Wave 3 landmark sub-cohort | DBP_wave3 | 0.00 | pmm |
| Wave 3 landmark sub-cohort | BUN_wave3 | 0.00 | pmm |
| Wave 3 landmark sub-cohort | Creatinine_wave3 | 0.00 | pmm |
| Wave 3 landmark sub-cohort | CRP_wave3 | 0.00 | pmm |
| Wave 3 landmark sub-cohort | UA_wave3 | 0.00 | pmm |
| Wave 3 landmark sub-cohort | PLT_wave3 | 0.89 | norm |
| Wave 3 landmark sub-cohort | FI_wave3 | 0.93 | norm |

***Table S4. Missing-value pattern of covariates subjected to multiple imputation by chained equations (MICE)***

*Abbreviations:* BMI, body mass index; Waist, waist circumference; HbA1c, glycated hemoglobin A1c; Glucose, fasting plasma glucose; TC, total cholesterol; TG, triglycerides; HDL, high‑density lipoprotein cholesterol; LDL, low‑density lipoprotein cholesterol; SBP, systolic blood pressure; DBP, diastolic blood pressure; BUN, blood urea nitrogen; Creatinine, serum creatinine; CRP, C‑reactive protein; UA, uric acid; PLT, platelet count; MCV, mean corpuscular volume; WBC, white blood cell count; eGFR, estimated glomerular filtration rate; FI, frailty index.

**Part 3.** **Operational Definitions of Cardiovascular-Kidney-Metabolic (CKM) Syndrome Stages 0-4 in CHARLS.**

The stages of the cardiovascular-kidney-metabolic (CKM) syndrome were defined according to the updated 2026 American Heart Association/American College of Cardiology/American Diabetes Association/American Society of Nephrology (AHA/ACC/ADA/ASN) CKM syndrome clinical staging framework, with modifications tailored to variable availability in the China Health and Retirement Longitudinal Study (CHARLS) cohort. The estimated glomerular filtration rate (eGFR) was calculated using the race-free 2021 CKD-EPI creatinine equation, which is recommended for contemporary chronic kidney disease risk stratification in Asian populations.

A hierarchical, mutually exclusive five-stage CKM classification system (stage 0 to stage 4) was applied in the present analysis. Notably, several variables required for the full 2026 CKM guideline criteria were unavailable in the CHARLS dataset, including urinary albumin-to-creatinine ratio, cardiac biomarkers, coronary artery calcium imaging, and detailed peripheral artery disease information. Therefore, CKM staging in the current study was simplified and adapted exclusively to accessible anthropometric, laboratory, and self-reported clinical variables. Participants with missing or unclassifiable CKM stage (n = 205) were retained in the analytic cohort and analyzed as a separate category in CKM subgroup analyses. Because CKM stage was used for baseline characterization and subgroup analyses rather than as an eligibility criterion for the primary analytical cohort, these participants were not excluded. Detailed operational definitions for each CKM stage are provided in the Table S5.

| **CKM Stage** | **Definition** | **Operational Definition in CHARLS** |
| --- | --- | --- |
| **Stage 0**  (No CKM risk factors) | Individuals with normal adiposity, absent cardiometabolic/renal risk factors, and no criteria for other CKM stages. | Asian participants with BMI <23 kg/m² and normal waist circumference (<80 cm for women, <90 cm for men), not satisfying diagnostic criteria for Stages 1-4. |
| **Stage 1**  (Excess dysfunctional adiposity) | Individuals with overweight/obesity, abdominal obesity, or prediabetes, without additional metabolic risk factors or CKD. | BMI ≥23 kg/m², or waist circumference ≥80 cm (women)/≥90 cm (men), or prediabetes (fasting blood glucose 100–125 mg/dL or HbA1c 5.7%–6.4%); exclusion of hypertension, diabetes, and eGFR <60 mL/min/1.73m². |
| **Stage 2**  **(**Metabolic risk factors and CKD) | Individuals with metabolic risk factors or moderate-to-high-risk CKD, in the absence of subclinical or clinical CVD. | Presence of hypertension or diabetes, or moderate CKD (eGFR 30–59 mL/min/1.73m²). Note: Urinary albumin-to-creatinine ratio unavailable in CHARLS, thus moderate-high-risk CKD was defined solely by eGFR. |
| **Stage 3**  (High-risk pre-clinical CKM) | Individuals with subclinical CVD or very-high-risk KDIGO CKD (risk equivalent status). | Two mutually eligible criteria: (1) Very-high-risk CKD (eGFR <30 mL/min/1.73m²); (2) 10-year total CVD risk ≥20% estimated via PREVENT risk equations. |
| **Stage 4**  (Clinical CVD in CKM) | Individuals with established clinical CVD. 4a: Clinical CVD without end‑stage renal disease (ESRD); 4b: Clinical CVD complicated by ESRD. | Self-reported coronary heart disease, heart attack, heart failure, or stroke.  4a: Clinical CVD and eGFR ≥15 mL/min/1.73m²  4b: Clinical CVD and eGFR <15 mL/min/1.73m² |

***Table S5. Definition and operational definitions of 0-4 stages of cardiovascular‑kidney‑metabolic (CKM) syndrome in CHARLS cohort.***

Staging follows a hierarchical, mutually‑exclusive order (Stage 4 > Stage 3 > Stage 2 > Stage 1 > Stage 0). Some guideline‑recommended biomarkers (urinary albumin‑to‑creatinine ratio, cardiac troponin, natriuretic peptides, arterial imaging) were unavailable in CHARLS; therefore, staging was adapted to accessible anthropometric, laboratory, and self‑reported clinical variables.

*Abbreviations:* CKM, cardiovascular‑kidney‑metabolic; CHARLS, China Health and Retirement Longitudinal Study; BMI, body mass index; eGFR, estimated glomerular filtration rate; CVD, cardiovascular disease; KDIGO, Kidney Disease: Improving Global Outcomes; ESRD, end‑stage renal disease.

**Part 4.** **Definitions of Frailty Index**

A 30-item frailty index (FI) was constructed using CHARLS data following the standard deficit-accumulation framework. The 30 health deficits covered eight domains: chronic diseases, sensory function, self-rated health, basic activities of daily living (BADLs), instrumental activities of daily living (IADLs), physical mobility limitations, depressive symptoms, and cognitive performance.

All chronic disease, functional difficulty, sensory impairment and depressive symptom items were recoded as binary deficit indicators (1=deficit present, 0=no deficit). Cognitive function was transformed into a continuous deficit score ranging from 0 to 1, calculated as the sum of memory test and orientation test scores divided by 14. The final FI value was the average of all non-missing deficit items for each participant. Individuals with fewer than 24 valid non-missing deficit items out of the 30 total items were assigned a missing FI and excluded from all FI-related analyses. Full operational definitions of each deficit component are presented in Table S6.

| **No.** | **Category** | **Deficit item description** | **Coding rule for deficit** |
| --- | --- | --- | --- |
| 1 | Chronic disease | Physician-diagnosed hypertension | 1=Yes; 0=No |
| 2 | Chronic disease | Physician-diagnosed diabetes | 1=Yes; 0=No |
| 3 | Chronic disease | Physician-diagnosed cancer | 1=Yes; 0=No |
| 4 | Chronic disease | Physician-diagnosed arthritis | 1=Yes; 0=No |
| 5 | Chronic disease | Physician-diagnosed chronic lung disease | 1=Yes; 0=No |
| 5 | Chronic disease | Physician-diagnosed chronic lung disease | 1=Yes; 0=No |
| 6 | Chronic disease | Physician-diagnosed asthma | 1=Yes; 0=No |
| 7 | Chronic disease | Physician-diagnosed psychiatric/nervous/emotional disorders | 1=Yes; 0=No |
| 8 | Chronic disease | Physician-diagnosed memory-related disease | 1=Yes; 0=No |
| 9 | Sensory function | Self-reported vision problems | 1=Yes; 0=No |
| 10 | Sensory function | Self-reported hearing problems | 1=Yes; 0=No |
| 11 | Self-rated health | Self-reported general health status | 1=Poor/Fair; 0=Excellent/Very good/Good |
| 12 | BADL | Difficulty dressing independently | 1=Yes; 0=No |
| 13 | BADL | Difficulty bathing/showering independently | 1=Yes; 0=No |
| 14 | BADL | Difficulty eating independently | 1=Yes; 0=No |
| 15 | BADL | Difficulty getting in/out of bed independently | 1=Yes; 0=No |
| 16 | BADL | Difficulty using the toilet independently | 1=Yes; 0=No |
| 17 | IADL | Difficulty managing money independently | 1=Yes; 0=No |
| 18 | IADL | Difficulty taking medications independently | 1=Yes; 0=No |
| 19 | IADL | Difficulty grocery shopping independently | 1=Yes; 0=No |
| 20 | IADL | Difficulty preparing meals independently | 1=Yes; 0=No |
| 21 | IADL | Difficulty completing housework independently | 1=Yes; 0=No |
| 22 | Physical mobility limitation | Difficulty walking 100 yards | 1=Yes; 0=No |
| 23 | Physical mobility limitation | Difficulty standing up after prolonged sitting | 1=Yes; 0=No |
| 24 | Physical mobility limitation | Difficulty climbing multiple flights of stairs without rest | 1=Yes; 0=No |
| 25 | Physical mobility limitation | Difficulty lifting/carrying objects over 10 jin | 1=Yes; 0=No |
| 26 | Physical mobility limitation | Difficulty picking up a coin from a table | 1=Yes; 0=No |
| 27 | Physical mobility limitation | Difficulty stooping, kneeling or crouching | 1=Yes; 0=No |
| 28 | Physical mobility limitation | Difficulty raising arms above shoulder level | 1=Yes; 0=No |
| 29 | Depressive symptoms | CESD-10 depression scale | 1=Total score>10; 0=Total score≤10 |
| 30 | Cognitive function | Combined memory and orientation test score | Continuous 0-1 |

***Table S6. Definitions of the 30 deficit items used to construct the frailty index (FI) in CHARLS cohort.***

*Abbreviations:* FI, frailty index; ADL, activities of daily living; IADL, instrumental activities of daily living; CESD-10, 10-item Center for Epidemiologic Studies Depression Scale.

**Part 5. Base model definition.**

Cox proportional‑hazards regression models were constructed with hierarchical covariate adjustment, including crude model, Model 1, Model 2, and Model 3. Variance inflation factor (VIF) was computed across all multiply‑imputed datasets and pooled by arithmetic mean to assess multicollinearity among covariates; a VIF < 5.0 was predefined as the threshold for absence of severe multicollinearity.

Crude model: Unadjusted Cox regression model, containing only the exposure indicator of interest without any covariates.

Model 1: Adjusted for chronological age (years), sex (male/female), educational attainment (Elementary school or below / Middle school / College or above), and residence type (urban / rural).

Model 2: Adjusted for all covariates included in Model 1, plus smoking status (Never Smoker / Former Smoker / Current Smoker) and alcohol consumption (Never drinker / Former drinker / Current drinker).

Model 3 (fully‑adjusted model): Adjusted for all covariates included in Model 2, plus waist circumference (cm) and high‑density lipoprotein‑cholesterol (HDL‑C, mg/dL). VIF analysis confirmed no severe multicollinearity in Model 3 for both Light‑BA and KDM‑BA model variants (all pooled VIF < 3.0, Table S7).

The fully‑adjusted Model 3 served as the base prediction model. Metabolic‑biological age indices (TyG, Light‑BA, KDM‑BA, TyG‑Light-BA, TyG‑KDM-BA) were appended to this base model for incremental‑value and nested‑model comparisons (Table 6). Model‑fit indices (log‑likelihood, AIC, BIC, C‑index) were pooled across multiply‑imputed datasets (Table 7). ΔC‑index, continuous NRI and IDI were evaluated at 66‑month follow‑up. This time‑point ensured adequate event counts, reduced censoring and attrition bias, and offered a unified benchmark for cross‑predictor comparison. All metrics were pooled via Rubin’s rules.

| Model 3 Predictors | VIF (Light-BA Model) | VIF (KDM-BA Model) |
| --- | --- | --- |
| TyG-BA (Composite) | 2.73 | 1.64 |
| Age (years) | 2.65 | 1.62 |
| Sex | 2.24 | 2.21 |
| Educational attainment | 1.37 | 1.37 |
| Residence type | 1.17 | 1.17 |
| Smoking status | 1.97 | 1.94 |
| Alcohol consumption | 1.49 | 1.50 |
| Systolic blood pressure (mmHg) | 1.17 | 1.21 |
| Waist circumference (cm) | 1.29 | 1.18 |
| HDL-C (mg/dL) | 2.73 | 1.64 |

***Table S7. VIF for Multivariable Cox Proportional Hazards Model 3***

Note: VIF values were calculated across all m imputed datasets and pooled as arithmetic means.

A VIF value < 5.0 indicates the absence of severe multicollinearity among the covariates in the fully adjusted model.

*Abbreviations:* TyG, triglyceride‑glucose index; KDM-BA, Klemera‑Doubal method biological age; HDL‑C, high‑density‑lipoprotein cholesterol.

**Part 6. Assessment of proportional‑hazards assumption.**

Proportional‑hazards (PH) assumption for Cox proportional‑hazards regression models was assessed using Schoenfeld residual‑based tests implemented in the ‘survival’ R package. Separate PH diagnostics were performed for the Light‑BA and KDM‑BA primary models on each multiply‑imputed dataset, including variable‑specific tests and model‑specific global PH tests. We summarized the mean χ² statistics across imputed datasets, median P‑value, and proportion of imputed datasets yielding *P* < 0.05. A predictor was considered to satisfy the PH assumption when median *P*‑value ≥ 0.05 across imputations. Results of PH assumption diagnostics were presented in Table S8.

| **Predictor** | **Mean χ²** | **Median *P*‑value** | **PH assumption** |
| --- | --- | --- | --- |
| **Light‑BA Model** |  |  |  |
| GLOBAL (overall test) | 7.375 | 0.832 | Satisfy PH |
| Age (years) | 0.196 | 0.658 | Satisfy PH |
| HDL‑C (mg/dL) | 0.006 | 0.937 | Satisfy PH |
| Sex | 0.006 | 0.936 | Satisfy PH |
| TyG‑Light‑BA composite (per SD) | 0.228 | 0.633 | Satisfy PH |
| Waist circumference (cm) | 2.176 | 0.140 | Satisfy PH |
| Alcohol consumption | 0.450 | 0.799 | Satisfy PH |
| Educational attainment | 1.926 | 0.382 | Satisfy PH |
| Residence type | 1.755 | 0.185 | Satisfy PH |
| Smoking status | 0.751 | 0.687 | Satisfy PH |
| **KDM‑BA Model** |  |  |  |
| GLOBAL (overall test) | 7.392 | 0.831 | Satisfy PH |
| Age (years) | 0.211 | 0.646 | Satisfy PH |
| HDL‑C (mg/dL) | 0.006 | 0.938 | Satisfy PH |
| Sex | 0.005 | 0.946 | Satisfy PH |
| TyG‑KDM‑BA composite (per SD) | 0.042 | 0.837 | Satisfy PH |
| Waist circumference (cm) | 2.103 | 0.147 | Satisfy PH |
| Alcohol consumption | 0.396 | 0.820 | Satisfy PH |
| Educational attainment | 1.896 | 0.388 | Satisfy PH |
| Residence type | 1.722 | 0.189 | Satisfy PH |
| Smoking status | 0.750 | 0.687 | Satisfy PH |

***Table S8. Proportional‑hazards assumption testing using Schoenfeld residuals for primary Model 3.***

The global‑test assesses the overall proportional‑hazards assumption for each corresponding Cox model. Median *P* ≥ 0.05 across imputations indicates.

**Part 7. Incremental predictive performance and discrimination analyses.**

The incremental predictive value of the TyG-BA composites beyond a prespecified base model was evaluated using inverse probability of censoring weighting (IPCW)-adjusted changes in the C-index, integrated discrimination improvement (IDI), and continuous net reclassification improvement (NRI) at 96 months of follow-up. The 96-month time point was prespecified as the primary prediction horizon because it provided a balance between long-term risk assessment, the accumulation of incident cardiometabolic multimorbidity events, and the extent of censoring in this cohort. Sensitivity analyses of the IPCW-adjusted ΔC-index were additionally conducted at 60 and 108 months to assess the robustness of incremental discrimination across different follow-up durations.

Time-dependent receiver operating characteristic analyses were performed using the timeROC package in R to estimate time-dependent area under the curve (AUC) and corresponding 95% confidence intervals at 3, 5, and 8 years. These time points were selected to characterize the discrimination of the candidate markers across short-, intermediate-, and longer-term follow-up. Pre-specified pairwise comparisons of time-dependent AUCs were performed using the independent-increment-based variance framework implemented in timeROC::compare(). Comparisons were restricted to clinically relevant marker pairs, including comparisons of the TyG-BA composites with TyG and selected alternative TyG-based composites. To account for multiple comparisons, the Benjamini-Hochberg false discovery rate (BH-FDR) procedure was applied within each time point.

Decision curve analysis was performed across the full 108-month follow-up to evaluate the potential clinical net benefit of models incorporating the TyG-BA composites across a range of threshold probabilities. Kernel SHAP analyses based on Cox models were conducted as supplementary model-interpretability analyses. All statistical tests were two-sided, with P < 0.05 considered statistically significant.
